# Sequential pembrolizumab cooperates with platinum/5FU to remodel the tumor microenvironment in advanced gastric cancer: a phase II chemoimmunotherapy trial

**DOI:** 10.1101/2023.04.03.23288062

**Authors:** Minae An, Arnav Mehta, Byung Hoon Min, You Jeong Heo, Milan Parikh, Lynn Bi, Razvan Cristescu, Hyuk Lee, Taejun Kim, Song-Yi Lee, Jeonghyeon Moon, Ryan J. Park, Matthew R. Strickland, Woong Yang Park, Won Ki Kang, Kyoung-Mee Kim, Seung Tae Kim, Samuel J. Klempner, Jeeyun Lee

## Abstract

Adding anti-PD1 antibodies to 5-FU/platinum chemotherapy improves survival in a subset of advanced gastroesophageal adenocarcinoma (GEA) patients. Beyond PD-L1 expression and mismatch repair status we have limited insight into molecular predictors of response or the relative contribution of PD-1 blockade. We conducted an investigator sponsored phase II trial (n = 47) sequentially adding pembrolizumab to standard 5-FU/platinum in previously untreated advanced GEA (ClinicalTrials.gov: NCT04249739). With an overall response rate of 67% the activity paralleled phase III chemoimmunotherapy trials. To understand on-treatment tumor and immune adaptations patients underwent serial biopsy of the primary tumor, including baseline, after one cycle of 5-FU/platinum, and after the addition of pembrolizumab. We leveraged transcriptional profiling from 358,067 cells to identify multicellular networks of malignant, stromal, and immune cells after chemotherapy and concurrent chemoimmunotherapy. The relative usage of pro-tumor and anti-tumor interaction hubs differed between fast and slow progressing patients. Chemotherapy induced early on-treatment formation of hubs centered on tumor-reactive T-cell and M1-oriented macrophage interactions with pro-inflammatory cytokines in slow progressors. Faster progression was characterized by increased MUC5A and MSLN containing programs in tumor cells and M2-oriented macrophages with immunosuppressive stromal interactions. After adding pembrolizumab we observed increased CD8 T-cell infiltration by scRNAseq and multiplex immunofluorescence and development of an immunity hub involving co-variation of the tumor-reactive CXCL13 program and epithelial interferon-stimulated gene programs enriched in slow progressors. Together this data provides prospective evidence of differential early on-treatment evolution of the gastric immune microenvironment and nominates candidate cellular interactions for clinical targeting.

## Introduction

The frontline treatment of advanced gastroesophageal adenocarcinomas (GEA) has rapidly transitioned toward the broad adoption of concurrent chemoimmunotherapy in the past two years. Unlike melanoma and NSCLC where a large portion of patients benefit from anti-PD-1 antibody monotherapy (aPD1), the activity of aPD1 in gastroesophageal adenocarcinomas is modest and heterogeneous^1, 2^. The change in treatment paradigm for GEA has outpaced our understanding of biomarkers to optimize patient selection and drug matching; as such, biomarkers beyond PD-L1 expression are needed to select immunotherapy responders, especially non-invasive biomarkers that may be used to understand early on-treatment resistance^1^. Importantly, the optimal cooperativity between different treatment modalities (e.g. chemotherapy, immunotherapy, targeted agents) is understudied, and we are therefore limited in rationally designing novel combination treatments or sequencing strategies. This is in large part due to suboptimal preclinical model systems and trial designs that use concurrent chemoimmunotherapy, thus making it difficult to dissect the relative contribution of individual modalities. Additionally, larger studies often contain limited correlative efforts to examine the effects on tumors of aPD1 and standard 5FU and platinum chemotherapy. To develop the next generation of gastroesophageal trial concepts we need a deeper understanding of our current paradigms.

To address this knowledge gap we previously conducted a small pilot serial biopsy study in metastatic gastric cancer (GC) patients treated with standard 5-FU/platinum (XELOX) chemotherapy and demonstrated important tumor microenvironment (TME) remodeling^3^. Several features including M1/M2 macrophage repolarization and CD8+ T-cell influx were conserved among patients responding to standard 5-FU/oxaliplatin. Similar findings have been seen in some post-treatment surgical studies from non-metastatic samples, highlighting a predictive and prognostic role for TME composition^4, 5^. These findings support an ability of chemotherapy to generate a favorable environment in which to introduce immunomodulatory approaches, but were limited by lack of aPD1 incorporation, small sample size, and retrospective analyses.

The immune cells of the TME can assume a large variety of phenotypes which can be defined by transcriptomic features and co-expression of multiple proteins. While modern spatial profiling technologies provide important insights into the spatial organization of the TME, substantial biology can be observed with TME reconstruction from bulk and scRNAseq ^6, 7^. Recently, co-varying transcriptional programs in single-cell RNA sequencing (scRNAseq) data have been leveraged to predict multicellular interaction networks, or hubs, several of which have been spatially localized within tumors^8–12^. While these studies have identified important insights into baseline tumor organization in microsatellite-high colorectal (MSI-H) patients, they have lacked on-treatment paired sampling of microsatellite stable (MSS) patients to understand how these hubs may evolve under therapeutic pressures, and whether remodeling differs between patients who benefit and/or do not benefit from a given approach. We therefore designed a prospective single arm phase II sequential chemoimmunotherapy trial in metastatic GC patients with serial sampling for deep correlative studies to build upon our prior observations. In the framework of this clinical trial, we sought to: 1) validate TME remodeling of 5-FU/platinum chemotherapy, 2) understand the contribution of pembrolizumab on top of standard chemotherapy, 3) explore peripheral surrogates for tissue level immune cell remodeling, and 4) identify candidate pathways and cell types to nominate for further exploration and rational combination studies.

## Results

### Clinical activity and molecular characteristics

Forty-seven treatment naive advanced GC (AGC) patients were enrolled between September 2020 and April 2022 and the overall schema and sample collection strategy is outlined in **Figure 1A**. All tissue biopsies were taken from the primary tumor, and an attempt was done to biopsy the same sites in all paired samples using endoscopic mapping. The median age of the patients was 55 years (range 34–75 yr), and the majority of patients were men (80.9%). All patients were Korean. Four patients (8.5%) had HER2-positive AGC. Forty-three patients (91.5%) received 5-FU/platinum chemotherapy and four (8.5%) received 5-FU/platinum chemotherapy with trastuzumab as frontline treatment per standard of care (Table S1). Demographic features are shown in **Table S1**. The primary endpoint was objective response rate by RECIST v1.1. At the data cut of April 21, 2022 the overall response rate (ORR) was **68%** (**Figure 1B and S1A,B**), and with a median follow up of 10.7 months the median progression free survival (PFS) and overall survival (OS) was 7.7 months and not reached, respectively (**Figure S1C,D**). The clinical activity is aligned with data from current phase III concurrent chemoimmunotherapy trials in Asian and Western patients^2, 13^.

**Figure 1.**
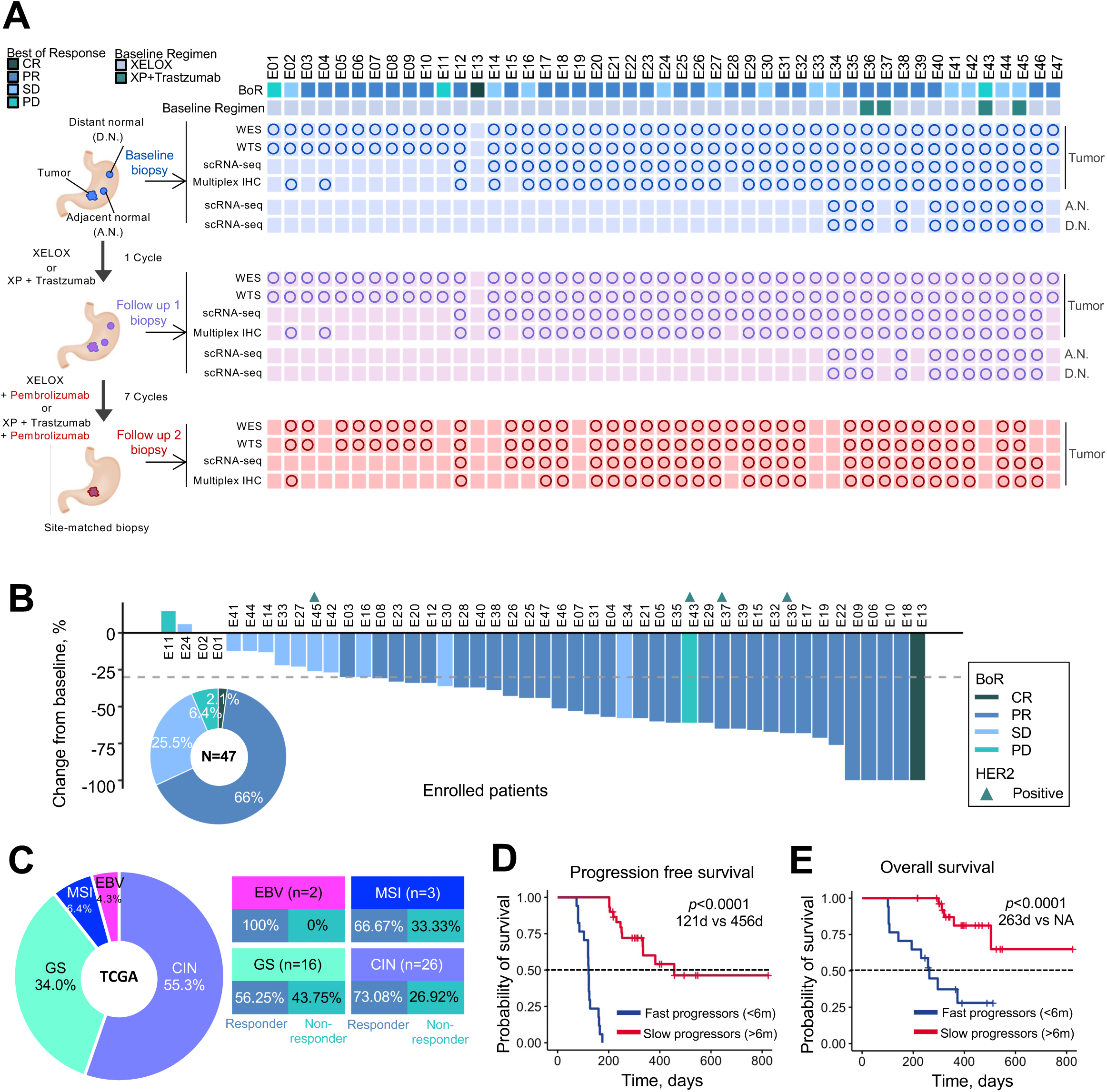
Phase II trial results and sample collection schema. (A) Sample collection schedule and analysis platforms in a phase II sequential chemoimmunotherapy trial. Circles correspond to samples included in analyses. (B) Waterfall plot demonstrating RECISTv1.1 response for patients in the trial. (C) Clinical trial patient composition and response rates by TCGA molecular subgroup. (D) Kaplan-Meir curve showing progression-free survival (PFS) by fast and slow progressor categorization. Statistical comparison performed using a log-rank test. (E) Kaplan-Meir curve showing overall survival (OS) by fast and slow progressor categorization. Statistical comparison performed using a log-rank test.

The enrolled patient population is representative of the established genomic landscape described in the The Cancer Genome Atlas (TCGA) and Asian Cancer Research Group classifications (**Figure S2**)^14, 15^. Two patients had Epstein-Barr virus (EBV)-positive (EBV+) tumors (4.3%), three patients had tumors with microsatellite instability (MSI-high) (6.4%), sixteen patients had genomically stable (GS) tumors (34%) and twenty-six patients had chromosomally instable (CIN) tumors (55%) (**Figure 1C**). Consistent with prior reports from our group, nearly all EBV+ or MSI-high patients achieved CR or PR^16, 17^. Because RECIST stable disease (SD) can encompass heterogeneous clinical benefits (i.e. some SD patients achieve durable control) we additionally analyzed patients by PFS, with slow progressors defined as PFS defined as > 6 months (median PFS and OS was 121 and 263 days, respectively) and fast progressors as PFS < 6 months (median PFS and OS was 456 days and not-reached, respectively)^18^. The median PFS of 6 months was selected from modern phase III frontline trials and we observed a shorter overall survival in fast progressors (**Figure 1D,E**)^13, 18–20^. There were no genomic features predictive of chemotherapy response, consistent with prior studies^3, 21^. TMB was not significantly associated with clinical response in our cohort (**Figure S2**).

### A single dose of 5-FU/platinum remodels cell type composition in the TME

Given the lack of genomic chemotherapy response predictors we sought to broadly understand the effects of standard platinum/5FU on the tumor immune microenvironment. Our prior pilot work had suggested that 5FU/platinum leads to increased CD8+ T cell infiltration and can reprogram macrophage populations^3^. Comparing baseline (BL) and follow up 1 (after 1 cycle of chemotherapy; FU1), bulk RNA analyses confirmed broad upregulation of T-cell trafficking pathways, NK and T-cell populations and Th1 signatures associated with IFNg production (**Figure 2A,B and S3A)**. Gene sets related to regulatory cells (including regulatory T cells [Treg], checkpoint inhibition and tumor associated macrophages [TAM]) were also significantly higher at FU1 though the magnitude of change was lesser (**Figure S3B**). We additionally observed on-treatment enrichment of pathways involved in PD1 signaling, TCR and co-stimulatory signaling by gene set variation analysis (GSVA) (**Figure S3A**). To confirm tumor level compositional changes after chemo we performed imaging mass cytometry (**Figure S3C and S4A,B**). Consistent with bulk transcriptome data, we observed an increase in CD8+ lymphocyte infiltration during chemotherapy (**Figure S3C and S4A**). Without informing our analyses by clinical categories (responder vs non-responder or fast vs slow progressor) our data confirms bulk immune cell compositional changes after a single cycle of standard 5FU/oxaliplatin chemotherapy.

**Figure 2.**
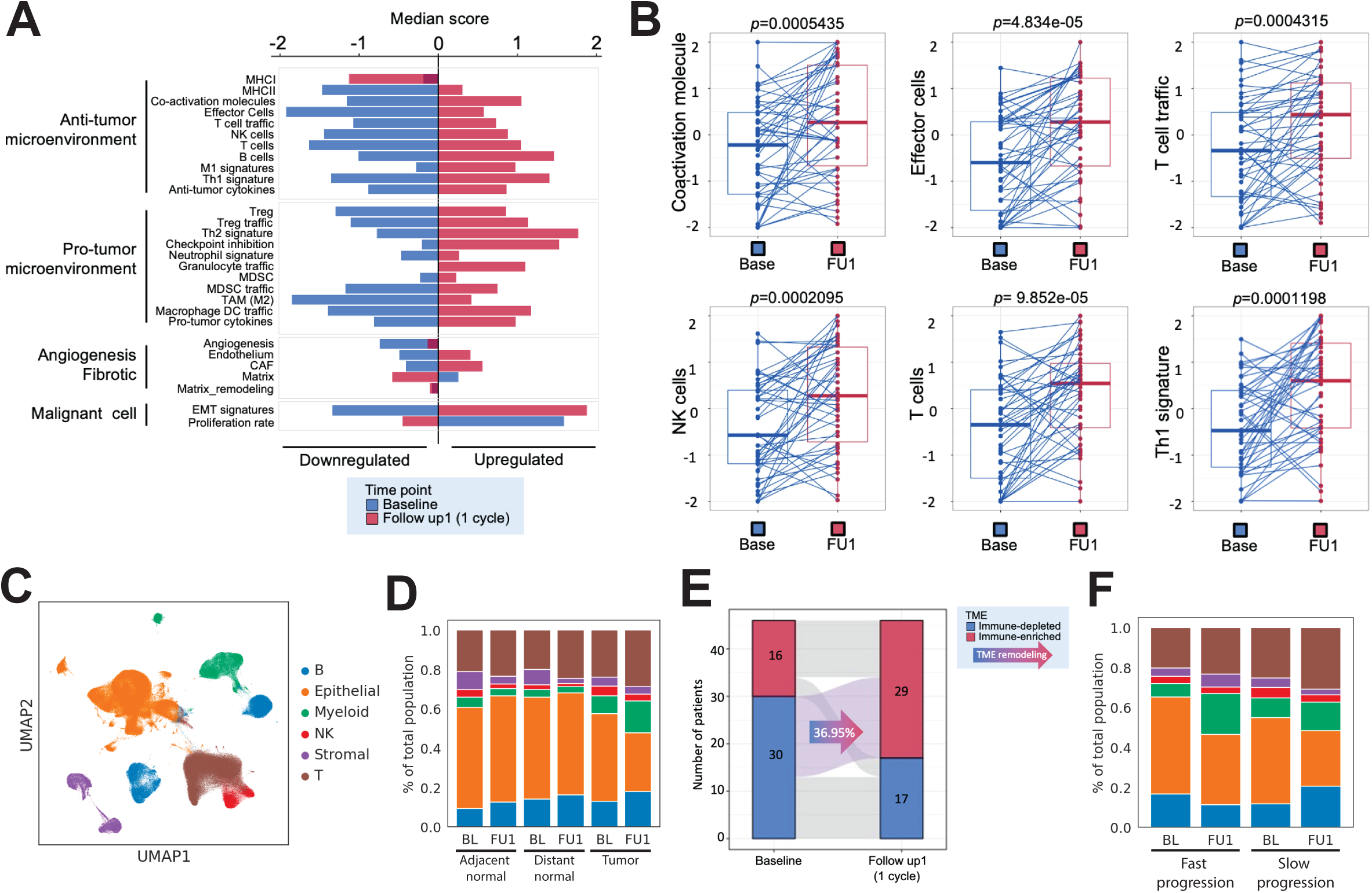
A single cycle of 5FU/platinum remodels the TME in advanced gastric cancer. (A) Pathway enrichment analysis on differentially expressed genes comparing bulk RNA-seq profiles for samples obtained pre-treatment (baseline) and samples obtained after 1 cycle of chemotherapy (follow up 1; FU1). (B) Changes in enrichment in bulk RNA-seq data of immune-related pathways from baseline (Base) to FU1. Statistical comparison performed using a Wilcoxon signed-rank test. (C) UMAP embedding of single cell transcriptomes obtained from all samples in this trial. Labeled are canonical cell types. (D) Cell type proportions, obtained from scRNAseq data, in adjacent normal, distance normal and tumor tissue, at baseline (BL) and after 1 cycle of chemotherapy (FU1). (E) Redistribution of TME subtypes following one cycle of 5FU/platinum chemotherapy. TME subtypes were obtained using a classification performed on bulk RNA-seq data. (F) Cell type proportions, obtained from scRNAseq data, in tumor samples of fast and slow progressing patients at BL and FU1.

**Figure 3.**
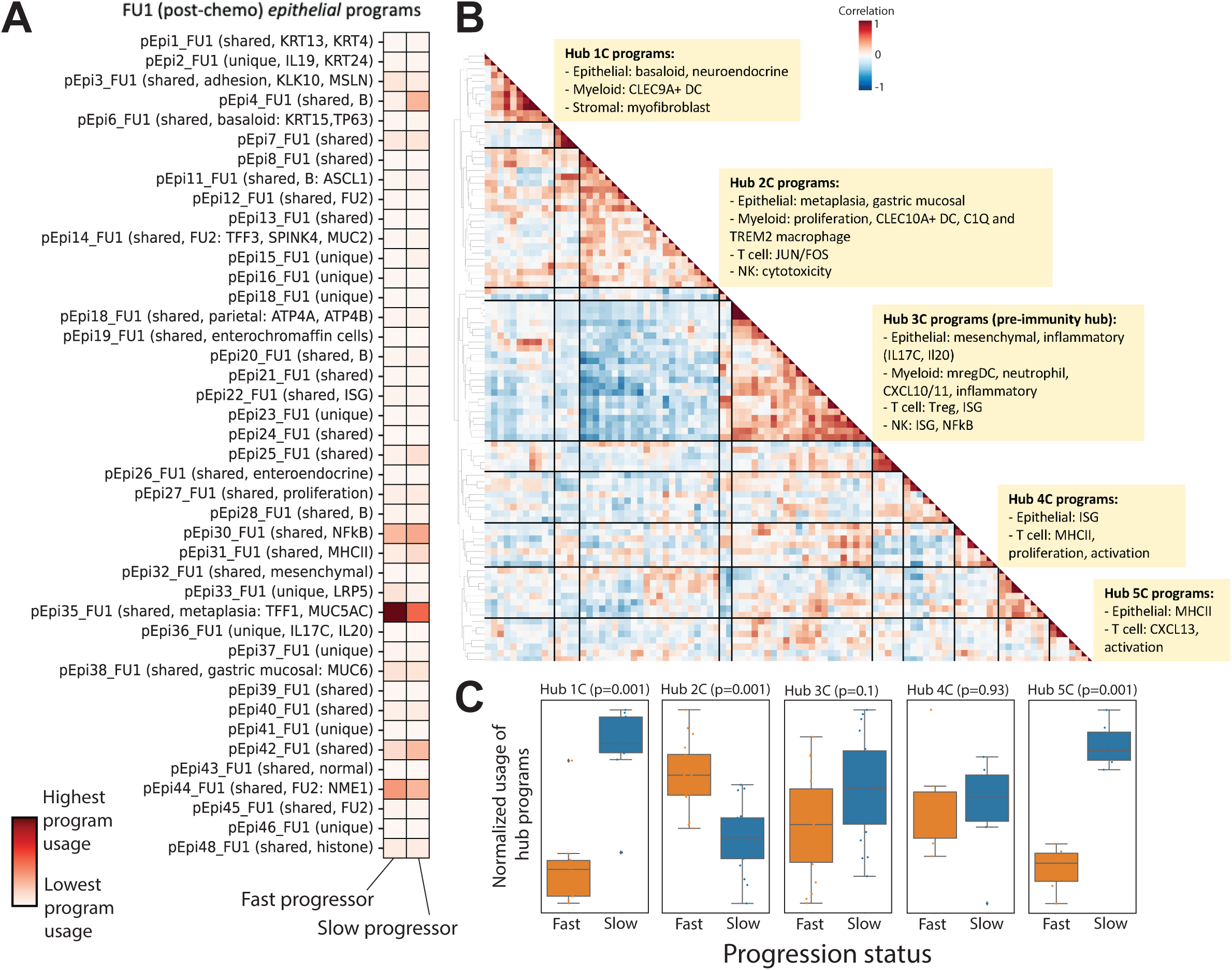
Identification of co-varying gene programs that underlie chemotherapy resistance and response. (A) cNMF was performed on epithelial cells at FU1. Shown is the mean usage of each cNMF gene program in the epithelial cells of fast and slow progressing patients. (B) Heatmap showing pairwise correlation of gene program activities across all patient samples at FU1 using the 90th percentile of patient-level program activity in epithelial, myeloid, T, NK and stromal cells. Hierarchical clustering was performed to identify clusters of co-varying proteins, which have been labeled as Hub1C to 5C. (C) Average z-scored usage of all gene programs in each hub split by fast and slow progressing patients. Statistical comparison performed using a two-sample t-test with Bonferroni correction.

**Figure 4.**
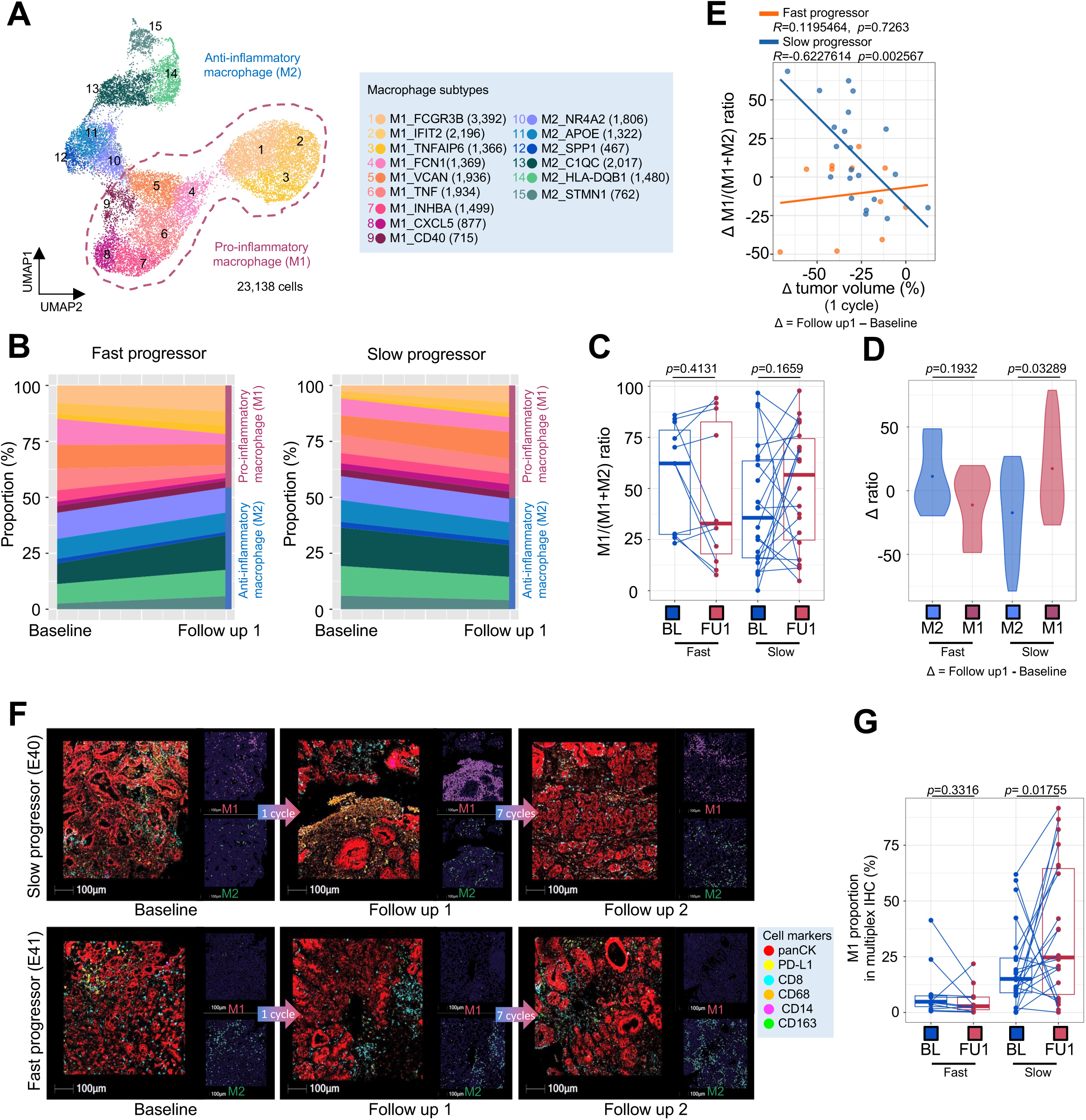
Chemotherapy leads to macrophage repolarization in patients with favorable response. (A) UMAP embedding of single cell transcriptomes of all macrophages from all samples in this trial. Labeled are granular macrophage subtypes, including designation of M1 and M2 subtypes. (B) Macrophage subtype proportions, obtained from scRNAseq data, at BL and FU1 in fast versus slow progressing patients. (C) Relative proportion of M1 macrophages of all macrophages, obtained from scRNAseq data, at BL and FU1 in fast and slow progressing patients. Statistical comparison performed using a Wilcoxon signed-rank test. (D) Change in M1 and M2 macrophage proportions from BL to FU1, obtained from scRNAseq data, in fast and slow progressing patients. Statistical comparison performed using a Wilcoxon signed-rank test. (E) Change in relative M1 proportion from BL to FU1 plotted against change in tumor volume after 1 cycle of chemotherapy, segregated by fast and slow progressing patients. (F) Multiplexed immunofluorescence (mIF) images of BL, FU1 and FU2 samples from two patients, E40 (slow progressor) and E41 (fast progressor), staining for panCK, PD-L1, CD163, CD68, CD14 and CD8. (G) Proportion of M1 macrophages, obtained from mIF images, at BL and FU1 in fast versus slow progressing patients. Statistical comparison performed using a Wilcoxon signed-rank test.

We then moved to identify granular cell types, substates and gene programs that underlie treatment response and resistance by leveraging our paired scRNAseq data from BL and FU1. We examined single-cell gene expression profiles of 138 samples (BL tumor [*n*=33], BL normal [adjacent normal, *n*=11; distant normal, *n*=11], FU1 tumor [*n*=33], FU1 normal [adjacent normal, *n*=11; distant normal, *n*=11] and FU2 tumor [*n*=28]). After filtering low-quality cells, we collected a total of 358,067 cells. We performed unsupervised clustering and identified six major cell types using canonical marker genes, including epithelial cells, stromal cells and immune cells (myeloid, T lymphoid, NK cells, B lymphoid) (**Figure 2C and S5**)^3, 17, 22–24^. Epithelial cells were categorized as tumor or normal cells using known marker genes of tumor cells and inferred copy-number variant (CNV) profiles with two methodologies, inferCNV^25^ and Numbat^26^ (**Figure S6,7**). Supportive of the inferCNV classification we found in the 4 patients diagnosed with HER2+ disease by IHC, there was clear, yet heterogeneous amplification predicted at the HER2 locus on chromosome 17; interestingly, we found smaller subclonal populations with putative HER2 amplification in a subset of patients found to have HER2-disease (**Figure S7**).

**Figure 5.**
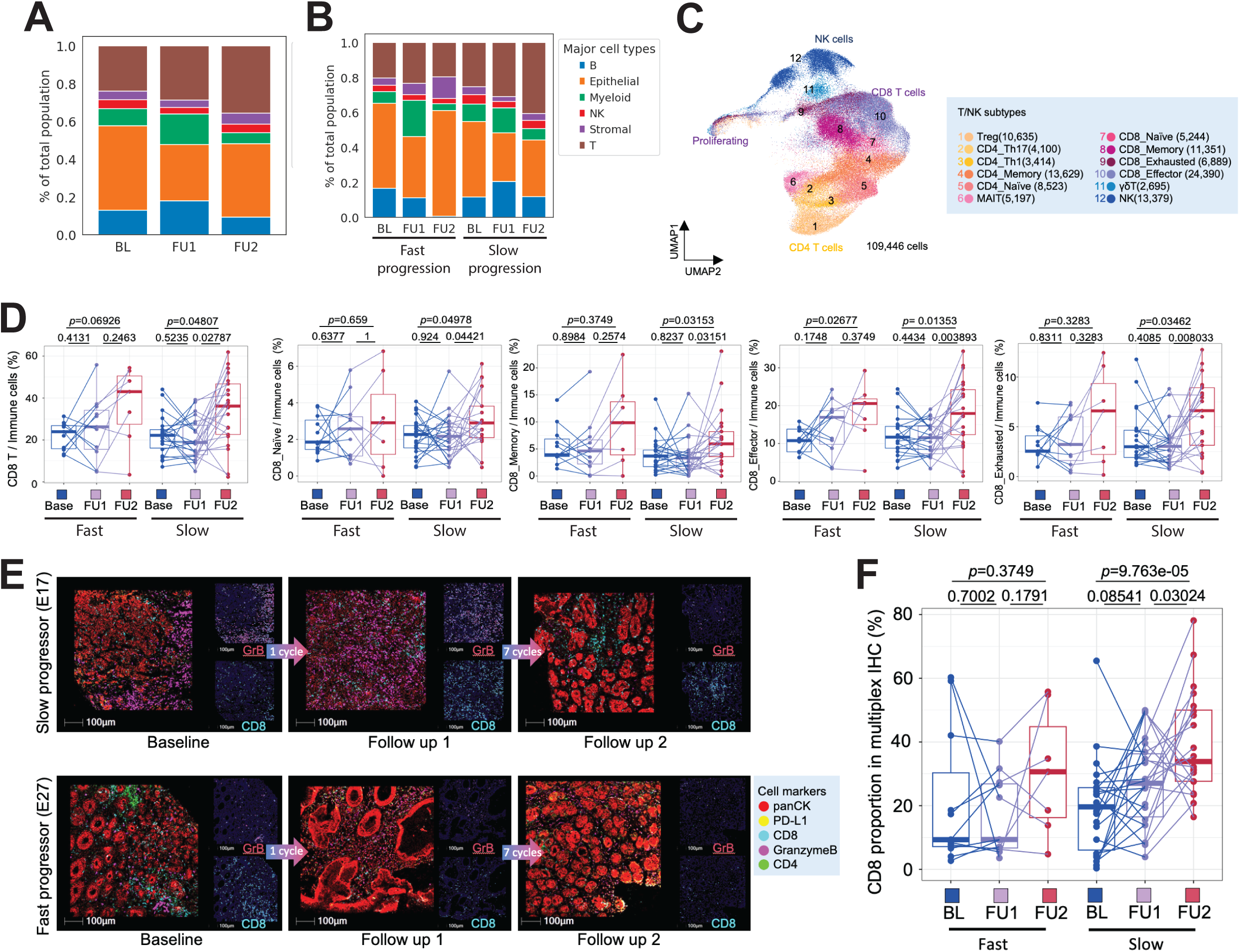
The addition of immunotherapy to 5-FU/platinum chemotherapy redistributes T cell phenotypes. (A) Cell type proportions, obtained from scRNAseq data, of all tumor samples at BL, FU1 and after immunotherapy treatment (FU2). (B) Cell type proportions, obtained from scRNAseq data, in tumor samples of fast and slow progressing patients at BL, FU1 and FU2. (C) UMAP embedding of single cell transcriptomes of all T and NK cells from all samples in this trial. Labeled are granular T and NK cell subtypes. (D) Cell type proportions as a proportion of all immune cells, obtained from scRNAseq data, of total, naive, memory, effector and exhausted CD8 T cells. Statistical comparisons performed using a Wilcoxon signed-rank test. (E) Multiplexed immunofluorescence (mIF) images of BL, FU1 and FU2 samples from two patients, E17 (slow progressor) and E27 (fast progressor), staining for panCK, PD-L1, CD4, CD8 and Granzyme B. (F) Proportion of CD8 T cells macrophages, obtained from mIF images, at BL, FU1 and FU2 in fast versus slow progressing patients. Statistical comparison performed using a Wilcoxon signed-rank test.

Consistent with tumor cell death we observed a contraction of the epithelial tumor component and expansion of myeloid and T cell compartments after 1 cycle of chemotherapy without significant changes in adjacent or distant normal samples (**Figure 2D**). We then classified all tumor samples into two distinct TME subtypes, immune-depleted or immune-enriched, using published TME subtypes associated with aPD1 outcomes^27^. Seventeen patients (36.95%) converted from immune-depleted subtype to immune-enriched subtype after one cycle of chemotherapy treatment consistent with favorable TME remodeling (**Figure 2E**). Interestingly, when stratified by fast vs slow progressors we noted a greater expansion of myeloid and stromal cells in fast progressors, and a greater expansion of T and B cells in slow progressors suggesting even a single dose of chemotherapy may create a favorable TME trajectory in some patients, aligned with known predictors of IO benefit (**Figure 2F**)^28^.

### 5-FU/platinum globally alters epithelial, stromal and inflammatory programs

To understand the gene expression programs induced within each cell type by a single dose of chemotherapy we performed consensus non-negative matrix factorization (cNMF) on each cell type at each timepoint individually, and identified between 9 to 48 programs for each cell type at each timepoint (**Figure S8-10 and Table S2**)^12, 29, 30^. To identify programs that were shared across timepoints for any given cell type, we performed a hypergeometric test of the top 100 weighted genes for all pairs of programs, yielding a subset of programs conserved across treatment conditions and another subset that was unique to each treatment condition (**Table S2**). Among the shared programs across timepoints, we found several specific to particular immune or stromal cell subsets, such as a myofibroblast program (pS2_B, pS5_FU1), a Treg program (pT12_B, pT3_FU1, pT6_FU2), a myeloid regulatory dendritic cell program (mregDC; pM8_B, pM17_FU1, pM20_FU2) and a plasmacytoid dendritic cell program (pDC; pM6_B, pM4_FU1, pM8_FU2), among others (**Figure S9,10**). Several shared programs were expressed across cell subtypes, such as stromal MHCII (pS1_B, pS11_FU1, pS17_FU2) and proliferation programs (pS3_B, pS9_FU1, pS9_FU2), myeloid inflammatory, interferon-stimulated gene (ISG) and interleukin 1 (IL1) programs (pM2_B, pM10_FU1, pM13_FU1, pM13_FU2, pM14_FU2), and T cell translation (pT1_B, pT10_FU1), HSP (pT4_B, pT12_FU1, pT8_FU2), NFkB/JUN/FOS programs (pT11_B, pT15_FU1, pT3_FU2) and MHCII, IFNg/ISG and activation programs (pT14_B, pT9_FU1, pT13_FU1, pT14_FU1, pT11_FU2) (**Figure S9,10**).

Importantly, we found a subset of shared epithelial programs across many patients at each timepoint (e..g pEpi3_B, among others), which included basaloid (pEpi7_B, pEpi6_FU1), parietal (pEpi9_B), proliferation (pEpi10_B), KRT20+ (pEpi16_B), gastric mucosal/MHCII (pEpi17_B) and adhesion programs (pEpi19_B) (**Figure S8**). We found epithelial mesothelin (pEpi3_FU1), ISG (pEpi22_FU1) and metaplasia (pEpi35_FU1) programs that were present only after chemotherapy treatment, suggesting these may reflect adaptive epithelial responses to treatment. Other programs, such as a neuronal-like (pEpi13_B, pEpi11_FU1) program marked by ASCL1 expression, were present across timepoints but were largely sample-specific. We next sought to identify programs that were differentially induced in epithelial cells when comparing fast and slow progressing patients (**Figure 3A and S11,S12**). We found that the metaplasia program, marked by TFF1^31^ and MUC5AC^32^ as top weighted genes, and the adhesion program, marked by MSLN as a top weighted gene, had significantly higher usages in epithelial cells post-chemotherapy (FU1) in fast progressors compared to slow progressors (**Figure 3A and S11A**). This potentially implicates chemotherapy-induced metaplastic programs as a poor prognostic marker and this difference in metaplasia program usage between fast and slow progressing patients was maintained after immunotherapy (**Figure S11B**). Recently, MUC5AC was observed to be enriched among colorectal cancer patients with peritoneal disease, a particularly poor prognosis and fast progressing clinical subgroup^33^. Similarly, MSLN expression is a poor prognostic feature and an active area of investigation for cellular therapies^34, 35^.

### 5-FU/platinum remodels cellular interaction networks and drives multicellular hubs in the TME

We next sought to identify cellular programs across all cell types that may influence each other after a single dose of chemotherapy. Toward this end, we employed a methodology to identify groups of cell type specific programs that co-varied across all samples at a given time point. For each sample, we quantified the program usage of each program within the cell type it was identified, focusing on epithelial, stromal, myeloid, T and NK cells. We subsequently looked at the correlation of these program usages over all samples and identified groups of co-varying programs (termed ‘multicellular hubs’) (**Figure S13-15 and Table S3**). We found several multicellular hubs at baseline (**Figure S13**) and after chemotherapy (**Figures 3B and S14**). Most notably, we found five major hubs after chemotherapy each with distinct properties (**Figure 3B**). This included: 1) covarying epithelial basaloid and myofibroblast programs (Hub 1C); 2) epithelial metaplasia, myeloid proliferation and C1Q and TREM2 macrophage programs (Hub 2C); 3) epithelial mesenchymal and inflammatory, mregDC, myeloid CXCL10/11, Treg and T cell ISG programs (Hub 3C); 4) epithelial ISG and T cell activation programs (Hub 4C); and 5) epithelial MHCII and T cell CXCL13 and activation programs (Hub 5C).

Hubs 3C and 5C, while distinct at this time point, appeared to collectively include the components of a recently described multicellular immunity hub^9^ that includes a positive feedback loop between CXCL10/11 expressing myeloid and tumor cells, and ISG expressing and tumor-reactive CXCL13+ T cell subsets^9, 36, 37^. These results suggest that a single cycle of standard 5-FU/platinum may steer the formation of key immunity hubs in GEA. We therefore hypothesized that the composition and degree to which anti-tumor immunity hubs are induced by chemotherapy may inform the ability of additional ICB to induce a durable anti-tumor response and durable clinical benefit. We next assessed the relative representation of each hub in slow versus fast progressors; Hubs 1C, 3C and 5C programs had significantly higher usage in slow progressors compared to fast progressors (**Figure 3C**), consistent with proposed role of Hubs 3C and 5C in portending favorable immune response. Importantly, Hub 2C programs had higher usage in fast progressors compared to slow progressors (**Figure 3C**). As Hub 2C included covariation of the poorly prognostic metaplasia program with several macrophage programs we opted to further probe the role of chemotherapy treatment in altering macrophage subsets (**Figure 3B and S9A-C**)

### 5-FU/platinum reprograms macrophages subsets

Previously, we had observed a role for early on-treatment tumor associated macrophage (TAM) orientation in identifying clinical responders in GEA, and a relationship between TAMs and T cell exhaustion programs in cancer is well described, particularly in the inner regions of tumors^38, 39^. Although overly simplistic, the M1 vs M2 framework is useful for conceptualizing TAM populations, and M2 oriented TAMs can impair antigen presentation and limit T cell activation^38, 40, 41^. In this context we explored TAM associations with response. We first performed a high-level clustering of all myeloid cells, identifying dendritic cell, monocyte/macrophage and mast cell populations (**Figure S16**). To further refine TAM subsets, we performed subclustering of the TAM population and identified cells divided among 16 subtype clusters (**Figure 4A and S17A,B**). To better dichotomize the TAMs, we grouped them into pro-inflammatory macrophage subsets (M1: M1_FCGR3B, M1_IFIT2, M1_TNFAIP6, M1_FCN1, M1_VCAN, M1_TNF, M1_INHBA, M1_CXCL5, M1_CD40) and anti-inflammatory subsets (M2: M2_NR4A2, M2_APOE, M2_SPP1, M2_C1QC, M2_TMEM176B, M2_CD74, M2_STMN1). Interestingly, we found that the C1Q and TREM2 macrophage programs that co-varied with the poorly prognostic epithelial metaplasia program were most highly expressed in M2 macrophage subsets (**Figure S9B**), consistent the proposed negative prognostic role of M2 macrophages. As has been seen in prior work, we found pro-inflammatory genes, such as *IL1B* and *S100A8* were significantly upregulated in M1 clusters (**Figure S17B**).

Using the M1/M2 conceptual structure we compared TAMs from tumor tissues at baseline and FU1. We observed divergent patterns of M1 proportions between fast and slow progressor groups (**Figure 4B**). In a prior pilot study we noted the relevance of the M1/M1+M2 ratio in segregating responders^3, 17^. In this prospective trial we confirmed changes in the M1/(M1+M2) ratio with increased M1/(M1+M2) ratio and greater magnitude of change in the slow progressor group, and this was consistent when doing the comparison between clinically defined RECIST responders and non-responders (**Figure 4C,D and S17C-E**). Within the M2-oriented subset SPP1 expression is associated with pro-angiogenic signaling and worse outcomes with immunotherapy^42, 43^. We examined M2-SPP1 expression and confirmed a decrease in M2-SPP1 proportions only in responders (**Figure S17F**). To highlight the clinical relevance of early on-treatment M1/(M1+M2) ratio changes we examined CT and endoscopic images. Patients with increased M1/(M1+M2) ratio showed decreased tumor volume by CT scan images, PET scan image and inflammation in the stomach by endoscopic images (**Figure 4E, S17G and S18A**). However, patients with low on-treatment M1/(M1+M2) ratio after 1 cycle of chemotherapy showed a limited response on CT scan, PET scan and endoscopy (**Figure S18B**). Given that the expansion of M1 in tissues was associated with a clinical response, we sought to confirm our scRNAseq findings with multiplex immunofluorescence (mIF). Using established macrophage lineage markers (CD14, CD68, CD163) we first confirmed early M1 expansion on treatment using multiplex immunofluorescence (mIF). Using the mIF-determined M1 proportion from 33 patients there was significant expansion during chemotherapy in the slow progressor group aligning with our scRNAseq data (**Figure 4F,G and S18C**). Overall, early on-treatment changes in macrophage subsets predicted subsequent clinical outcomes.

### Stromal cell and macrophage interactions confer an unfavorable TME

Having confirmed the clinical relevance of macrophage subset composition we wanted to more deeply understand M2 interactions with other TME components. Interestingly, we found that the four patients with increased stromal cell proportion (E24, E27, E30 and E33) were all fast progressors and had an increased proportion of M2 macrophages (**Figure S19A,B**). We next performed a subclustering of stromal subsets to and identified five distinct clusters of cells: fibroblasts, myofibroblasts, endothelial cells, pericytes and glial cells (**Figure S19C,D**). We subsequently noted that myofibroblasts are decreased in number in slow progressors at FU1, but not in fast progressors (**Figure S19E**). To further investigate how M2 macrophages might influence stromal components, and vice versa, we scored M2 macrophage subsets on signatures for phagocytosis, angiogenesis and the PDGF pathway, which is known to influence fibroblast behavior. We found APOE and C1QC macrophages to have the highest phagocytosis scores, SPP1 macrophages to have the highest angiogenesis scores, and finally for NR4A2, APOE and SPP1 macrophages to have the highest PDGF pathway scores (**Figure S20**).

To better delineate the specific cellular interactions that may underlie cell-cell interactions after chemotherapy, we applied a methodology to infer ligand-receptor pairs (CellphoneDB^44^) between cell types using scRNAseq data and applied it to each treatment time point and to normal tissues separately (**Table S4**). We found that the number of predicted interactions were highest for stromal cells with other stromal cells or with myeloid cells (**Figure S21**). We honed in on predicted L-R interactions within tumor tissues that were distinct from normal tissue (**Table S5**), and additionally were unique to FU1 samples compared to baseline samples (**Table S5**). We found that after chemotherapy, LGALS9 and SIRPa on myeloid cells served as ligands to CD47 on stromal cells, thus inhibiting phagocytosis by myeloid cells^45^. Additionally, we found several other myeloid-stromal ligand-receptor interactions that may influence macrophage polarization, including stromal cell produced E-selectin and macrophage inhibitory factor (MIF). Importantly, we found several ligands in myeloid cells shared at the baseline timepoint that may influence stromal behavior, including CD55, IL8, TNF, TGFB1 and SPP1.

### Identification of peripheral biomarkers associated with chemotherapy TME remodeling

In an effort to identify peripheral biomarkers associated with early on-treatment TME remodeling, we performed plasma proteomics (3,072 proteins) using a commercial proximity extension assay (Olink, **see Methods**) at matched timepoints to our tissue sample collections (**Figure 1A**). We found that plasma protein levels were overall poorly correlated with tissue gene expression as measured by bulk RNAseq (**Figure S22A**), and therefore individual proteins would not serve well as surrogate markers for tissue expression. We identified differential proteins associated with progression status, time on therapy (B, FU1 and FU2) and the interaction between the two using a linear mixed effect model (LMM) (**Table S6**). We had previously used this method to associate elevated plasma levels of interferon-regulated proteins with immune checkpoint inhibitor (ICI) resistance in melanoma^46^. We now found several proteins associated with fast progression, including tumor markers CEACAM5, MUC13, and MUC16, that may be associated with changes in overall tumor bulk, inflammatory markers including IL1A, IL6, IL15 and CCL19, myofibroblast markers including ACTA2, and pro-angiogenic proteins including VEGFA (**Figure S22B-F and Table S6**). Importantly, we found that using proteins differentially identified between fast and slow progressors, these two patient subsets clustered distinctly (**Figure S22C**).

To better identify co-abundant proteins that may serve as biomarkers for different treatment responder subsets we looked at the correlation of differential proteins across samples after chemotherapy treatment (FU1). We found strong correlation in abundance of proteins associated with slow progression that clustered into two subsets (**Figure S22D**), one implicated in lipid regulation (LEP, FABP9) and extracellular matrix composition (DPT), and the other that included integrin proteins (ITGAL, ITGA2) and select reported biomarkers across other tumor types including CLEC3B^47, 48^. This therefore suggests that there may be protein groups (modules) detected in the plasma that may underlie functionally conserved processes in response to chemotherapy treatment. To better elucidate this, we looked for correlations of protein abundance with bulk RNA-seq gene signatures identified as differential between BL and FU1 biopsies (**Figure S3A**). We found several proteins strongly correlated with EMT signatures, such as FLT3LG and CD276, that were also differentially abundant over time in our LMM (**Figure S23A and Table S7**). Importantly we identified several peripheral biomarkers that may serve as indicators of tumor T cell infiltration and MHC presentation (**Figure S23B and Table S7**).

Finally, for each of our differential plasma proteins with respect to progression status, we considered the expression of these proteins within subsets of cells in our scRNAseq data. We found that a majority of proteins had highest gene expression in stromal cells, with smaller subsets of proteins specific to T and NK cells, epithelial cells or shared across cell types (**Figure S24**).

### The addition of pembrolizumab enhances T-cell anti-tumor immune remodeling

We next investigated how the addition of the aPD1 agent pembrolizumab after chemotherapy may confer durable treatment responses in some patients. We first applied a validated gene expression signature predictive of pembrolizumab benefit and confirmed an increase with chemotherapy alone that was furthered by the addition of pembrolizumab across all patients (**Figure S25A**)^49–51^. After a single cycle of 5FU/oxaliplatin we observed increased expression of representative genes of immunogenic cell death (ICD) in the responder group, but not the non-responder group (**Figure S25B**)^52^. Consistent with these findings, we found a larger proportion of slow progressor patients had remodeled TMEs towards immune enriched environments compared to slow progressors (**Figure S25C**). In our serial scRNAseq data, the composition of the TME was altered after aPD1 with an increased proportion of T cells after chemotherapy treatment (**Figure 5A**). When comparing fast and slow progressors, we found that the expansion of T cells after aPD1 was predominantly in slow progressors, and was not observed in fast progressors (**Figure 5B**).

To understand which T cell subsets were most strongly affected by aPD1 treatment, we next subclustered the T and NK cells in our scRNAseq dataset to identify more granular subtypes (**Figure 5C and S26**). We found significant CD8 T cell expansion across timepoints (B, FU1, FU2) in slow progressor patients but not in fast progressor patients (**Figure 5D**), and this was consistent when comparing responders to non-responders (**Figure S27A**). We validated that there was increased CD8 T cell expansion in slow progressors (and treatment responders) using mIF for CD4, CD8, GzmB and PD-L1 (**Figure 5E, F and S28**). Among CD8 T cell subsets, we found that CD8 naive, memory and exhausted subsets were significantly expanded after aPD1 in slow progressors, but not in fast progressors (**Figure 5D and S27B**). CD8 effector subsets were expanded in both slow and fast progressors after aPD1 (**Figure 5D**); however, when performing the analysis comparing responder and non-responder patients, this only held true in the responder subgroup (**Figure S27B**). Of note, within the CD4 T cell compartment, we found that Tregs and Th1 cells were most altered in frequency across treatment timepoints (**Figure S27C,D**).

In our analysis of gene expression programs, we had identified a tumor-reactive T cell program, which included CXCL13 as a top weighted gene, that was only identified post-chemotherapy (pT18_FU1) and ICB (pT12_FU2) (**Figure 3A** and **6A**). We found that the usage of these programs was higher in T cells of slow progressors compared to fast progressors (**Figure 3A and S25**), consistent with literature reports that have identified CXCL13+ CD8 T cells as a tumor-reactive T cell subset^36, 37, 43^. We further validated that CXCL13+ was increased in single CD8 T cells across timepoints, in addition to other tumor-reactive (CD39, CD103) and co-stimulatory markers (4-1BB, GITR) (**Figure 6B**). To further investigate the evolution of co-varying hubs identified after chemotherapy treatment (**Figure 3B**), we next looked for co-varying gene programs across samples after immunotherapy treatment (**Figure 6C**). We identified 4 major hubs post-immunotherapy, and notably found that the CXCL13 T cell program co-varied with epithelial adhesion, ISG and proliferation programs, myeloid dendritic cell and IL1 programs, Treg and T cell cytotoxicity programs, and stromal proliferation programs (Hub 2I; **Figure 6C and S15**). We termed this the ‘immunity hub’ and note that we find that compared to chemotherapy treatment alone, the addition of immunotherapy led to the co-variation of the tumor-reactive CXCL13 program with other programs that have several similar features to those seen in other multicellular hubs identified in colorectal cancer (CRC)^9^. In particular, this includes the presence of epithelial ISG and Treg programs, but also with distinct properties from the hubs identified in CRC that include the presence of myeloid IL1 and dendritic cell programs.

**FIgure 6.**
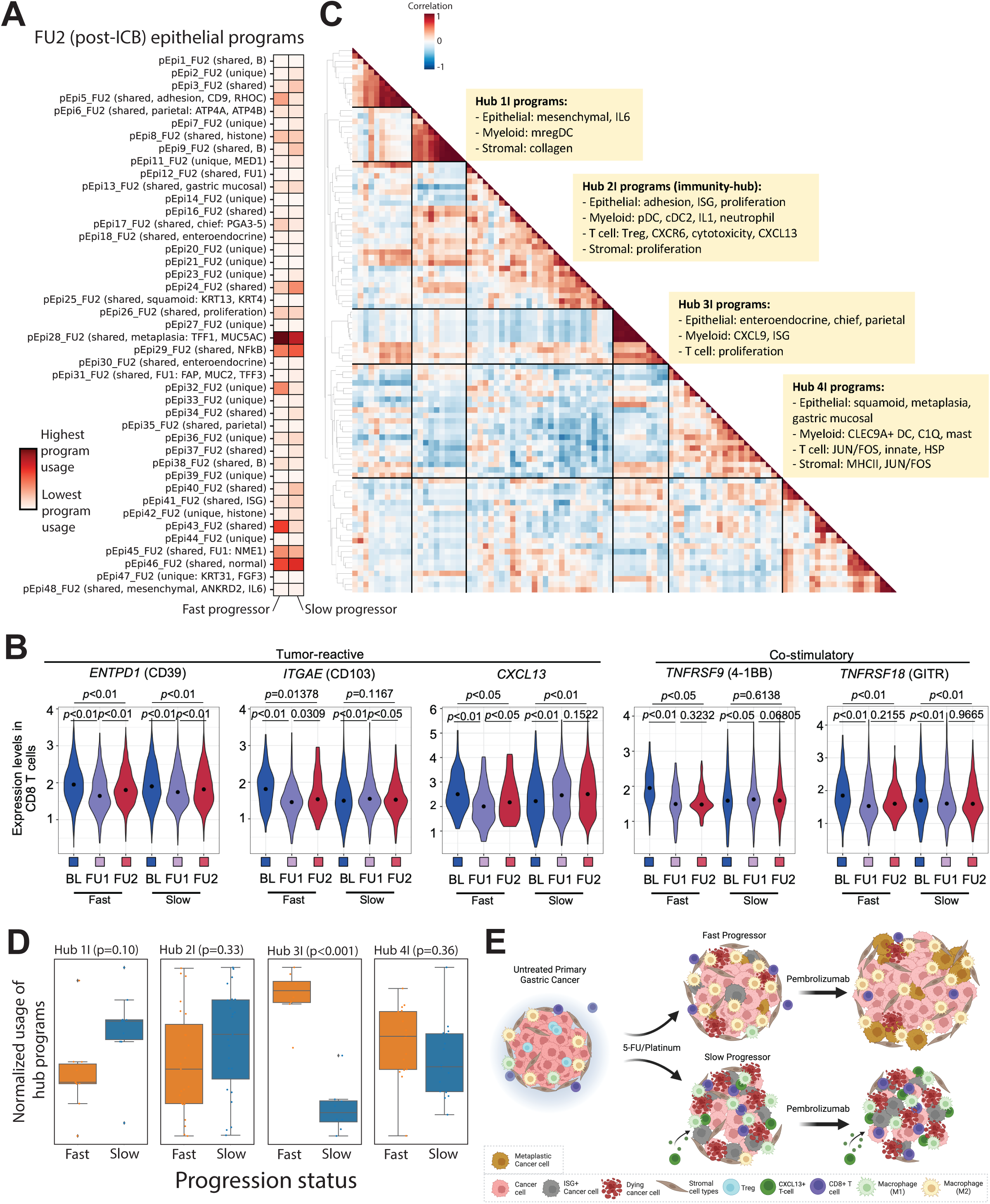
Multicellular hubs underlie chemoimmunotherapy resistance and response. (A) cNMF was performed on epithelial cells at FU2. Shown is the mean usage of each cNMF gene program in the epithelial cells of fast and slow progressing patients. (B) Expression of CD39, CD103, CXCL13, 4-1BB and GITR in single CD8 T cells segregated by timepoint (BL, FU1, FU2) and by fast versus slow progressing patients. Statistical comparison performed using a Wilcoxon signed-rank test. (C) Heatmap showing pairwise correlation of gene program activities across all patient samples at FU2 using the 90th percentile of patient-level program activity in epithelial, myeloid, T, NK and stromal cells. Hierarchical clustering was performed to identify clusters of co-varying proteins, which have been labeled as Hub1C to 5C. (D) Average z-scored usage of all gene programs in each hub split by fast and slow progressing patients. Statistical comparison performed using a two-sample t-test with Bonferroni correction. (E) Summary schematic of proposed changes to the TME after 1 cycle of chemotherapy and chemoimmunotherapy in fast versus slow progressing patients; in particular, fast progressing patients have induction of metaplasia programs and increased abundance of suppressive M2 macrophages. Slow progressing patients have increased infiltration of CXCL13+ CD8 T cells after chemotherapy, and increased tumor-intrinsic ISG induction and inflammatory M1 macrophage subsets.

Of note, we identified three other notable multicellular hubs in the aPD1 treated samples that include: 1) epithelial mesenchymal and IL6, mregDC and stromal collagen programs (Hub 1I); 2) epithelial neuroendocrine, chief and parietal, and myeloid CXCL9 and ISG programs (Hub 3I); and 3) epithelial squamous and metaplasia, myeloid C1Q macrophage and CLEC9+ DC, T cell JUN/FOS, innate and HSP, and stromal MHCII and JUN/FOS programs (Hub 4I). We next analyzed hub participation after aPD1 between slow and fast progressors, and we found that compared to fast progressors, slow progressors trended towards higher usage of programs in Hub 1I and Hub 2I, the immunity hub, and had significantly lower usage of 3I, both of which may therefore be features of aPD1 resistance (**Figure 6D**).

Despite being more highly represented in slow progressors, Hub 1I centered around several immune features known to directly antagonize immunotherapy response most notably mregDC^53^ and IL-6^54^. Notably, circulating plasma IL-6 was higher among fast progressors in our trial (**Figure S20**). We suspect this paradox may likely be due to the source of IL-6 in circulation and as reported in previous studies^55^. Whereas in Hub 1I we find epithelial cells as a major source of IL-6, several studies have reported that IL-6 produced by other cell types, particularly myeloid cells, may be poorly prognostic^55, 56^. Hub 4I contained immunosuppressive C1Q+ TAM^57^ programs covarying with markers known to be involved in T-cell exhaustion and impaired antitumor immunity (JUN/FOS)^58^. The epithelial programs covarying with the immunosuppressive immune cell programs may hint at epithelial cell plasticity and are enriched for squamoid and metaplasia (Hub 4I) programs, which are implicated in therapeutic resistance^59, 60^.

## Discussion

Efforts to identify determinants of response to cytotoxic chemotherapy and immune checkpoint blockade, and dissecting the evolution of immune cell populations and interactions under therapeutic pressures has been largely limited to comparisons of pre-treatment and temporally distant post-treatment and/or progression samples across solid tumors^10, 61, 62^. While useful, this framework limits the ability to develop early, true on-treatment biologic insights and biomarkers to inform biologically adapted clinical trial concepts and nominate therapeutic targets. Here we present a novel and large sequential sampling cohort in the framework of a prospective phase II chemoimmunotherapy trial in advanced gastric cancer. To our knowledge this is the first trial of its kind to couple serial true on-treatment biopsies with multiparametric molecular characterization in advanced GEA.

Taken together our data point toward an initial complex coordinated response to cytotoxic chemotherapy that differs between patients who go on to develop response/benefit and those that do not (**FIgure 6E**). Notably, the baseline TME composition appears to have a lesser impact than the early adaptive changes induced by chemotherapy. Using an initial window after 1 cycle of 5FU/platinum (∼2-3 weeks between baseline; BL, and follow up 1; FU1 samples) we uncover early on treatment patterns that converge on central components of the TME, specifically the myeloid and T-cell interactions with both the epithelial and stromal elements. We leveraged co-varying gene programs to infer multicellular interaction networks across our timepoints. This strategy has identified key conserved programs and “immune hubs” in colorectal cancer, but was limited to untreated earlier stage patient samples^9, 29^. How these networks evolve during therapeutic pressure has been a scientific blindspot. The described hubs with colocalization of IFNG+ and CXCL13+ T-cells programs is something we observed, but only after treatment with chemotherapy (FU1). Interestingly, Pelka and colleagues did not observe this pre-existing hub when restricting analysis to microsatellite stable (MSS) colorectal cancers^9, 63^. This may point to an early “priming” achieved after chemotherapy and underlie the limited activity of aPD1 monotherapy across GEA cancers, where the vast majority are MSS.

Conceptually, a single dose of chemotherapy looks to incite a turf war between opposing communities (hubs), the prevailing composition and usage of which will influence the relative contribution of PD1. This idea is reflected in the relative hub usage in fast and slow progressors with fast progressors demonstrating increase in epithelial metaplasia, myeloid proliferation and C1Q and TREM2 macrophage programs (Hub 2C). Predicted cellular interactions in this group are associated with known adaptive resistance mechanisms including tumoral plasticity and IL-6^64–66^. Conversely, we observe a portion of patients with suggestions of pro-immunity hubs evolving after chemo and furthered by the addition of aPD1 and it is tempting to speculate that patients who are able are indeed some of the patients who truly derive very durable benefit, often described in the “tail of the curve” in aPD1 clinical trials. We in fact see greater representations of the aforementioned hubs after chemotherapy and after aPD1 in patients that are slow to progress (have more durable responses) compared to those that progress faster. From a clinical perspective it remains unclear whether the optimal strategy is to attempt to restrain the initial formation of anti-immunity hubs with agents that deprive the TME of immunosuppressive signals as has been attempted with neutralizing antibodies to IL-6^54^ and DKK1^67^, or to try to directly stimulate the induction of pro-immunity hubs with agonist approaches like STING^68, 69^ or TLR7/8^70, 71^. Towards the later approach, we find that programs for plasmacytoid and conventional dendritic cell subsets were present in hubs that favored slow progression (e.g. Hub 2I).

Although our dataset lacks deep spatial characterization some of the broad features raised in our scRNAseq analyses are recapitulated in our multiplex IHC/IF analysis. To date some of the correlations between inferred network structure from scRNAseq and ground truth analysis such as IHC and IF and emerging spatial transcriptomics appear to largely hold, which is reassuring for our data^9, 10, 38, 62, 63, 72, 73^. As we needed to prioritize patient safety, our serial samples are limited to endoscopic biopsies of the primary tumor and we recognize the potential value for complementary spatial transcriptomics approaches. For instance, we nominate an increase in cDC1 programs that parallels CD8 effector increase and M2 decrease only in responders. Receptor-ligand predictions point to a direct communication among these cell types but we cannot confirm the spatial orientation. In a recent analysis from a PD-1 containing CRC trial (Keynote-177) the spatial proximity between CD74+ macrophages and PD1+ CD8 T-cells was a robust response predictor^74^.

Tumor associated macrophages (TAMs), particularly the alternatively activated (M2-like) subset are known to facilitate tumor promoting processes including angiogenesis, impede antigen presentation and suppress the tumoricidal functions of CD8 T-cells and NK cells^75–78^. Recent data suggest SPP1-high TAMs are linked to EMT, higher angiogenesis scores and liver metastasis^42, 79, 80^. We observed a decrease in SPP1 expression only among responding patients, and a suggestion of increased macrophage-derived MIF inhibiting T-cell proliferation when examining M2 macrophage interactions with T-cells. MIF is known to directly antagonize immune responses in melanoma^81^. Despite preclinical rationale for repolarizing M2-like TAMs toward the more anti-tumor M1-like state, the clinical development has fallen short in several chemotherapy free approaches, including in GEA (reviewed in^39^).

While the deep sampling in our dataset is a unique strength, we recognize that broad implementation of short interval serial biopsy is unlikely to be clinically feasible. While circulating tumor DNA is a validated prognostic and predictive marker across multiple cancers it is inherently biased toward examining tumor intrinsic genomic features and general tumor burden^82–87^. As our focus was primarily on immune cell adaptations we paired a broad (∼3,000 analytes) proteomic panel to our tissue sampling in an effort to explore peripheral surrogates of tissue level changes. We noted protein modules that generally correlated with scRNAseq features and moved differentially among fast and slow progressors. In examining the major protein constituents of the representative modules we observed multiple presumed tumor-derived and immunosuppressive cytokines increased in fast progressors suggesting our proteomic data reflects a conserved biology. While it remains unclear whether the source of these proteins in the plasma are from the tumor or normal stroma elsewhere in the body, these data lay the groundwork for further exploration towards proteomic biomarkers that may be used to monitor cellular adaptations to treatment. Together, our analysis highlights the exploratory utility in integrating plasma proteomic datasets with tumor tissue data to learn composite modules of proteins that may underlie therapy relevant TME changes. The concept of composite markers to better predict outcomes is recently supported by a large analysis in immunotherapy outcomes, lending additional support to our findings^88^.

The inciting signals, or mix thereof, that drives the initial formation of pro and anti-immunity hubs is an area of important ongoing investigation. Coupling WTS and scRNAseq can reconstruct the immune TME and provide prognostic clinical information, but it does not confirm the functional impact of predicted relationships. Pre-clinical models to functionalize the study of immune TME manipulation remains a problem in immunomodulatory drug development, including GEA^89–91^. We had intentionally structured our trial with serial timepoints to partly address this limitation and enhance confidence in our observations and we hope the future work with improved models can test some of our observations^92^. In addition, the focus of our work is on the immune interaction networks and detailed analyses incorporating tumor genetics, neoantigen prediction, and T cell receptor sequences are beyond the scope of this manuscript but remain areas of interest. In fact, overall we envision our dataset to serve as an in-silico framework to directly support and evaluate observations from investigators exploring preclinical models in GEA and hope our data will be a valuable resource for the field.

## Data Availability

Sequencing data from the patient cohort will be deposited in the European Nucleotide Archive (ENA) and will be made available at the time of publication.

## Acknowledgements

Dana Farber Cancer Institute / Harvard CancerCare GI SPORE Career Enhancement Award (AM), Sky Foundation Pancreatic Cancer Research Grant (AM), Doris Duke Charitable Foundation Physician Scientist Fellowship (AM), the DeGregorio Family Foundation (SJK), SU2C Gastric Cancer Interception Research Team Grant (Grant Number: SU2C-AACR-DT-30-20) Award (HL, SJK, JL). This research grant is administered by the American Association for Cancer Research, the Scientific Partner of SU2C. This research was supported by a grant of the Korea Health Technology R&D Project through the Korea Health Industry Development Institute (KHIDI), funded by the Ministry of Health & Welfare, Republic of Korea (grant number: HR20C0025 [SK]). This paper was supported by SKKU Excellence in Research Award Research Fund, Sungkyungkwan University, 2022 (JL).

## Methods

### Clinical Trial

All patients were enrolled in this prospective open-label phase II trial (ClinicalTrial.gov identifier : NCT04249739). Eligible patients were required to meet the following criteria: (i) at least 19 years old, (ii) histologically confirmed diagnosis of unresectable, metastatic gastric cancer, (iii) adequate organ function per protocol, and (iv) Eastern Cooperative Oncology Group performance status of 0 or 1. All patients were naïve to prior chemotherapy. The trial protocol was approved by the Institutional Review Board (IRB) of Samsung Medical Center (Seoul, Korea; IRB No. 2019-11-089) and was conducted in accordance with the Declaration of Helsinki and the Guidelines for Good Clinical Practice. All patients provided written informed consent before enrollment. If the tumor was HER2 negative, the patient was enrolled on to capecitabine/oxaliplatin/pembrolizumab. If the tumor was HER2 positive, the patient received capecitabine/cisplatin/trastuzumab and pembrolizumab. The trial was registered at clinicaltrials.gov.

### Tumor sample and peripheral blood collection

All primary tissues were collected from the patients who had biopsies at day 1 before cycle 1, day 1 before cycle 2 and day 1 before cycle 7. Matched PB was collected prior to initiation of treatment. Tumor tissues were obtained from endoscopic site-mapping biopsies. If tumor purity was estimated to be more than 40% after pathological reviews, tumor DNA and RNA were extracted by using a QIAamp Mini Kit (Qiagen, Hilden, Germany) according to the manufactures’ instructions for exome and transcriptome sequencing. Concentration, 260/280 and 260/230nm ratios were measured with ND1000 spectrophotometer (Nanodrop Technologies, Thermo-Fisher Scientific) and then DNA/RNA was quantified using a Qubit fluorometer (Life Technologies).

### PD-L1 IHC

Tissues were freshly cut into 4-μm sections, mounted on Fisherbrand Superfrost Plus Microscope Slides (Thermo Fisher Scientific), and then dried at 60°C for 1 hour. IHC staining was carried out on a Dako Autostainer Link 48 system (Agilent Technologies) using the Dako PD-L1 IHC 22C3 pharmDx Kit (Agilent Technologies) with the EnVision FLEX Visualization System and counterstained with hematoxylin according to the manufacturer’s instructions. PD-L1 protein expression was determined using CPS, which was the number of PD-L1–stained cells (tumor cells, lymphocytes, and macrophages) divided by the total number of viable tumor cells and multiplied by 100. The specimen was considered to have PD-L1 expression if CPS ≥ 1.

### MSI status

Tumor tissue MSI status was determined using PCR analysis of five markers with mononucleotide repeats (BAT-25, BAT-26, NR-21, NR-24, and NR-27). Briefly, each sense primer was end-labeled with FAM, HEX, or NED. Pentaplex PCR was performed, and the PCR products were run on an Applied Biosystems PRISM 3130 automated genetic analyzer. Allelic sizes were estimated using Genescan 2.1 software (Applied Biosystems). Samples with allelic size variations in more than two microsatellites were considered MSI-H.

### Sequencing of Whole Exome and Whole Transcriptome

Sequencing was performed using genomic DNA (gDNA) from the tumor tissues and matched blood samples using a QIAamp DNA Blood kit (QIAGEN). For construction of standard exome capture libraries, we used the Agilent SureSelect Target Enrichment protocol for an Illumina paired-end sequencing library together with 1 μg of inputted gDNA. In all samples, the SureSelect Human All Exon V6 probe set was used. We assessed the quantity and quality of DNA by PicoGreen and agarose gel electrophoresis. We diluted 1 μg of gDNA in the elution buffer and sheared it to a target peak size of 150 to 200 bp using the Covaris LE220 focused ultrasonicator device (Covaris Inc.) according to the manufacturer’s recommendations. The fragmented DNA was repaired, and “A” was ligated to the 3′-end. Then, we ligated the fragments with Agilent adapters and amplified them using PCR. The prepared libraries were quantified using the TapeStation DNA ScreenTape D1000 (Agilent). For exome capture, 250 ng of DNA library was mixed with hybridization buffer, blocking mixes, RNase block, and 5 μg of SureSelect all exon capture library, according to the standard Agilent SureSelect Target Enrichment protocol. Hybridization to the capture baits was conducted at 65°C using a heated thermal cycler lid option at 105°C for 24 hours on the PCR machine. The captured DNA was washed and amplified. The final purified product was quantified by qPCR according to the qPCR Quantification Protocol Guide (KAPA Library Quantification kits for Illumina Sequencing platforms) and qualified using the TapeStation DNA ScreenTape D1000 (Agilent). Samples were multiplexed, and flow-cell clusters were created using the TruSeq Rapid Cluster kit and the TruSeq Rapid SBS kit (Illumina). Indexed libraries were submitted to an Illumina HiSeq2500 (Illumina), and paired-end (2 × 100 bp) sequencing was performed.

### scRNA-seq

For single-cell preparation, tumor tissue was dissociated with the gentleMACS Dissociator and Tumor infiltrating lymphocyte Kit (Miltenyi Biotec) according to the manufacturer’s protocol. The cells were then cryopreserved in liquid nitrogen until use. All samples showed a viability of around 90% on average after thawing. We performed 5′ gene expression profiling on the same single-cell suspension using the Chromium Single Cell V(D)J Solution from 10x Genomics according to the manufacturer’s instructions. Up to 8,000 cells were loaded onto a 10x Genomics cartridge for each sample. Cell-barcoded 5′ gene expression Libraries were constructed and sequenced at a depth of approximately 50,000 reads per cell using the NovaSeq 6000 platform (Illumina).

### WES Analysis

*Somatic variant calling*: WES reads were aligned to the reference human genome GRCh37 using BWA-MEM^93^ followed by preprocessing steps, including duplicate marking, indel realignment, and base recalibration using the Genome Analysis Toolkit (GATK; version 4.1.1.0)^94^, generating analysis-ready BAM files. To increase the sensitivity for identifying both the lower and higher allele frequencies of somatic variants in the given tumor and paired normal BAM files at the genomic locus, we used the union variant callsets from two tools: MuTect2^95^. Default parameters were applied, and both variant callers were run with dbSNP (version 138)^96^, 1000G (phase I)^97^ and HapMap (phase III)^98^ data for known polymorphic sites. Filtered variants with minimum depth ≥ 5 and minimum alternative alleles ≥ 2 were annotated using the Ensembl Variant Effect Predictor (VEP; release version 87)^99^ with the GRCh37 database.

#### Mutational Signature Analysis

Mutational signature analysis was performed using the deconstructSigs package (version 1.6.0) in R (PMID: 26899170). Exome regions were defined by the Agilent Sureselect V5 target region. Only somatic mutations in exome regions were considered, and trinucleotide counts were normalized by the number of times each trinucleotide context was observed in the exome region. Mutational signatures were represented by the following terms: age (SBS1 and SBS5), apolipoprotein B mRNA editing enzyme, catalytic polypeptide-like (APOBEC; SBS2 and SBS13), UV (SBS7a, SBS7b, SBS7c, and SBS7d), smoking (SBS4), homologous recombination deficiency (HRD; SBS3), mismatch repair deficiency (MMRD; SBS6, SBS15, SBS20, and SBS26), nucleotide excision repair deficiency (NERD; SBS8), DNA proofreading deficiency (DPD; SBS10a and SBS10b), and base excision repair deficiency (BERD; SBS18).

### Whole-Transcriptome Sequencing Analysis

We annotated RNA sequence reads with ENSEMBL (version 98) and aligned them to the human reference genome (GRCh38) using STAR (version 2.6.1)^100^. We quantified in units of transcript per million (TPM) as a function of gene expression using RSEM (version 1.3.1)^101^, applying the option parameters suggested by the GTEx project. TPM values less than one were considered unreliable and substituted with zero. By integrating the transcriptomic data, we classified each tumor sample into two microenvironment subtypes (immune-depleted, immune-enriched) using the molecular functional portrait^27^. The GSVA algorithm was used to validate our previous findings in this cohort^102^.

### Single cell RNA Sequencing Analysis

#### Preprocessing and annotations

We aligned scRNA-seq reads to the GRCh38 human genome reference and quantified them using a Cellranger (version 5.0). The data from all samples were combined in R v.4.1 using the Seurat package v.4.0^103^. We filtered out doublets using Scrublet^104^. In addition, cells with low-quality libraries (<400 genes) and high mitochondrial read proportion (>30%) were filtered out. Data from each sample were normalized, scaled, and subjected to principal component analysis, followed by a batch correction using the Harmony^105^. We used the UMAP algorithm to reduce the dimension for visual representation, and identified cell clusters using a shared nearest neighbor modularity optimization–based clustering algorithm. We identified various cell type clusters using the “FindAllMarkers” function in Seurat for each cluster and annotated them based on the expression of representative lineage markers. Gene enrichment related to phagocytosis, angiogenesis and PDGF pathway was estimated using the “AddModuleScore” function in Seurat.

#### InferCNV

inferCNV (https://github.com/broadinstitute/infercnv) was run on the unprocessed epithelial cells from each scRNA-seq tumor sample individually to infer copy number variations. When available, a matching normal sample was used as the reference. For the tumor samples without a matching normal sample, a standard reference was generated consisting of a random sampling of 20,000 cells from the combined population of epithelial cells from all normal samples, stratified by patient. The inputs include the combined count matrices of both the sample of interest and reference formatted as an R dgCMatrix, group annotations per cell barcode, and a list of the groups to use as reference. A gene ordering file was generated from the standard hg38 genome reference using the script provided in the above github repo. The i6 HMM implementation of inferCNV was used with a cutoff of 0.1 (as advised for 10x data), the subclusters analysis mode and median filtering enabled. This tool was run on Terra (https://app.terra.bio) via a workflow available here: https://dockstore.org/workflows/github.com/MilanParikh/infercnv_workflow.

#### Numbat

Numbat (https://github.com/kharchenkolab/numbat/) was run on each full tumor sample (all cell types) individually to call copy number variations and differentiate normal and tumor cells. When available a matching normal sample was used as the reference. For the tumor samples without a matching normal sample, a standard reference was generated consisting of a random sampling of 25% of all cells from normal samples, about 26,000 cells. The inputs include the bam file, bam index file, 10x barcodes file, and the raw count matrix as an R dgCMatrix for the sample of interest, as well as the cell type annotations and count matrix as an R dgCMatrix for the reference. All default parameters were used, including the provided SNP VCF and phasing panel from the 1000 Genome project. This tool was run on Terra (https://app.terra.bio) via a workflow available here: https://dockstore.org/workflows/github.com/MilanParikh/numbat_workflow.

#### Consensus NMF (cNMF)

cNMF (https://github.com/dylkot/cNMF) was run on each cell type at each timepoint separately for tumor samples or for each cell type separately in the normal samples to derive gene programs. For each of these runs, the input was an unprocessed AnnData file subset to the population of interest. The custom parameters used include the number of highest variance genes to use in the factorization step (2000) and the range of k values tested for by cNMF (3 to 101 with intervals of 3). All other default parameters were used. Once the prepare, factorization, and combine steps were completed, the generated k selection plot (plotting stability or error vs k values) was used to manually select the most appropriate k value, and then run the consensus step.To filter overlapping programs between timepoints or normal/malignant samples within the same cell type, a hypergeometric test was performed between the top 100 genes of each pair of programs. The Bonferroni corrected p-value was calculated and program pairs with a p-value of less than or equal to 0.05 were counted as overlapping. This tool was run on Terra (https://app.terra.bio) via a workflow available here: https://portal.firecloud.org/?return=terra#methods/mparikh/cnmf_parallel/7. The cNMF programs were subsequently manually annotated by running GSEA on the top 100 weighted genes for each program using the Enrichr implementation in GSEApy (https://gseapy.readthedocs.io/en/latest/introduction.html), by comparing program correlation in the single-cell data with other known program signatures, and by manually examining the top weighted genes in each program.

#### Co-varying programs

To find sets of co-varying gene programs, or hubs, a patient by program matrix was generated for each timepoint where the values were the 90th percentile of the cNMF program score for that program and patient. This was repeated for the 25th, 50th, 75th, and 95th percentiles as well. For each of these matrices a pairwise correlation of columns was calculated using pandas’ DataFrame.corr() method, resulting in a program by program correlation matrix. This was then visualized using Seaborn’s clustermap function with Euclidean distance as the spatial distance metric. The 90th percentile data is displayed in Figures 3 and 6, and was used to identify groups of co-varying programs.

To find sets of co-varying gene programs, or hubs, a patient by program matrix was generated for each timepoint where the values were the 90th percentile of the cNMF program score for that program and patient. The Pearson correlation coefficient, R, was calculated for each pair of programs using pandas’ Dataframe.corr() method. To calculate significance, the patient assignments for each cell were randomly shuffled using pandas’ Series.sample() method and then the 90th percentile scores and correlation coefficients were recalculated. This was repeated for a 1000 iterations to generate a null distribution of correlations for each pair of programs. A p-value was then calculated by counting how many times the randomly shuffled patient assignment correlations were higher than the true correlation, divided by the total number of iterations.

*Ligand-receptor analysis*: CellphoneDB’s (https://github.com/ventolab/CellphoneDB) statistical analysis method was run on each timepoint and normal cells separately to infer ligand-receptor pairs. The inputs include an Anndata file (.h5ad) for each timepoint and cell type annotations provided as a two column csv with cell barcodes and their corresponding cell type. The default parameters were used including 1000 iterations, a p-value of 0.05, and a threshold of 0.01, with no subsetting. This tool was run on Terra (https://app.terra.bio) via a workflow available here: https://dockstore.org/workflows/github.com/MilanParikh/cellphonedb_workflow.

### Olink plasma proteomics

#### Sample collection

Peripheral blood samples (volume = 10 mL) were collected at protocol specified timepoints in EDTA tubes and plasma was separated by centrifuging for 10 min at 1000 rpm twice. Plasma was stored at -80 degrees Celsius and plasma aliquots were prepared for Olink according to manufacturer guidance (https://olink.com/). Plasma aliquots were processed using the Olink proteomics platform as previously described<u>(Filbin et al. 2021)</u>.

#### Sample pre-processing

The R (version 4.0.3) programming environment was used for analysis of all plasma proteomic data unless otherwise specified. A data frame containing plasma proteomic data in the form of normalized protein expression (NPX) values was loaded into R using the read_NPX function in the OlinkAnalyze package (https://github.com/Olink-Proteomics/OlinkRPackage). Dimensionality reduction using principal components analysis (PCA) was performed using the prcomp function in the prcomp package. Principal components (PCs) were visualized using the autoplot function in the prcomp package. A Uniform Manifold Approximation and Projection (UMAP) embedding was generated for visualization using the umap function in the umap package. A custom built plotting function was used to visualize UMAP embeddings (https://github.com/arnav-mehta/covid19-proteomics/blob/main/umap_plot.R). All heatmaps in the paper were generated using the heatmap.2 or heatmap3 functions.

#### Linear mixed-effect model (LMM)

LMMs were fit to each Olink assay using the lme4 package. Each LMM included a main effect of time, a main effect of progression status (fast or slow), the interaction between time and progression status, and the patient ID as a random effect to account for the fact that measurements coming from the same individual are not independent. Significance testing of the three model terms was performed using an F-test using Satterthwaite degrees of freedom and implemented with the lmerTest package. P-values were adjusted to control the false discovery rate (FDR) at 5% using the Benjamini-Hochberg method. Estimated differences in NPX means between fast and slow progressors were calculated from the LMMs for each assay using the emmeans package. P-values for estimates from each assay were adjusted using the Tukey method. Results from this analysis are in **Table S6**. Point range plots for each protein were visualized using the ggplot2 package.

#### Correlation of plasma proteins with bulk RNA-seq gene expression and TME phenotypes

To calculate correlations between plasma protein abundance and gene expression in tumors across all sample collections, we calculated the correlation across all samples of NPX values in the plasma and TPM values from bulk RNA-sequencing data of the tumor for each gene using all sets of matched peripheral blood and biopsy collections. Correlations were computed using the corr function in Python (Python 3.8) and visualized using the distplot function in the Seaborn package in Python. To look at the correlation between plasma proteins and different TME phenotypes, we calculated the correlation across all samples of NPX values in the plasma and GSVA scores calculated from bulk RNA-seq data for curated gene sets representative of different TME phenotypes. Correlations were calculated using the corr function in Python as above, and the correlation for each protein / gene combination with TME phenotype was visualized using the scatterplot function in the Seaborn package in Python.

#### Mapping proteins to single-cell gene expression

For each significant protein identified in our LMMs, we next visualized the gene expression of each protein in each high-level cell subset in our scRNAseq data at each timepoint. We pseudobulked the total gene expression counts for each cell subset at each timepoint, and used the heatmap function in the Seaborn package in Python to visualize to visualize this expression.

### Statistical analyses

All statistical analyses were conducted using R V.4.1.1 (The R Foundation for Statistical Computing, www.R-project.org) or as specifically specified for each method used. All variables were compared using Wilcoxon signed-rank test and paired Wilcoxon signed-rank test. All p-values were two-sided, and results were determined to be significant at p < 0.05.

### Imaging mass cytometry analysis with 17 cell markers

5-µm-thick tissue sections were made from formalin-fixed paraffin-embedded tissue blocks. Tissue sections were mounted on glass slides and were prepared according to the manufacturer’s instructions. Tissue sections were baked at 60°C for 2 hour and deparaffinized using Neo-Clear (Sigma Aldrich, St. Louis, MO, USA) and rehydrated in descending grades of ethanol (100%, 95%, 80%, 70% of ethanol) for 5 minutes each. Antigen retrieval was performed under 96°C for 30 minutes (IHC-Tek^TM^ , IHC WORLD, USA). After that, tissues were cooled down to 70°C and washed using deionized water. Before staining, tissues were blocked using 3% BSA for 45 minutes at room temperature in the hydration chamber. The tissues were incubated overnight with antibody cocktails including 17 antibodies (Extended Data table 1) at 4°C in the hydration chamber. After staining, tissues were washed in 0.2% Triton X-100 (Sigma Aldrich, St. Louis, MO, USA) dissolved in PBS two times for 8 minutes. Tissues were also stained in Ir-intercalator solution which was diluted 1:400 in PBS to detect nuclei. Stained tissues were analyzed using the Imaging Mass Cytometry (IMC), Hyperion (Standard BioTools Inc, South San Francisco, CA, USA) after tuning the instrument according to manufacturer’s instructions. 700 µm x 700 µm regions were selected and analyzed for further analysis. IMC image data were acquired at a laser frequency of 50 Hz. Raw image data were preprocessed using MCD Viewer (Standard BioTools Inc, South San Francisco, CA, USA).

Cell segmentation was performed using Highplex FL algorithms in HALO v3.4 (Indica Labs) based on nuclei staining (Ir-intercalator solution). After classification, cells were classified into two groups, tumor and stromal using a random forest model. We also annotated macrophage M1 and M2, CD8 T cells, CD4 T cells based on signals of each immune cell marker. First, tumor cells were annotated based on the signal of pankeratin (panCK) and these cells were excluded for further cell annotation. CD14 positive and CD 68 positive cells were annotated to M1 macrophages derived from monocytes and CD68 positive CD163 negative cells were annotated to M1 macrophages. M2 macrophages were annotated with CD163 positive and CD68 positive cells. CD8 positive cells were annotated to CD8 T cells and CD4 positive cells were annotated to CD4 T cells. All cell segmentation was performed using Highplex FL algorithms. After cell segmentation, the number of each cell was acquired from HALO software and multiplex images of each tissue were visualized in HALO software. All statistical analysis was performed using R software.

## Supplemental Figure Legends

**Figure S1.**
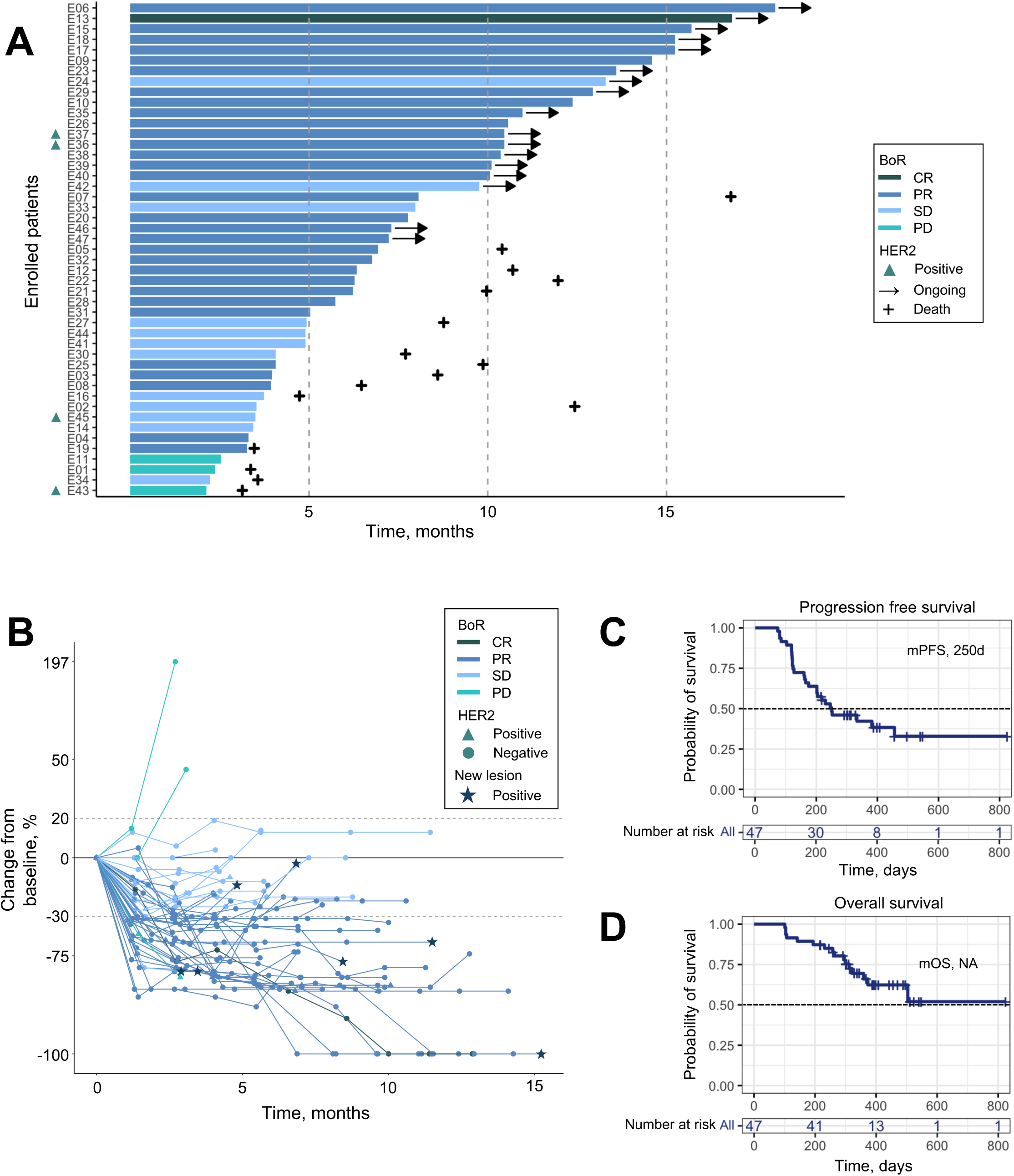
(A) Swimmers plot of patient response and duration in the described clinical trial. (B) Scatter plot showing temporal course of tumor response measured by RECIST for all patients in the reported clinical trial. (C) Progression free survival (PFS) and (D) overall survival (OS) for all patients in the reported clinical trial.

**Figure S2.**
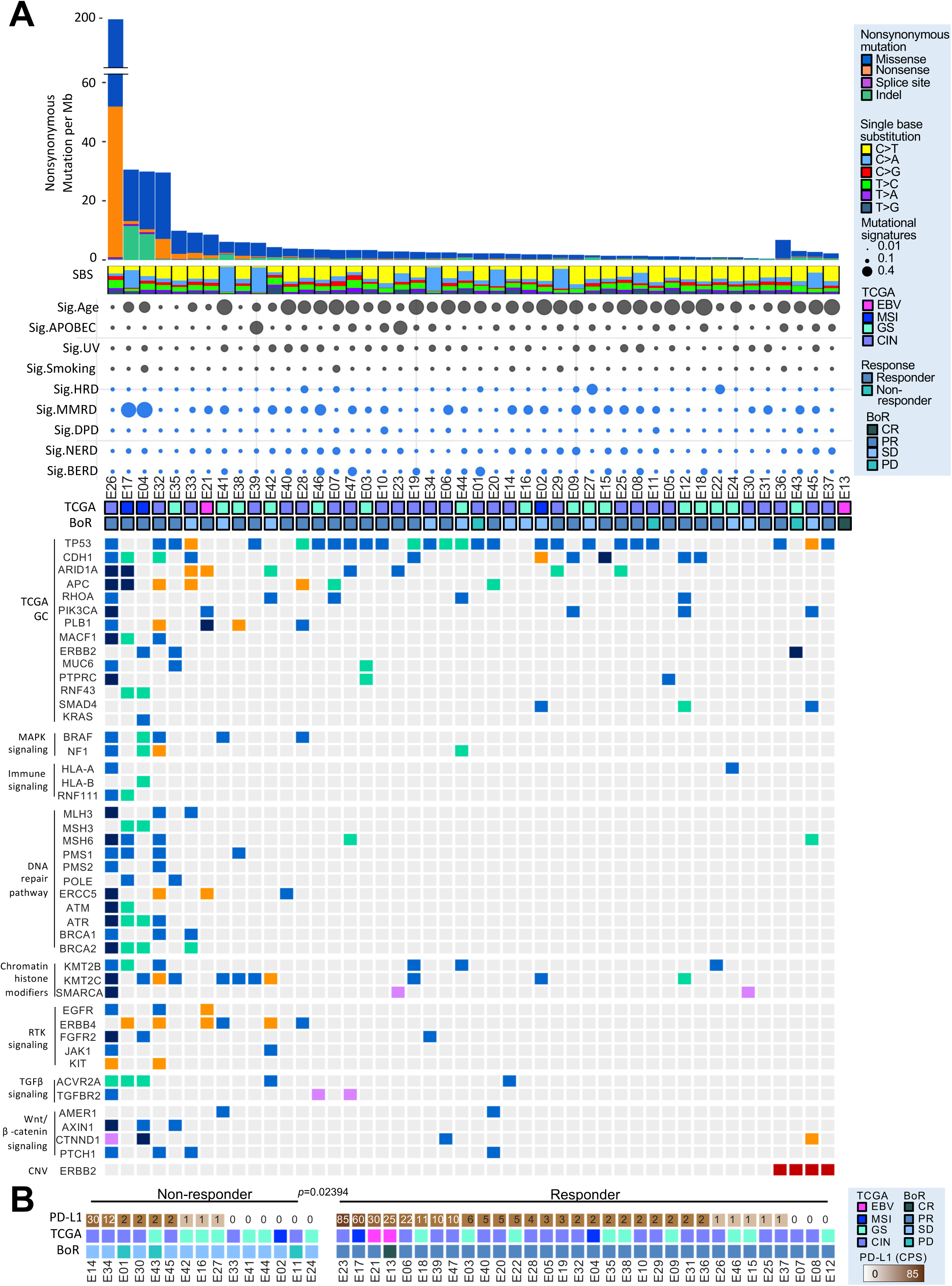
(A) CoMut plot demonstrating TCGA subtype, tumor mutational burden, mutational signature enrichment and mutational profiles of key driver genes for all patient samples at baseline in the reported clinical trial. (B) PD-L1 expression for each patient sample at baseline in the reported clinical trial.

**Figure S3.**
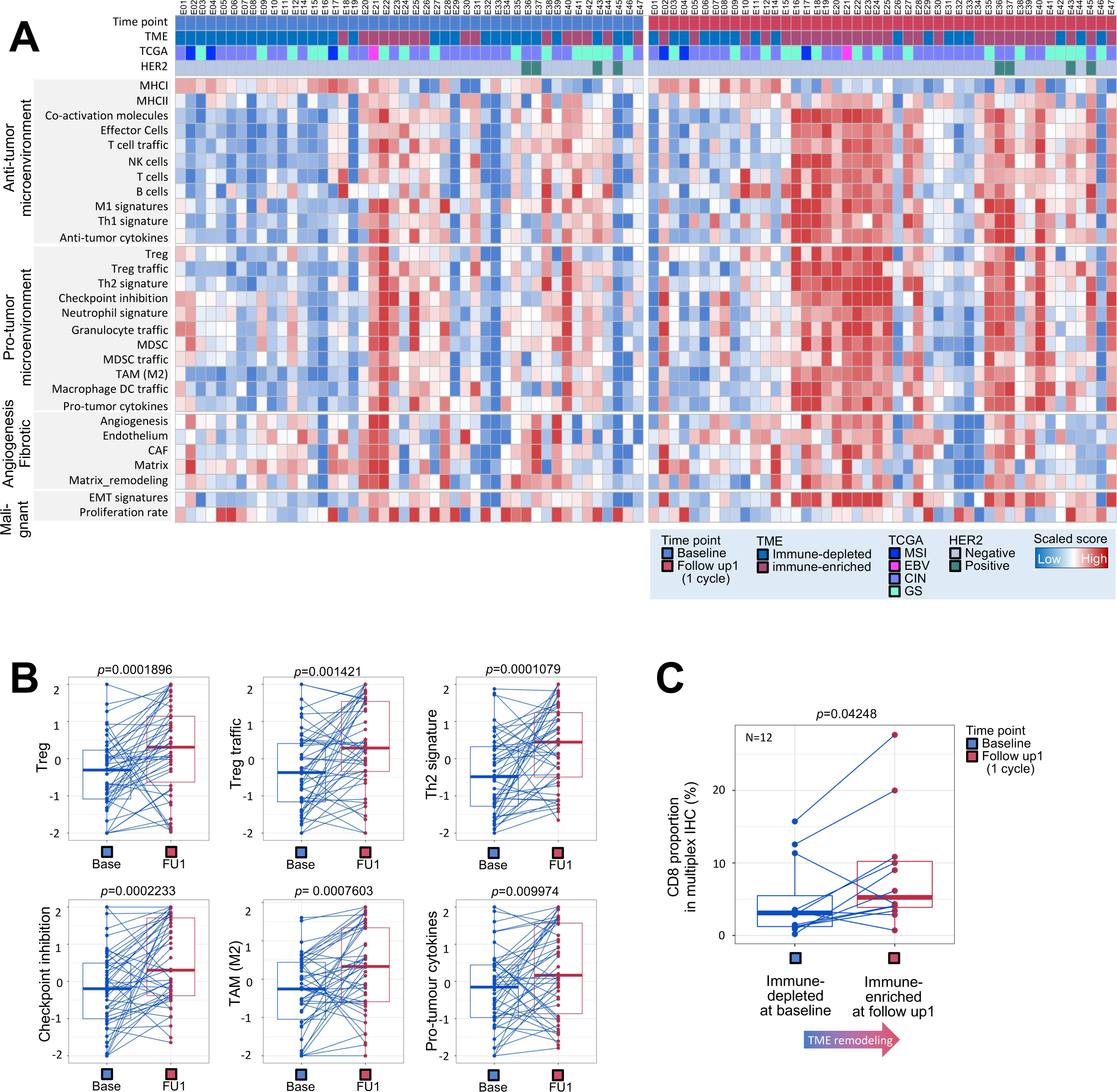
(A) Enrichment of gene expression signatures for TME subtypes using bulk RNA-sequencing data for all patient samples at baseline and after chemotherapy (FU1). (B) Plots demonstrating change in immune related gene signatures (Treg, Treg trafficking, Th2, immune checkpoints, M2 macrophages, pro-tumorgenic cytokines) after chemotherapy treatment. Lines connect samples obtained from the same patient. Statistical comparison performed using a Wilcoxon signed-rank test. (C) Quantification of CD8+ cell abundance using immunohistochemistry (IHC) at baseline and after chemotherapy (FU1) in 12 patient samples. Statistical comparison performed using a Wilcoxon signed-rank test.

**Figure S4.**
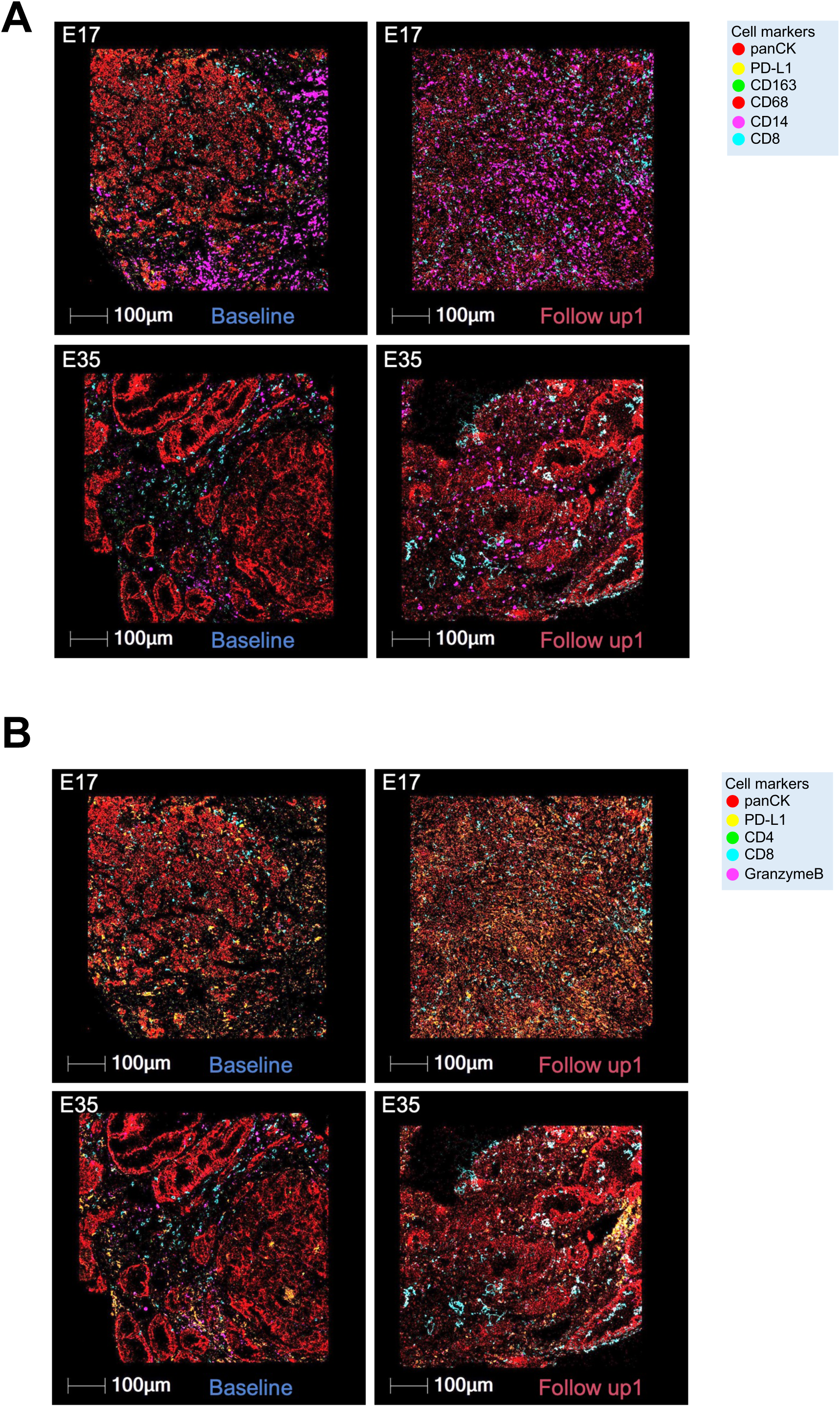
Multiplexed immunofluorescence (mIF) images of BL and FU1 samples from two patients, E17 (slow progressor) and E35 (fast progressor), staining for (A) panCK, PD-L1, CD163, CD68, CD14 and CD8, and (B) panCK, PD-L1, CD4, CD8 and Granzyme B.

**Figure S5.**
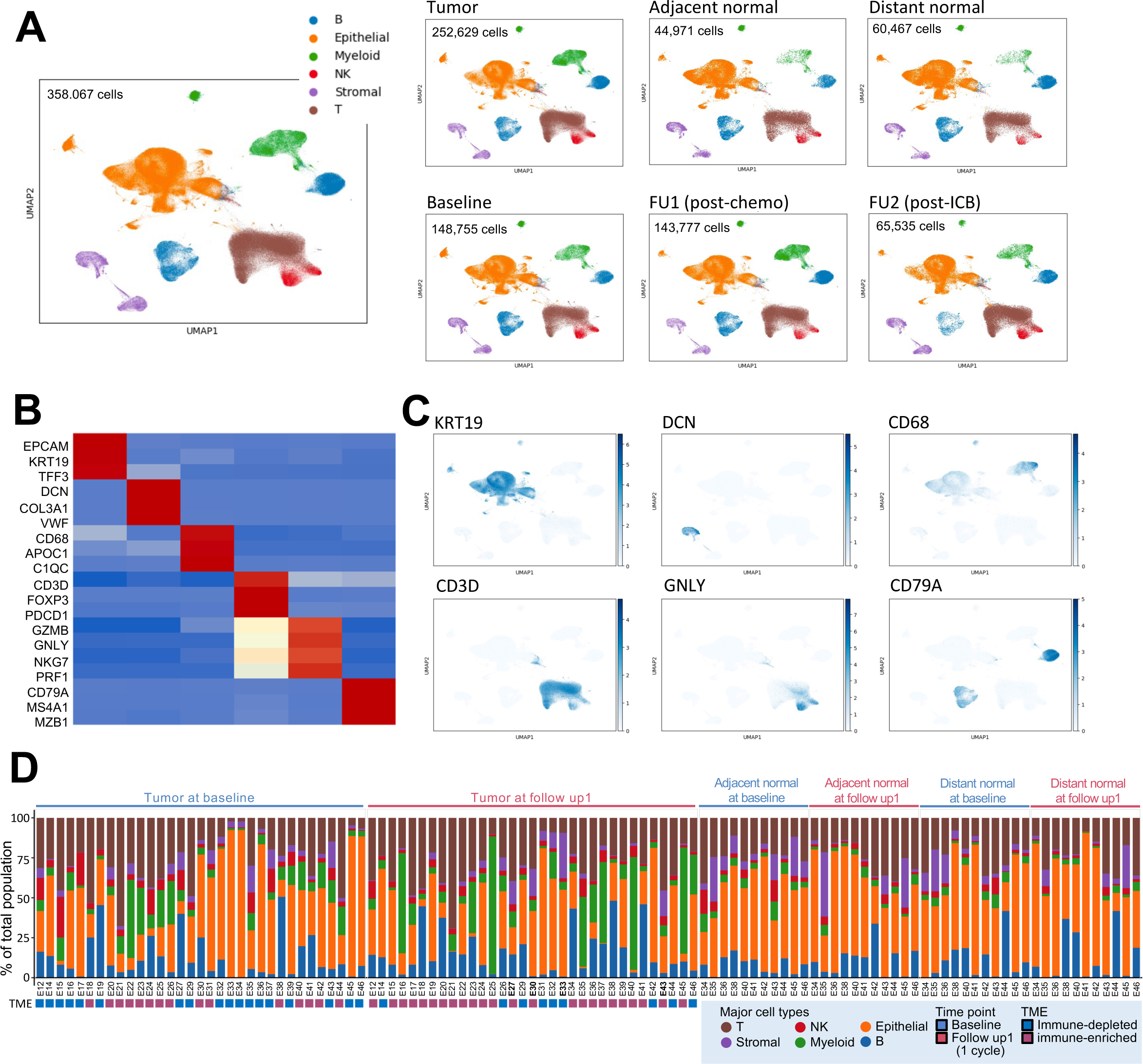
(A) UMAP embedding of single cell transcriptomes obtained from all samples in this trial. Labeled are canonical cell types separated by tumor v.s. normal tissue, and by timepoint. (B) Heatmap and (C) UMAP embeddings showing marker gene expression for broad cell types in (A). (D) Broad cell type composition per sample across tumor and normal tissue.

**Figure S6.**
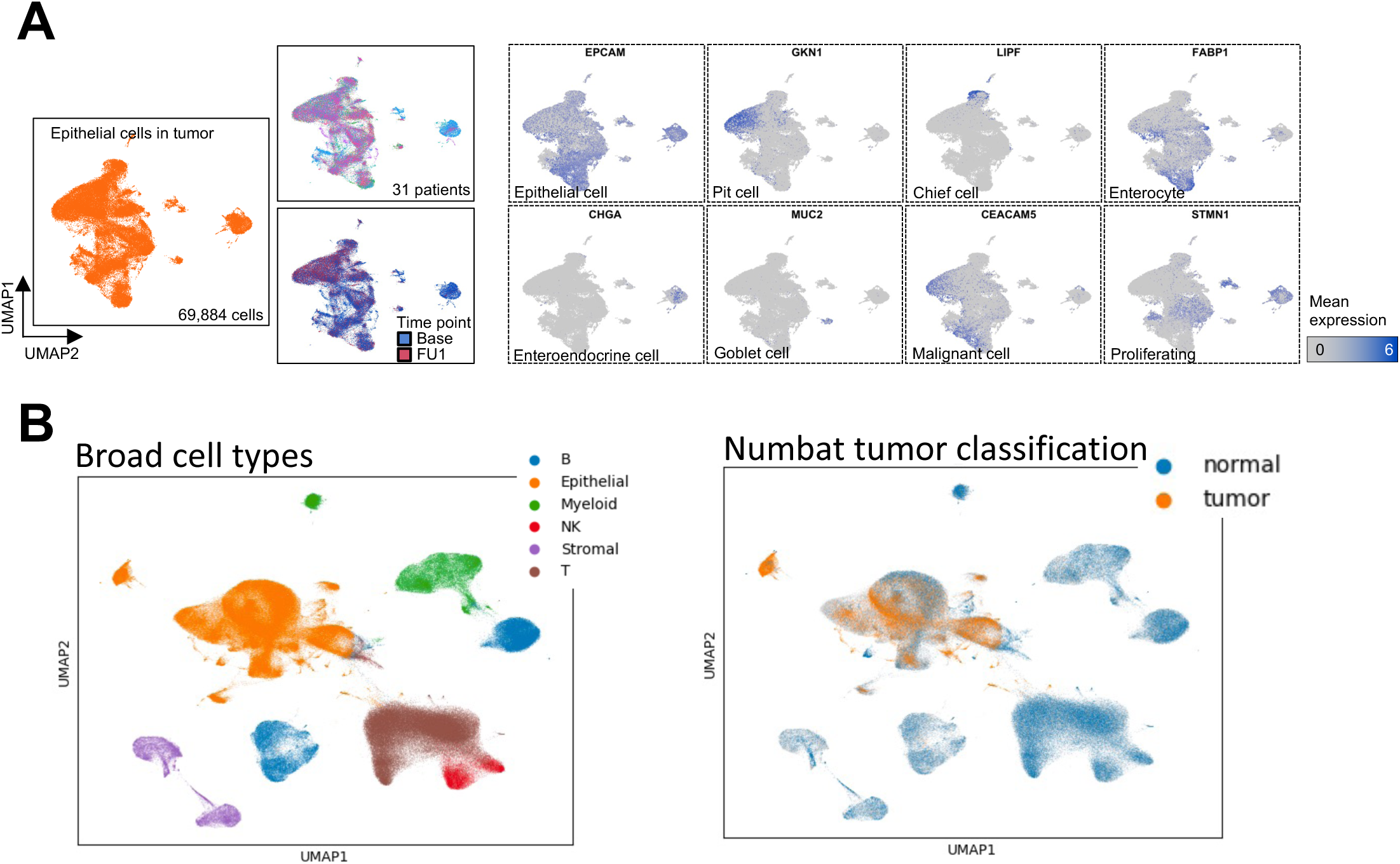
(A) UMAP embedding of single cell transcriptomes from epithelial cells obtained from all samples in this trial. Shown are UMAP embeddings with expression of key epithelial genes. (B) UMAP embeddings color coded by predicted annotation of tumor cell v.s. normal cell using Numbat.

**Figure S7.**
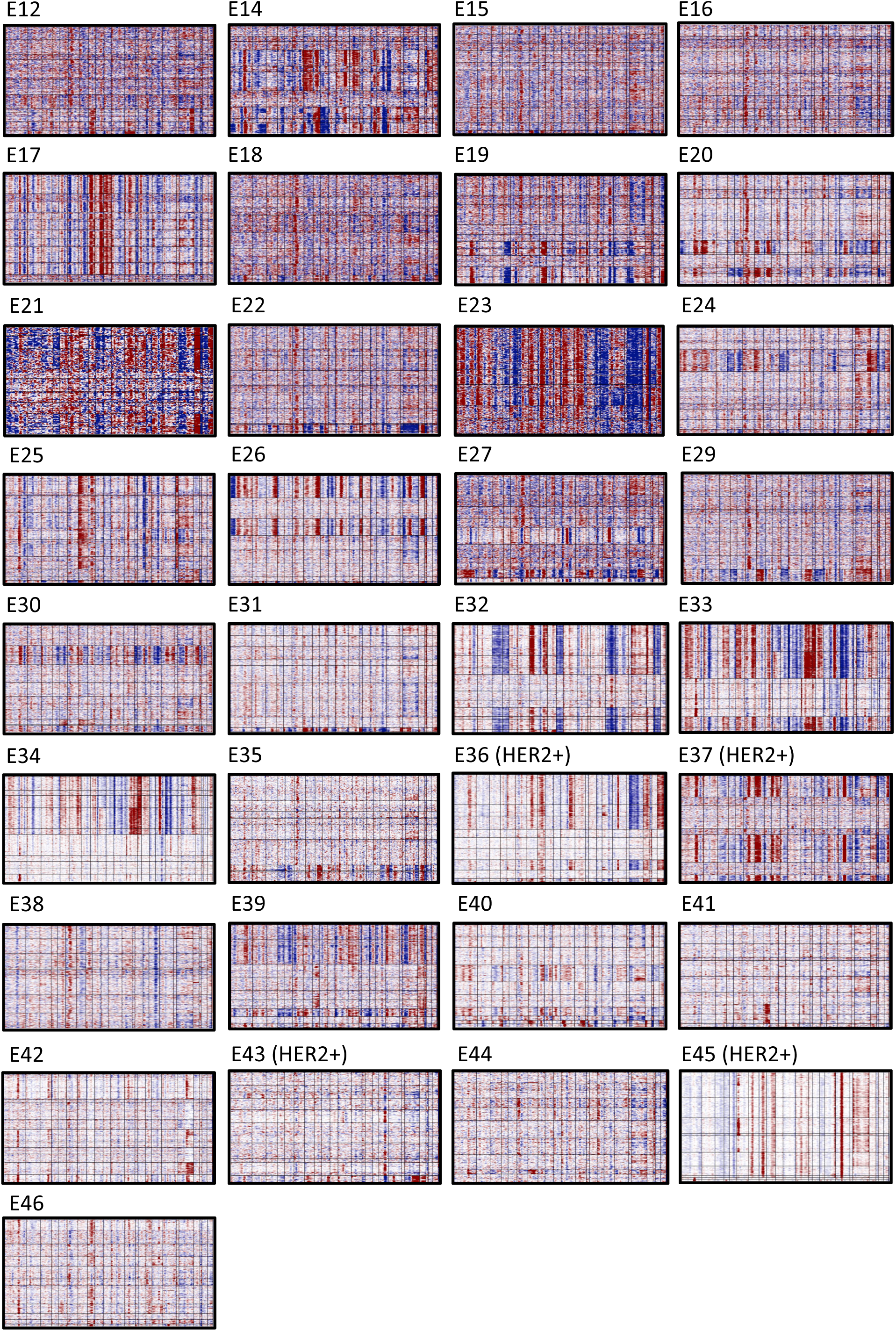
Copy number profiles of all epithelial cells from each patient’s samples inferred using inferCNV.

**Figure S8.**
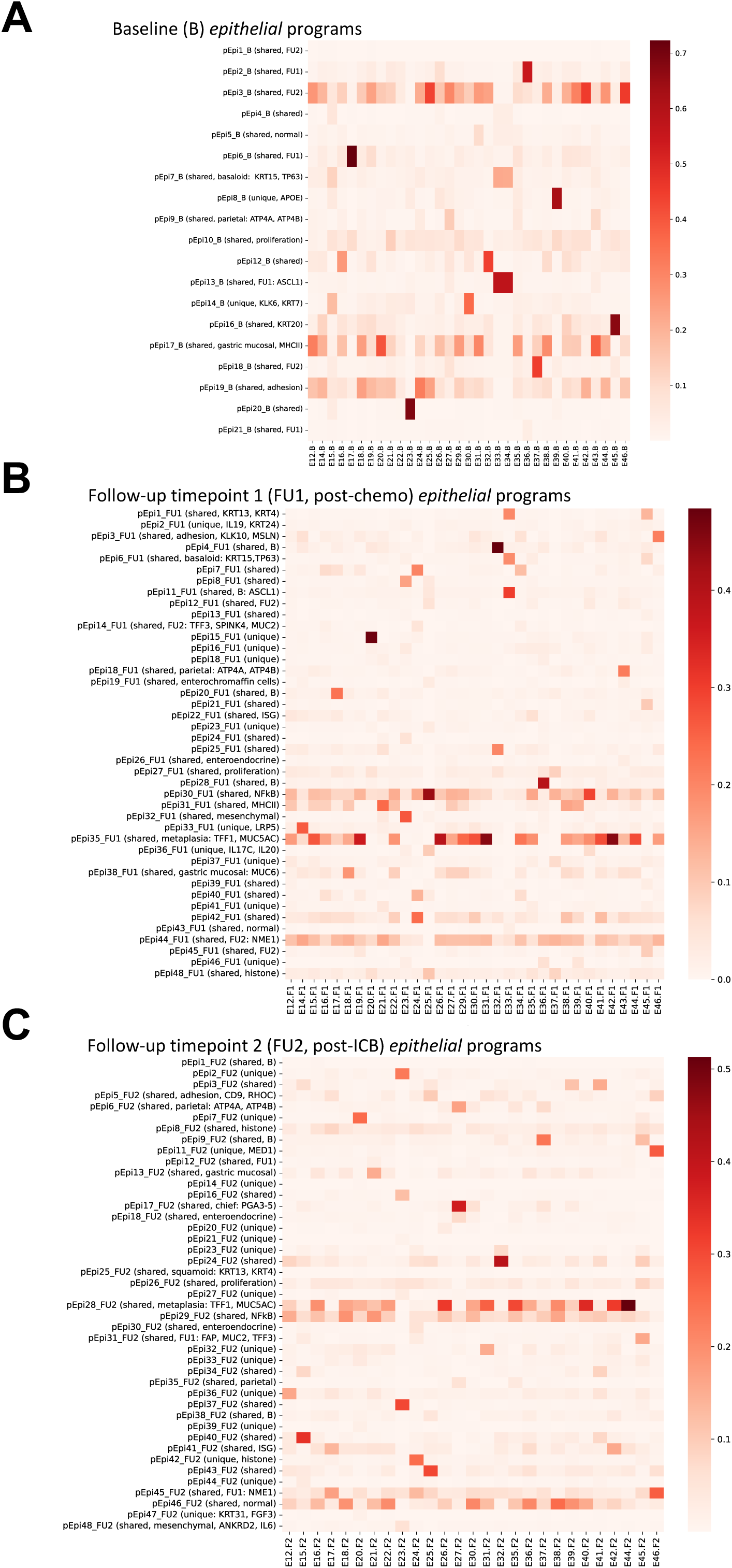
Mean usage of epithelial cNMF programs per sample at (A) baseline, (B) post-chemotherapy (FU1) and (C) post-immunotherapy (FU2).

**Figure S9.**
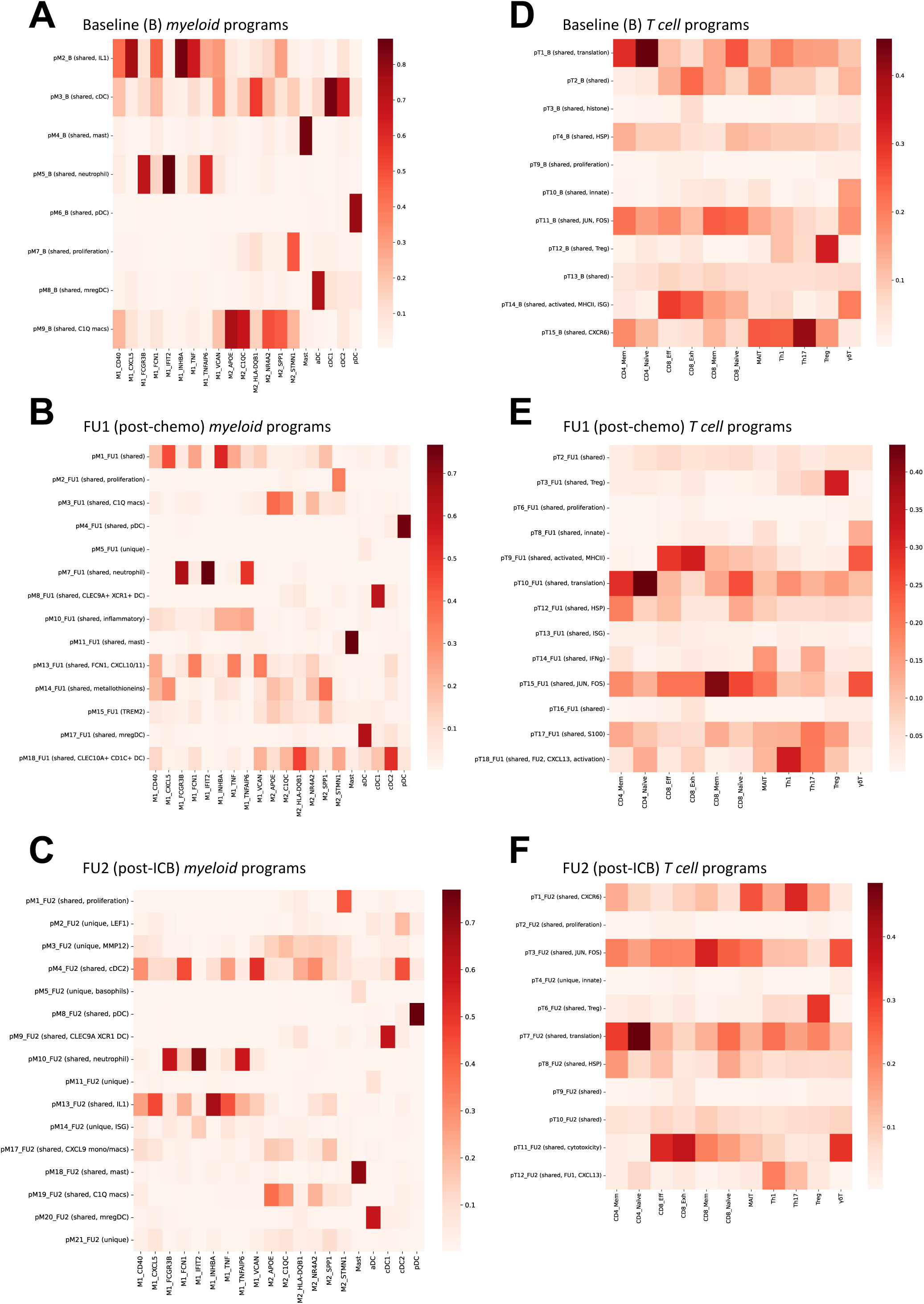
Mean usage of myeloid cNMF programs per myeloid cell subset at (A) baseline, (B) post-chemotherapy (FU1) and (C) post-immunotherapy (FU2). Mean usage of T cell cNMF programs per T cell cell subset at (D) baseline, (E) post-chemotherapy (FU1) and (F) post-immunotherapy (FU2).

**Figure S10.**
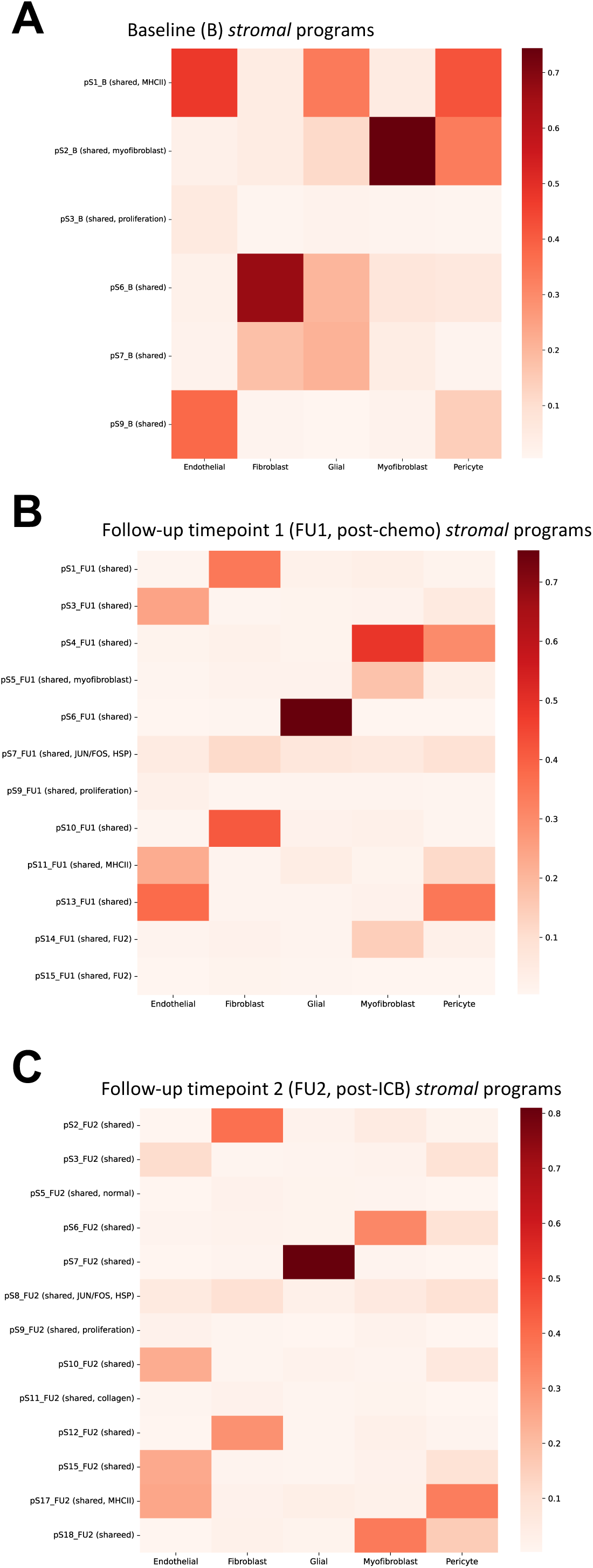
Mean usage of stromal cNMF programs per stromal cell subset at (A) baseline, (B) post-chemotherapy (FU1) and (C) post-immunotherapy (FU2).

**Figure S11.**
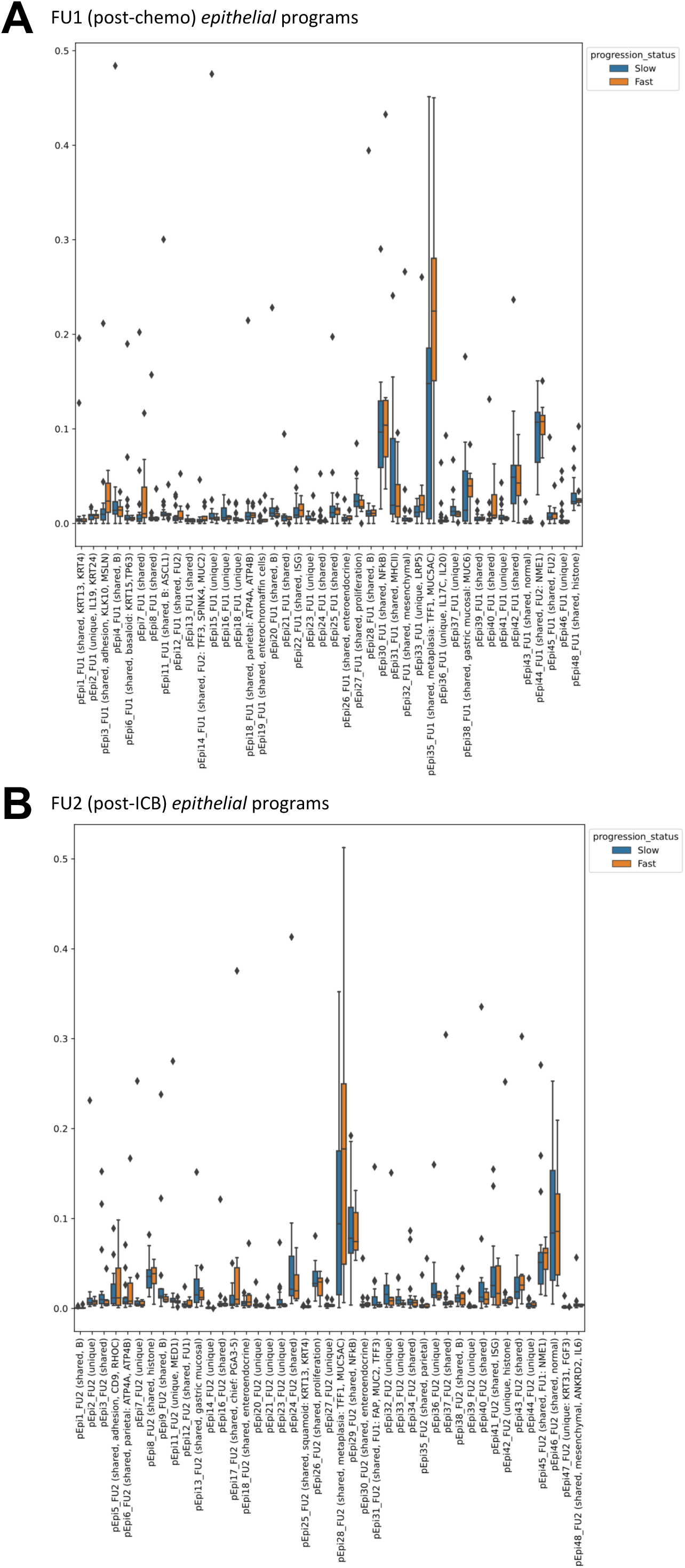
Boxplots showing usage of epithelial cNMF programs in epithelial cells of fast and slow progressors (A) post-chemotherapy (FU1) and (B) post-immunotherapy (FU2).

**Figure S12.**
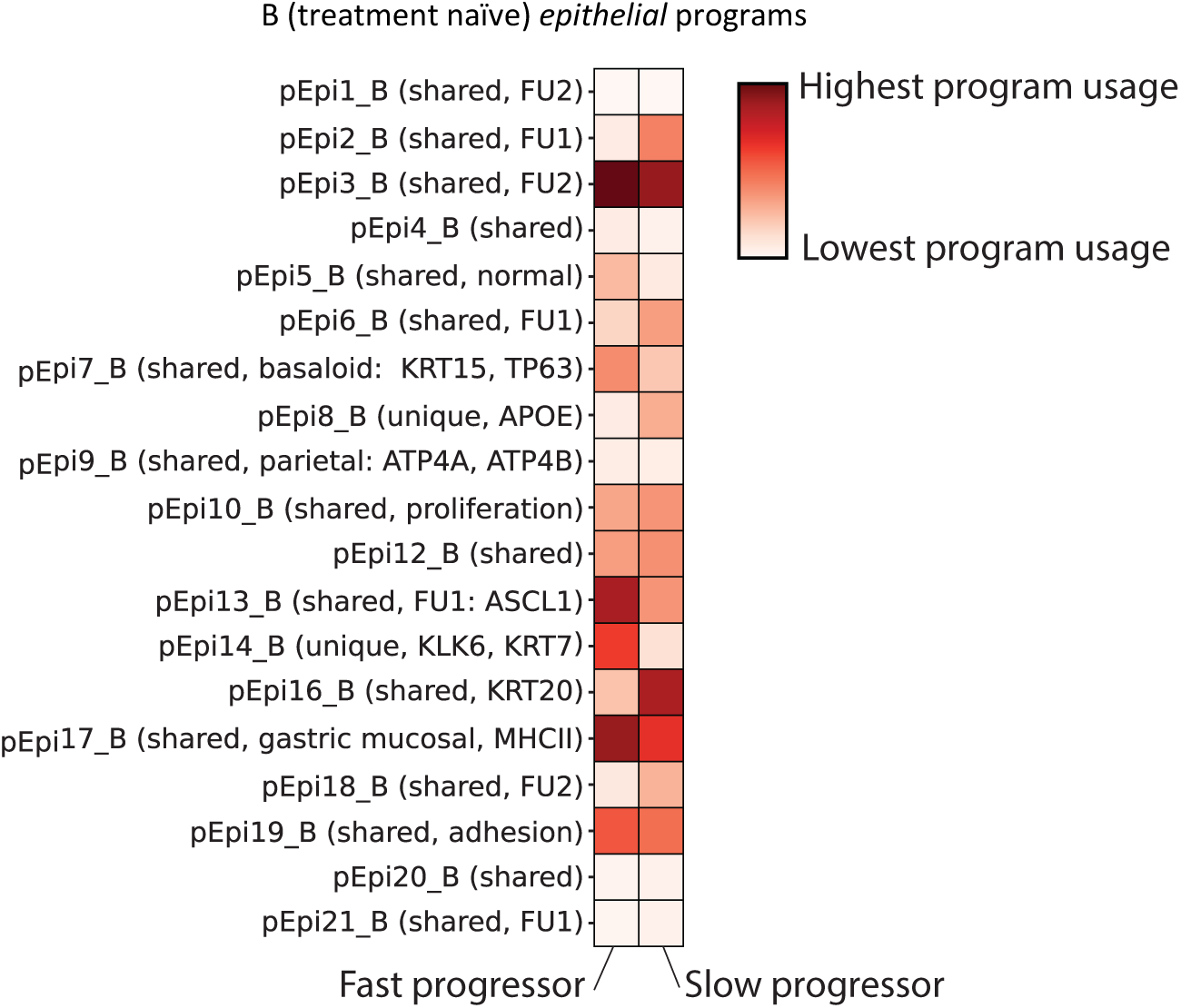
cNMF was performed on epithelial cells at baseline. Shown is the mean usage of each cNMF gene program in the epithelial cells of fast and slow progressing patients.

**Figure S13.**
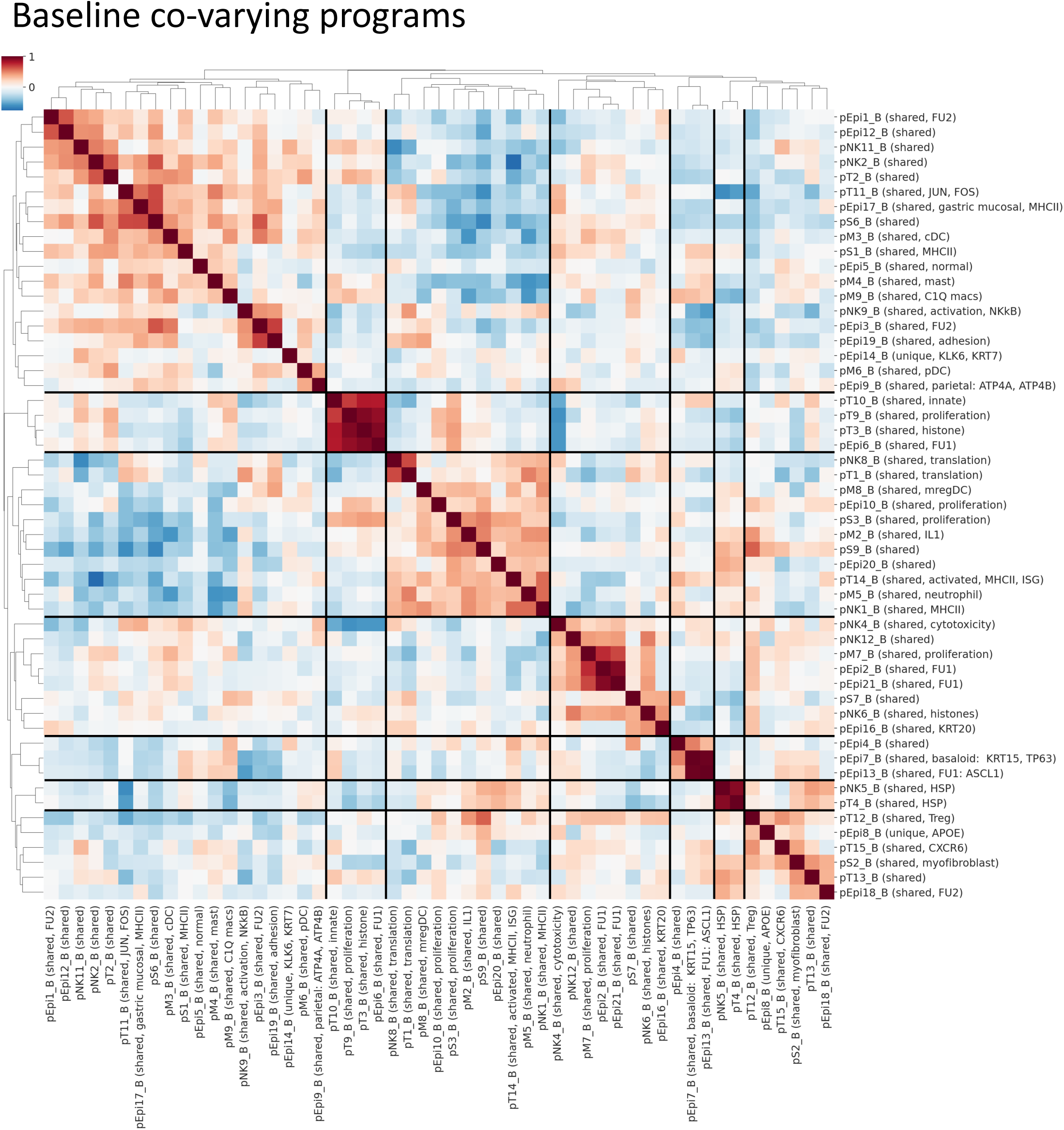
Heatmap showing pairwise correlation of gene program activities across all patient samples at baseline using the 90th percentile of patient-level program activity in epithelial, myeloid, T, NK and stromal cells. Hierarchical clustering was performed to identify clusters of co-varying proteins.

**Figure S14.**
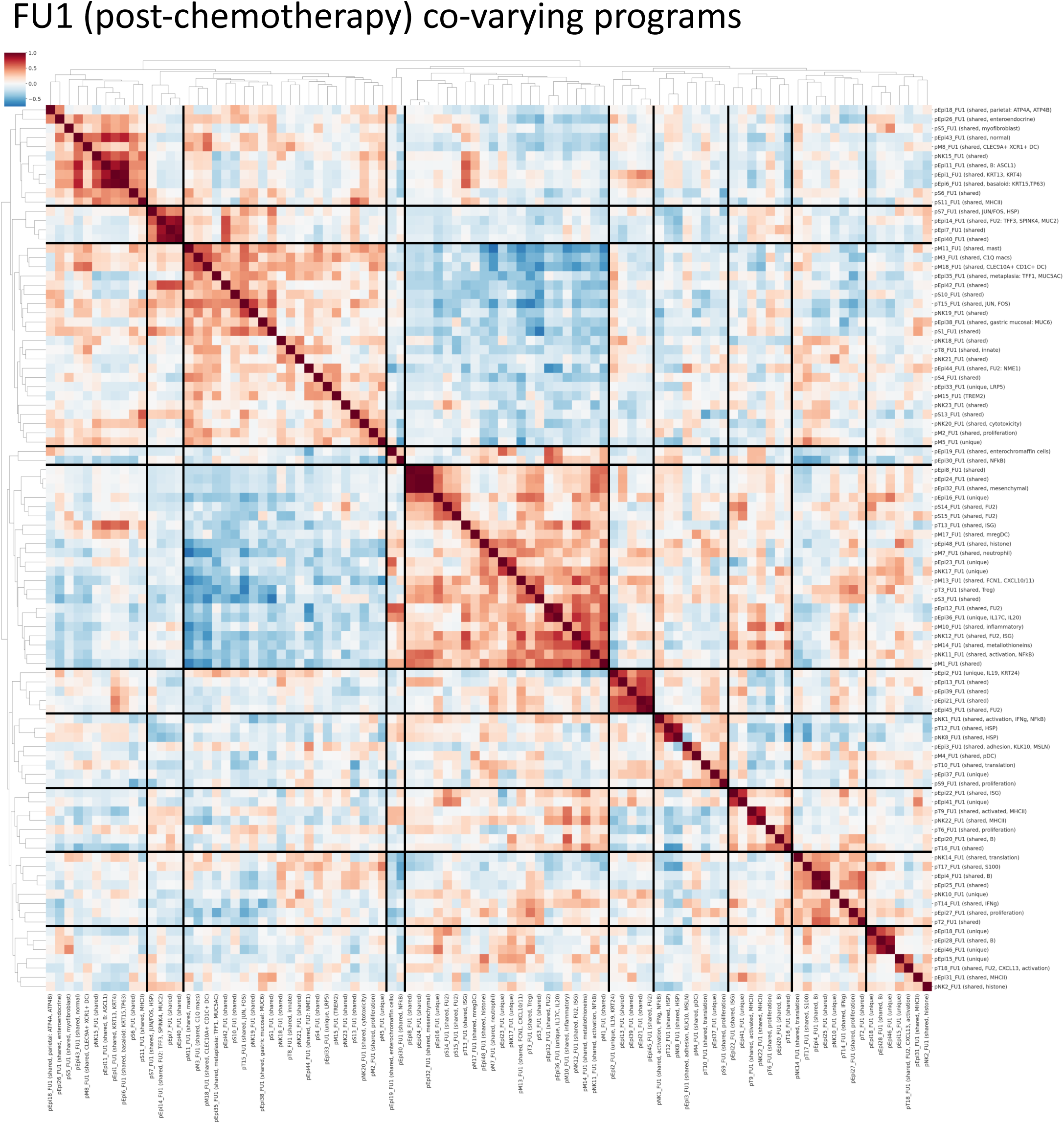
Heatmap showing pairwise correlation of gene program activities across all patient samples post-chemotherapy (FU1) using the 90th percentile of patient-level program activity in epithelial, myeloid, T, NK and stromal cells. Hierarchical clustering was performed to identify clusters of co-varying proteins.

**Figure S15.**
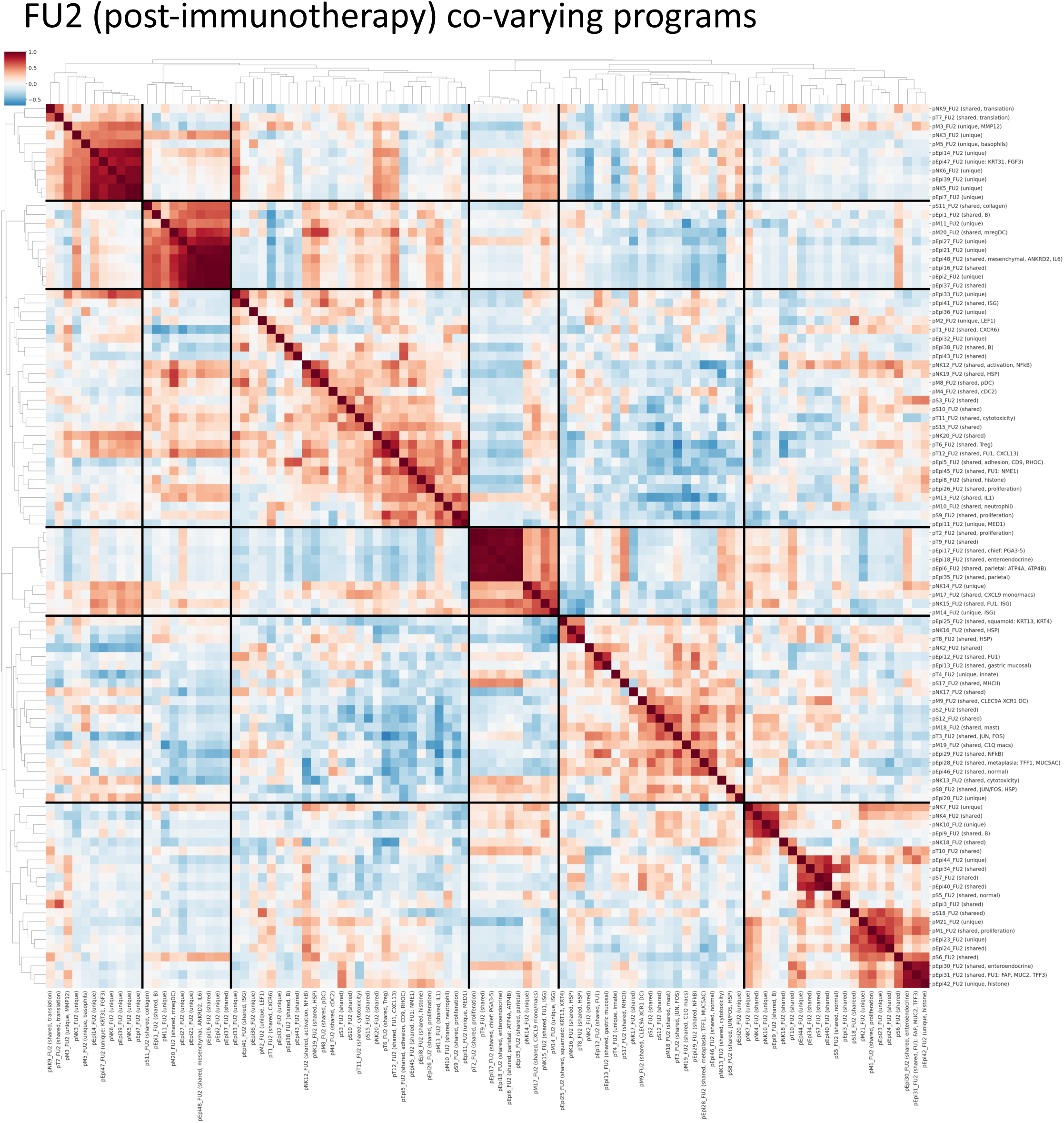
Heatmap showing pairwise correlation of gene program activities across all patient samples post-immunotherapy (FU2) using the 90th percentile of patient-level program activity in epithelial, myeloid, T, NK and stromal cells. Hierarchical clustering was performed to identify clusters of co-varying proteins.

**Figure S16.**
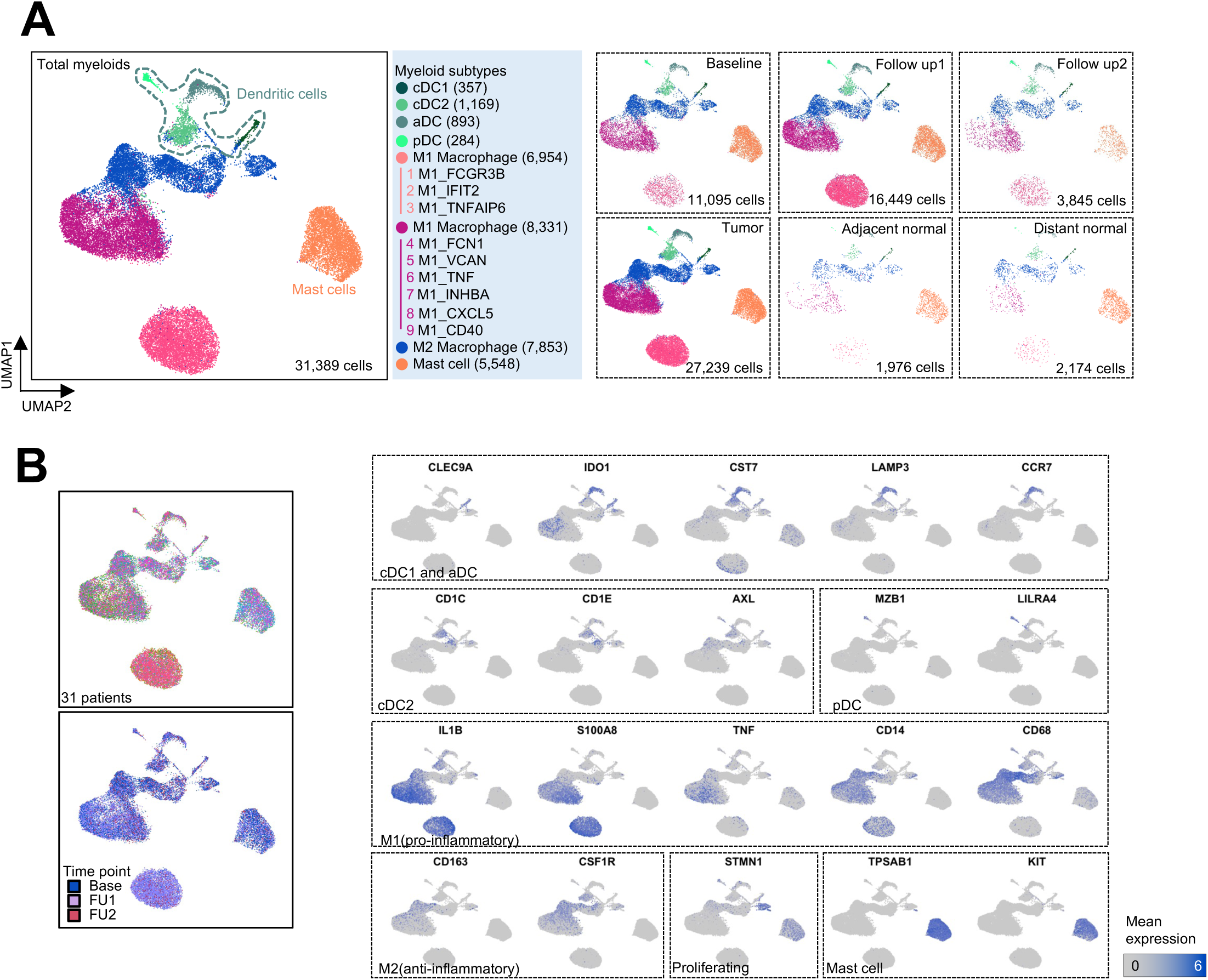
(A) UMAP embedding of single cell transcriptomes of all myeloid cells obtained from all samples in this trial. Labeled are granular myeloid cell types separated by tumor v.s. normal tissue, and by timepoint. (B) UMAP embeddings showing marker gene expression for granular myeloid cell types in (A).

**Figure S17.**
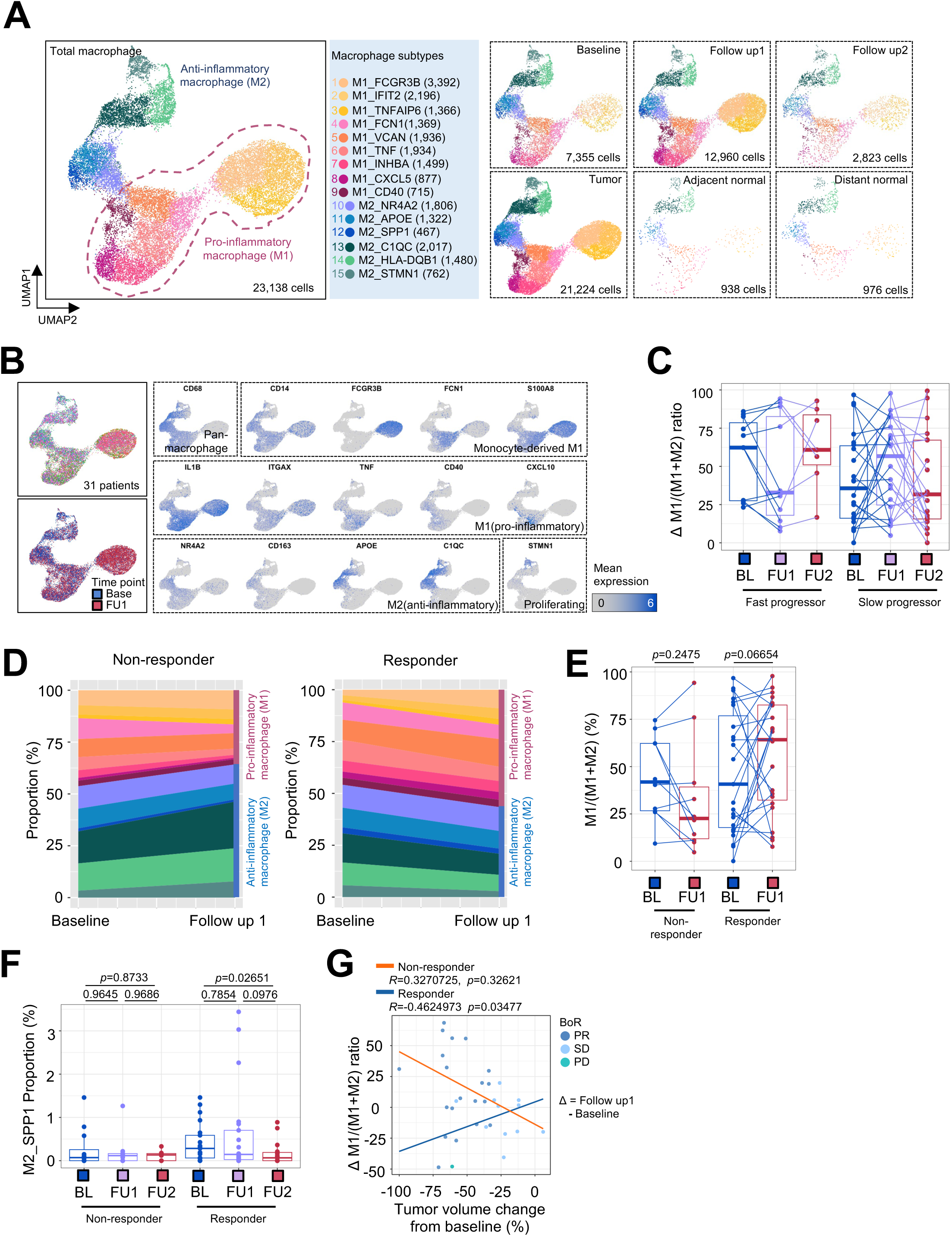
(A) UMAP embedding of single cell transcriptomes of all macrophages obtained from all samples in this trial. Labeled are granular macrophage subsets separated by tumor v.s. normal tissue, and by timepoint. (B) UMAP embeddings showing marker gene expression for macrophage subsets in (A). (C) Relative M1 macrophage proportion in fast and slow progressing patients separated by timepoint. (D) Macrophage subtype proportions at baseline and FU1 in non-responder and responder patients. (E) Relative M1 macrophage proportion in non-responder and responder patients separated by timepoint. (F) Proportion of SPP1+ macrophages in non-responder and responder patients split by timepoint. Statistical comparisons performed using a Wilcoxon signed-rank test. (G) Change in relative M1 proportion from BL to FU1 plotted against change in tumor volume after 1 cycle of chemotherapy, segregated by non-responder and responder patients.

**Figure S18.**
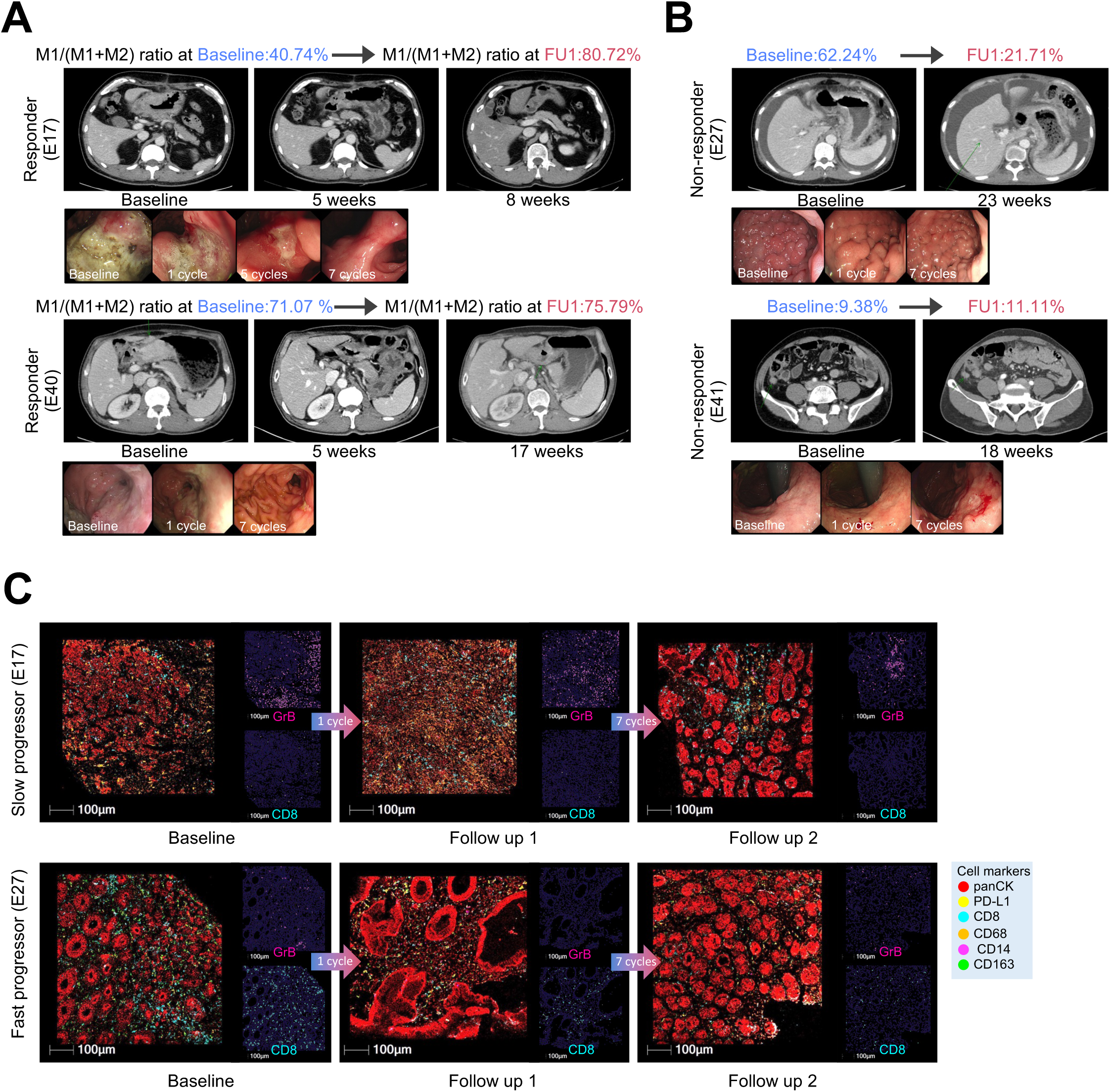
(A) CT imaging of tumor burden in a pair of patients classified as either a responder with increased relative M1 ration or non-responder with unchanged relative M1 ration. (B) CT scans of a second pair of patients as in (A). (C) Multiplexed immunofluorescence (mIF) images of BL, FU1 and FU2 samples from two patients, E17 (slow progressor) and E35 (fast progressor), staining for panCK, PD-L1, CD163, CD68, CD14 and CD8.

**Figure S19.**
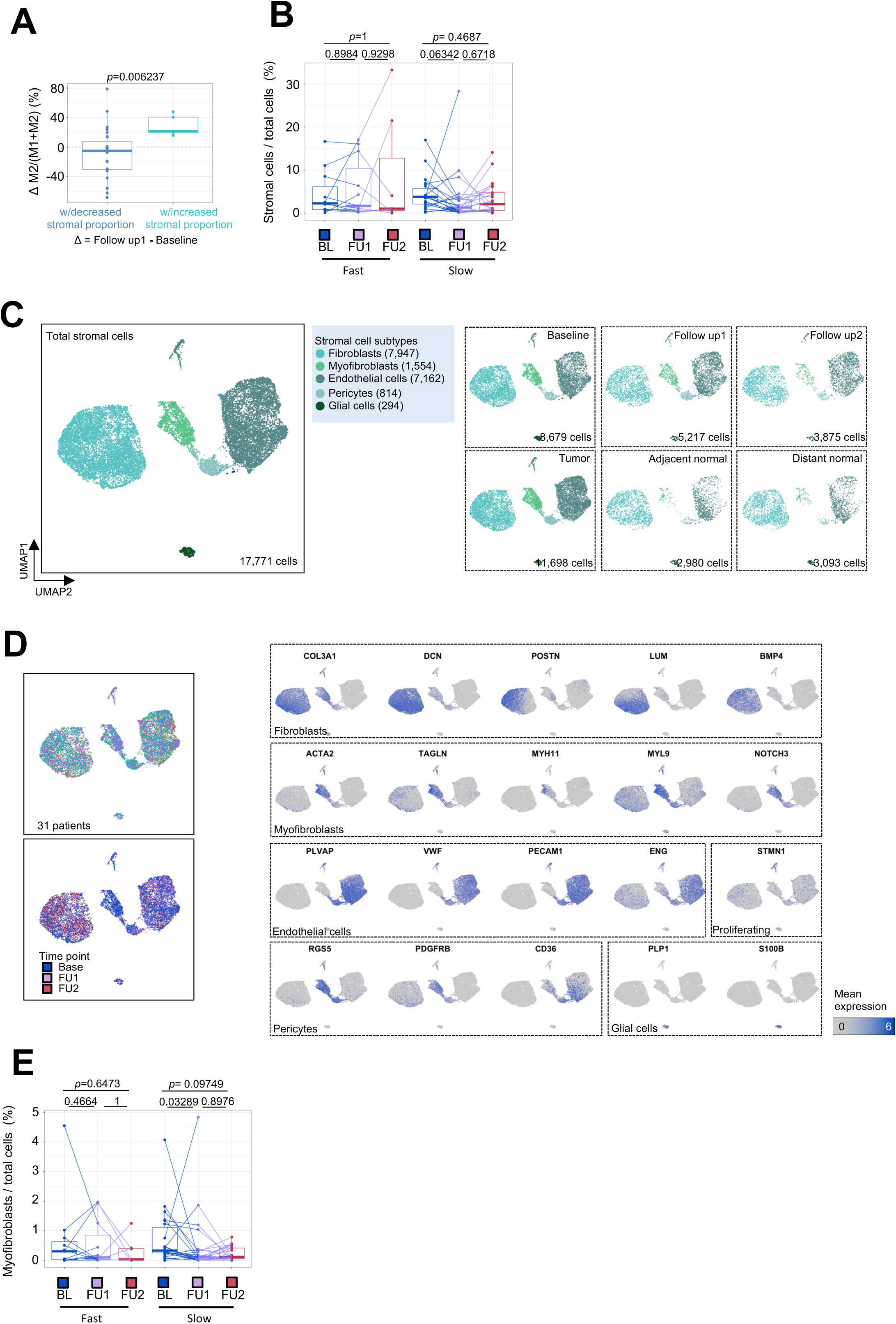
(A) Change in M2 ratio from baseline to FU1 in patients with decreased or increased stromal proportions. (B) Proportion of stromal cells in patient samples from fast and slow progressing patients split by timepoint. (C) UMAP embedding of single cell transcriptomes of all stromal cells obtained from all samples in this trial. Labeled are granular stromal cell types separated by tumor v.s. normal tissue, and by timepoint. (D) UMAP embeddings showing marker gene expression for granular stromal cell types in (C). (E) Proportion of myofibroblasts in patient samples from fast and slow progressing patients split by timepoint.

**Figure S20.**
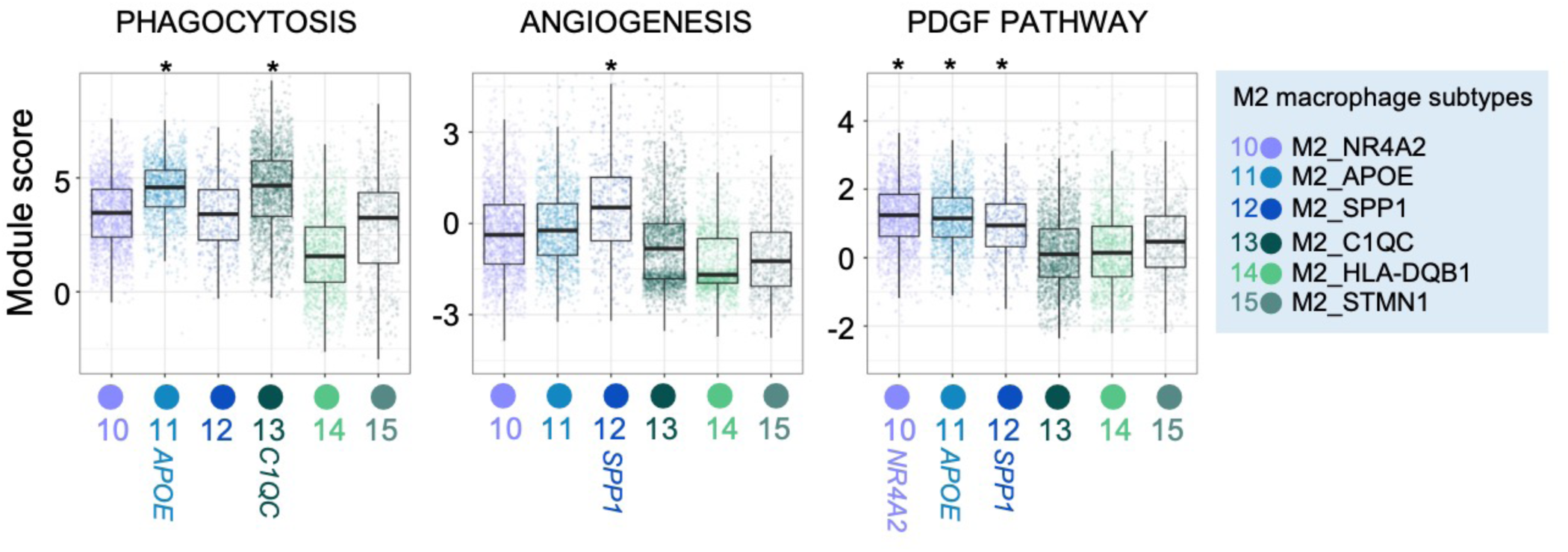
Phagocytosis, angiogenesis and PDGF pathway gene module scores in different M2 macrophage subsets from scRNAseq data.

**Figure S21.**
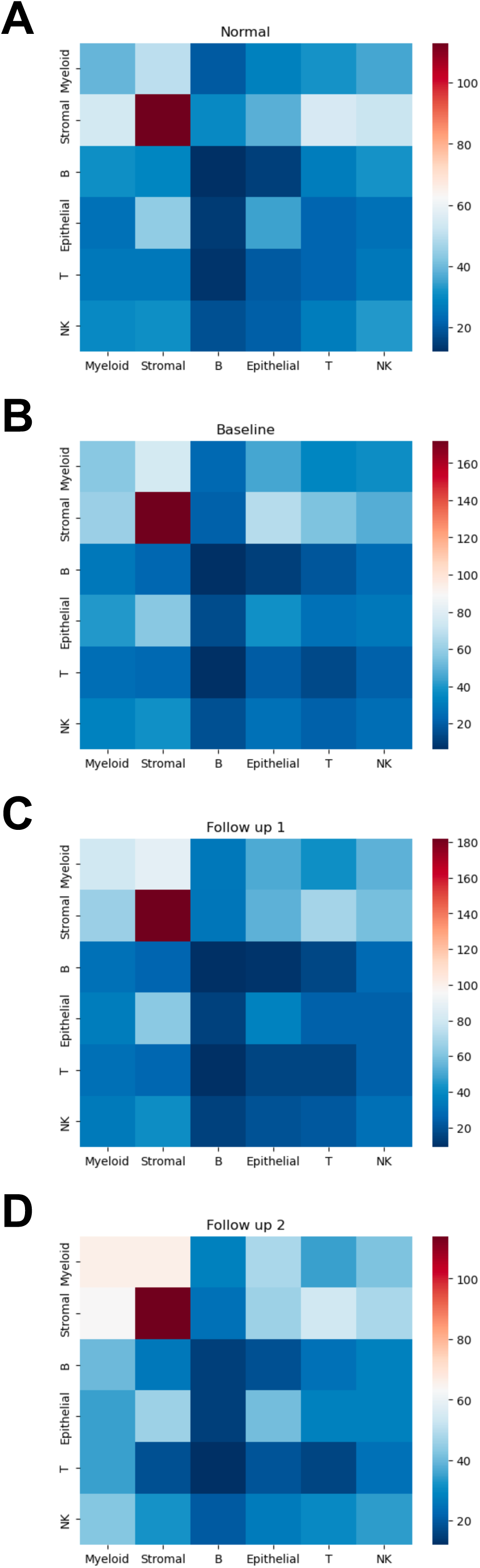
Heatmaps showing the number of significant inferred ligand-receptor interactions between major cell types using CellPhoneDB on scRNAseq data in (A) normal, (B) baseline tumor, (C) post-chemotherapy (FU1) tumor and (D) post-immunotherapy (FU2) tumor samples.

**Figure S22.**
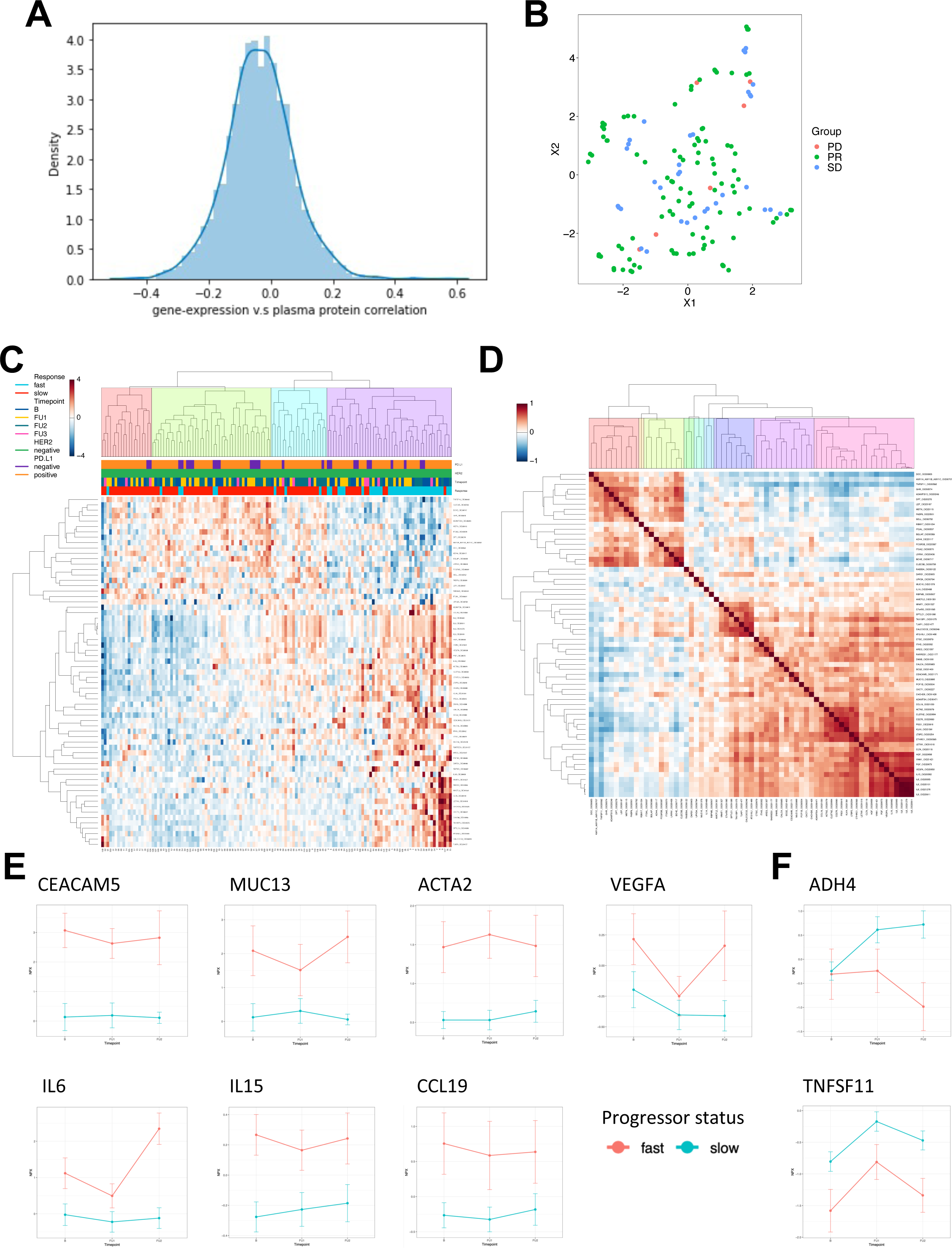
(A) Histogram of all correlations for each protein in plasma (NPX) with respective gene expression in tissue (TPM). (B) UMAP embedding of all patient samples based on plasma protein abundances color coded by patient response status. (C) Heatmap showing differentially abundant plasma proteins between fast and slow progressing patients across all patient samples. (D) Heatmap showing protein-protein correlations for all differentially abundant proteins in fast v.s. slow progressing patients shown for the FU1 timepoint. (E) Point range plots showing temporal changes for several differentially abundant proteins between fast v.s. slow progressing patients.

**Figure S23.**
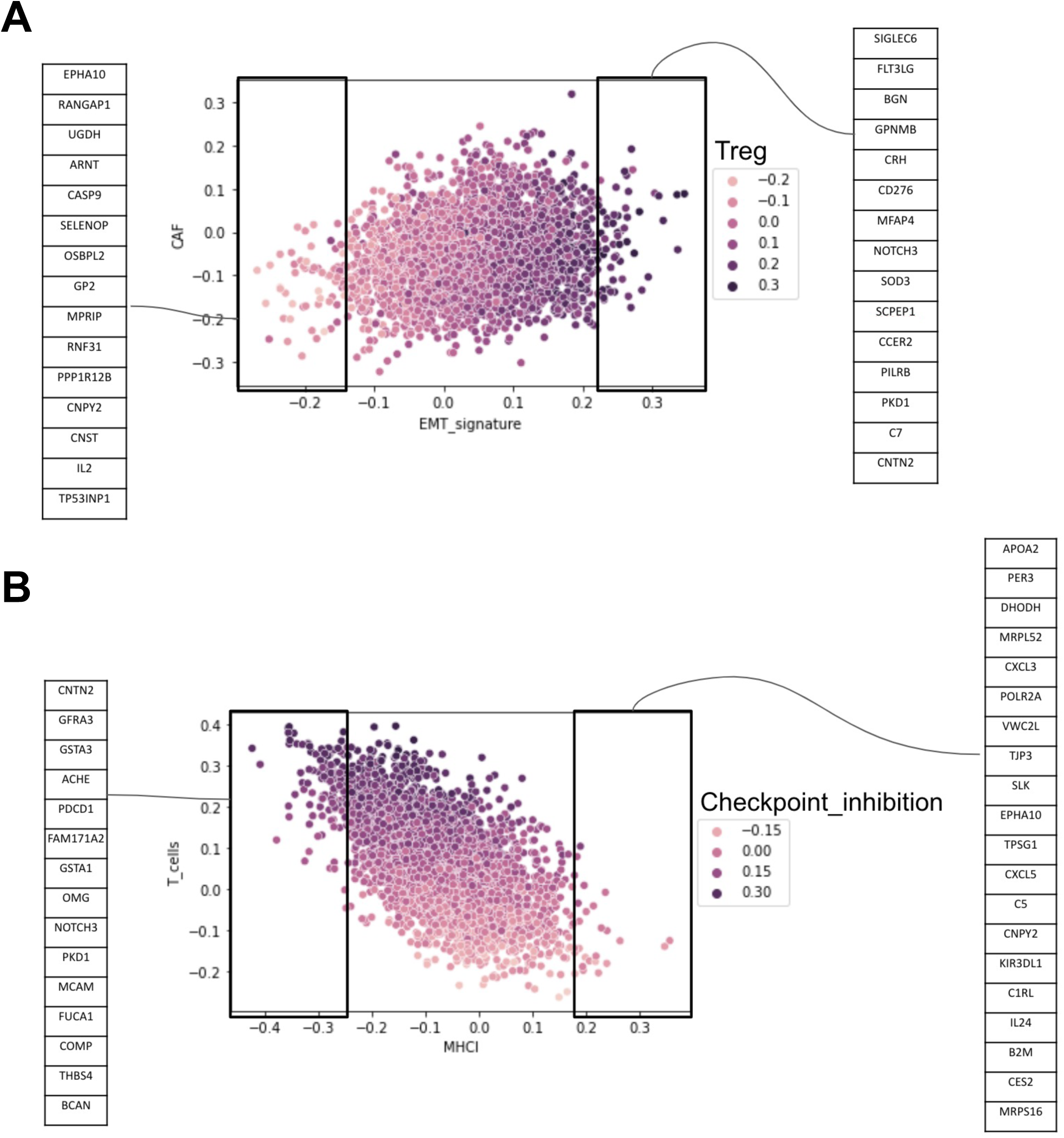
(A) Correlation between plasma protein abundance (NPX) and enrichment of TME CAF and EMT signatures from bulk RNA-seq data. (B) Correlation between plasma protein abundance (NPX) and enrichment of TME T cell and MHCI signatures from bulk RNA-seq data.

**Figure S24.**
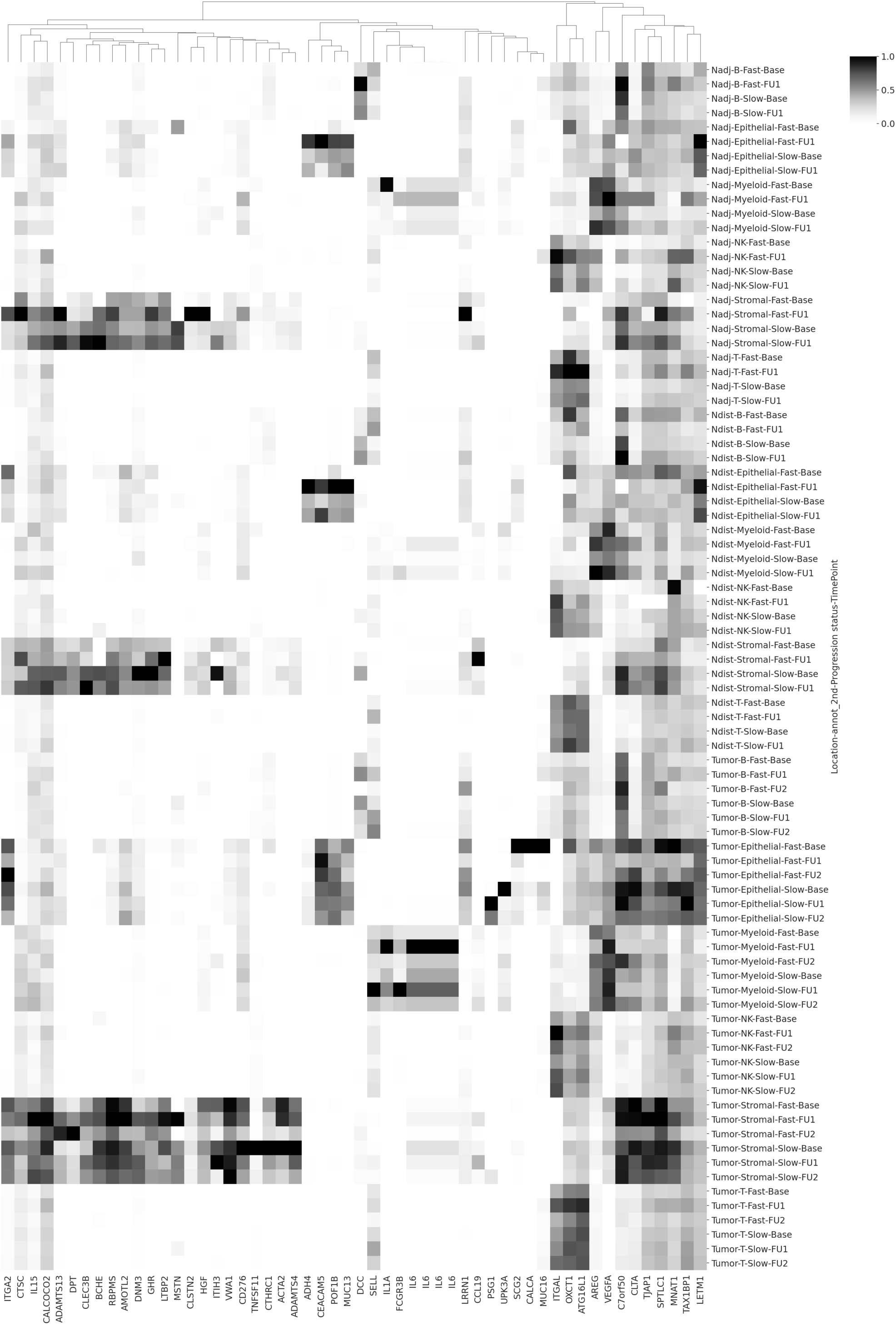
Heatmap demonstrating relative gene expression across normal and tumor broad cell subsets of plasma proteins that are significantly differentially expressed between fast and slow progressing patients.

**Figure S25.**
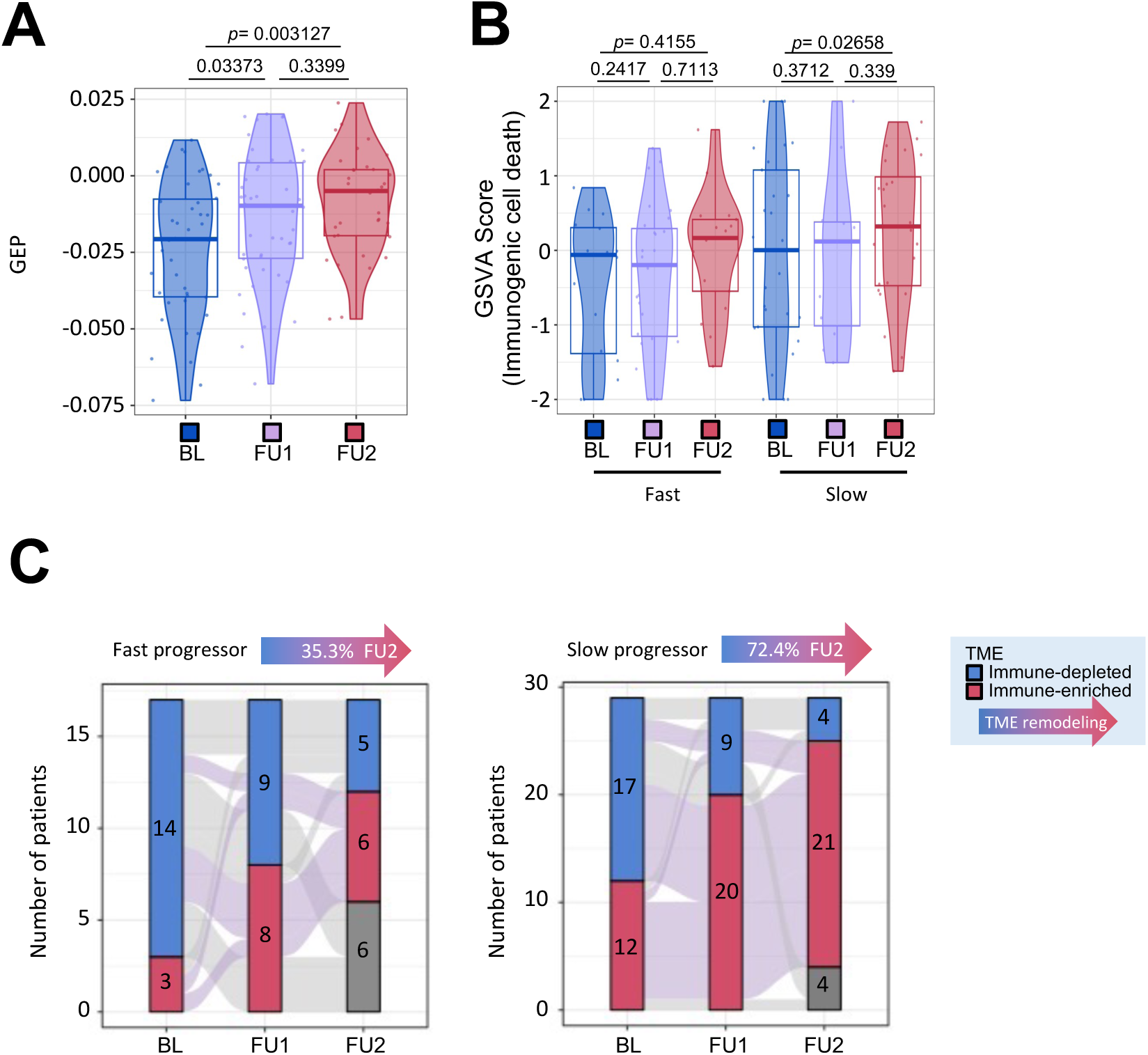
(A) Immune gene expression program signature scored from bulk RNA-seq profiles across all samples separated by timepoint. (B) Immunogenic cell death signatures scored from bulk RNA-seq profiles split by fast and slow progressing patients and by timepoint. (C) Remodeling of TME from immune depleted to immune enriched environments derived from bulk RNA-seq profiles in fast and slow progressing patients, shown across timepoints.

**Figure S26.**
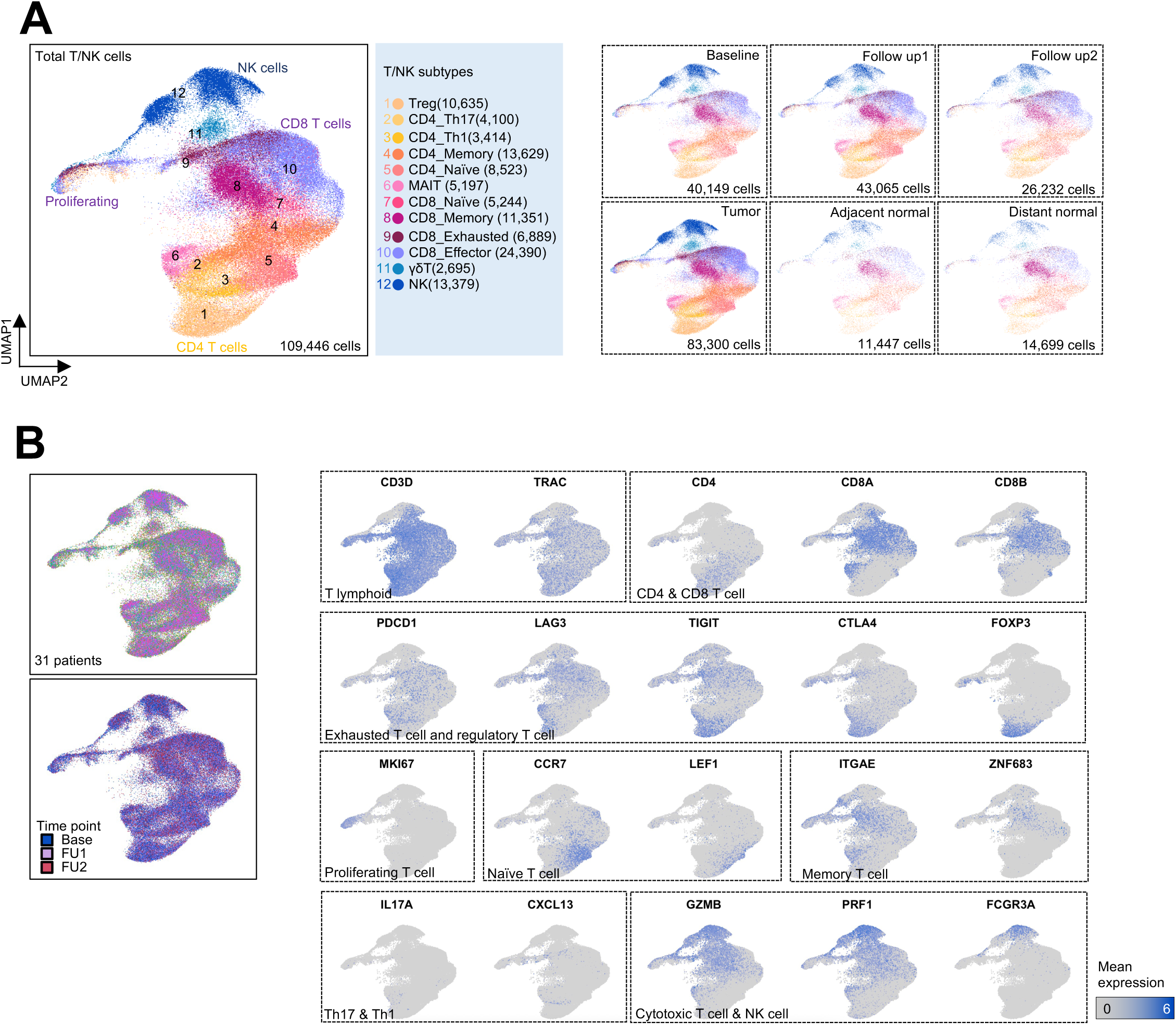
(A) UMAP embedding of single cell transcriptomes of all T cells obtained from all samples in this trial. Labeled are granular T cell subsets separated by tumor v.s. normal tissue, and by timepoint. (B) UMAP embeddings showing marker gene expression for T cell subsets in (A).

**Figure S27.**
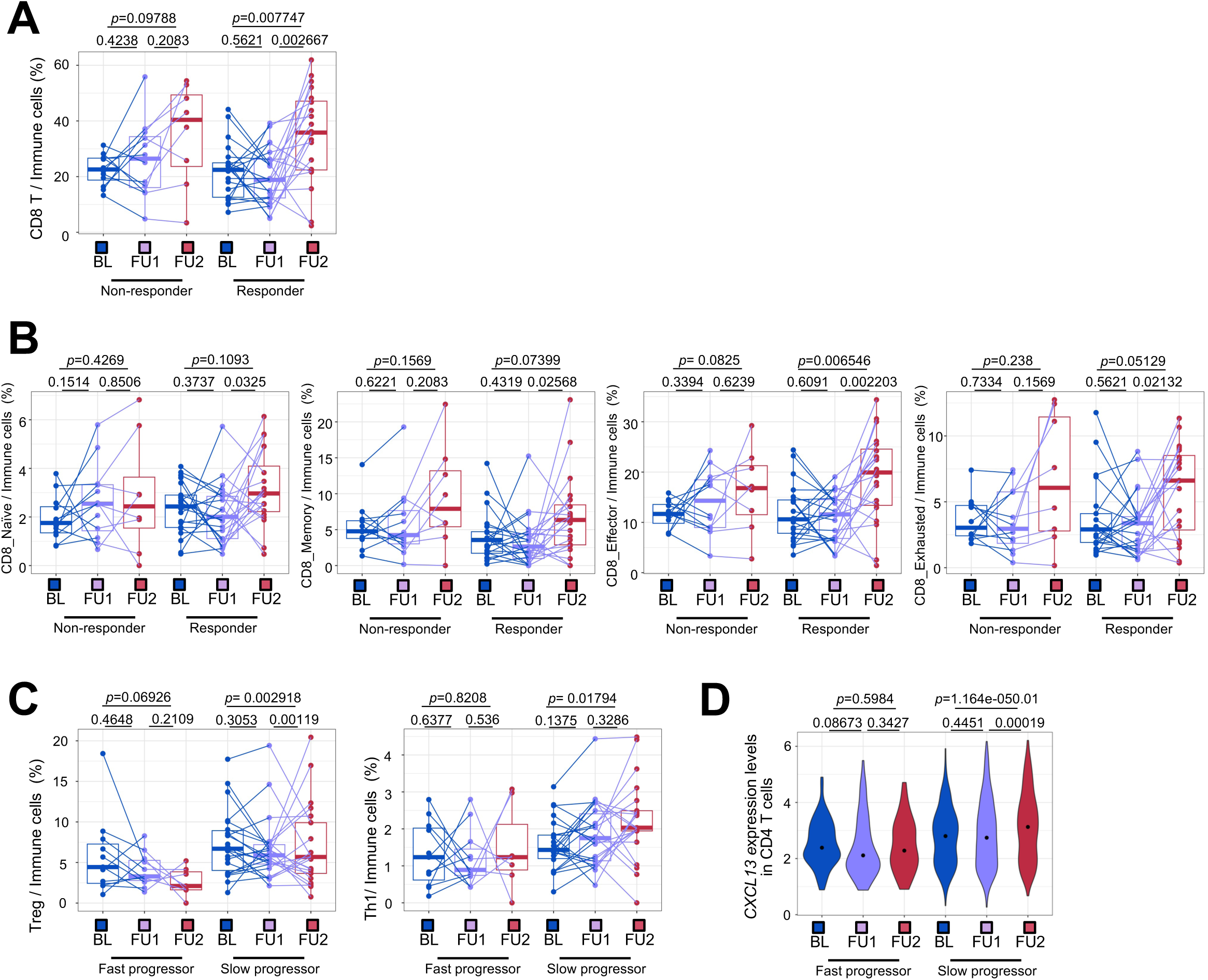
(A) Proportion of CD8 T cells in non-responder and responder patients split by timepoint. Statistical comparisons performed using a Wilcoxon signed-rank test. (B) Proportion of naive, memory, effector and exhausted CD8 T cell subsets in non-responder and responder patients split by timepoint. Statistical comparisons performed using a Wilcoxon signed-rank test. (C) Proportion of regulatory T cell and Th1 CD4 T cell subsets in fast v.s. slow progressor patients split by timepoint. Statistical comparisons performed using a Wilcoxon signed-rank test. (D) Expression of CXCL13 in CD4 T cells in fast v.s. slow progressor patients split by timepoint. Statistical comparisons performed using a Wilcoxon signed-rank test.

**Figure S28.**
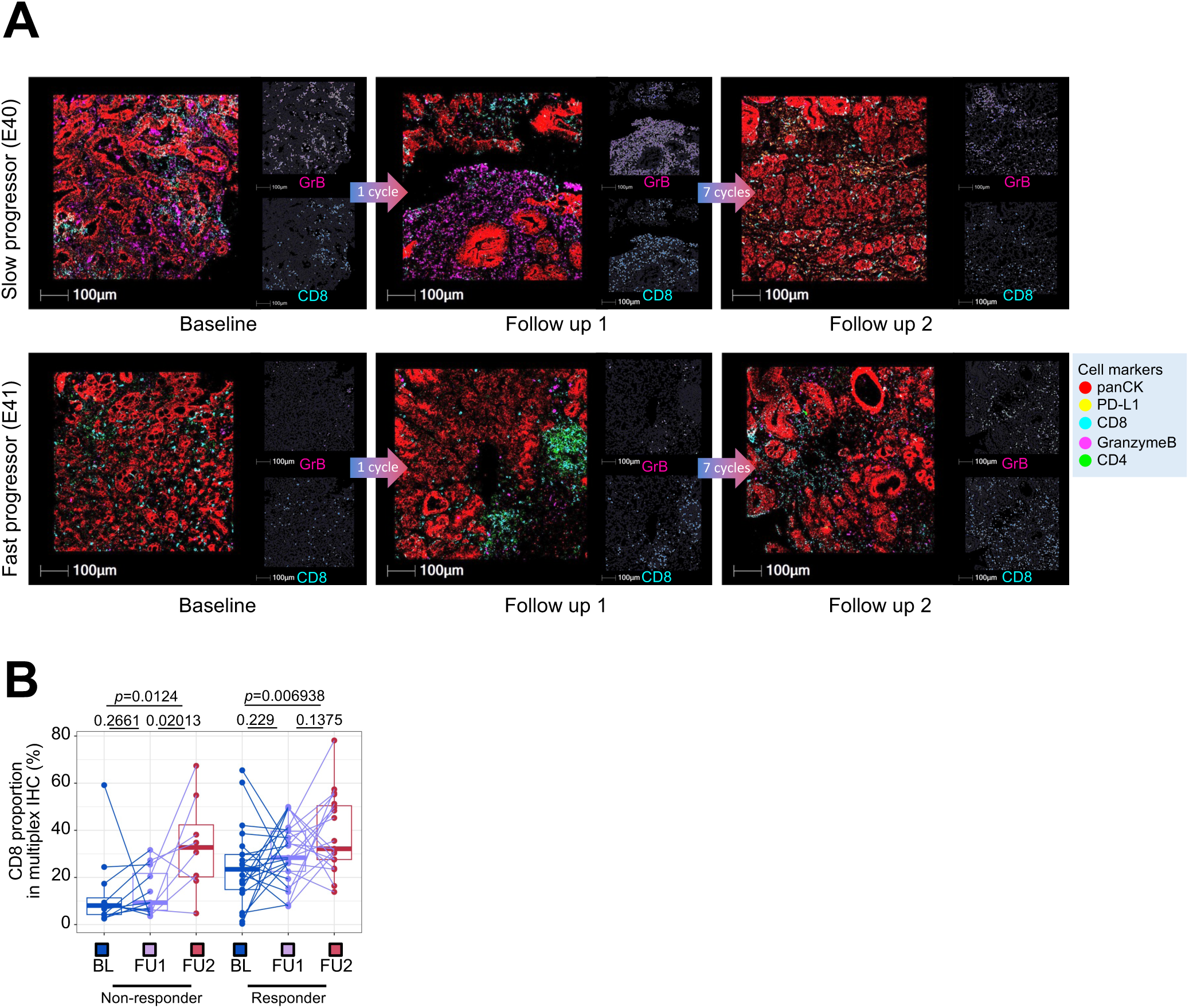
(A) Multiplexed immunofluorescence (mIF) images of BL, FU1 and FU2 samples from two patients, E40 (slow progressor) and E41 (fast progressor), staining for panCK, PD-L1, CD4, CD8 and Granzyme B. (B) Proportion of CD8 T cells macrophages, obtained from mIF images, at BL, FU1 and FU2 in non-responder and responder patients. Statistical comparison performed using a Wilcoxon signed-rank test.

**Figure S29.**
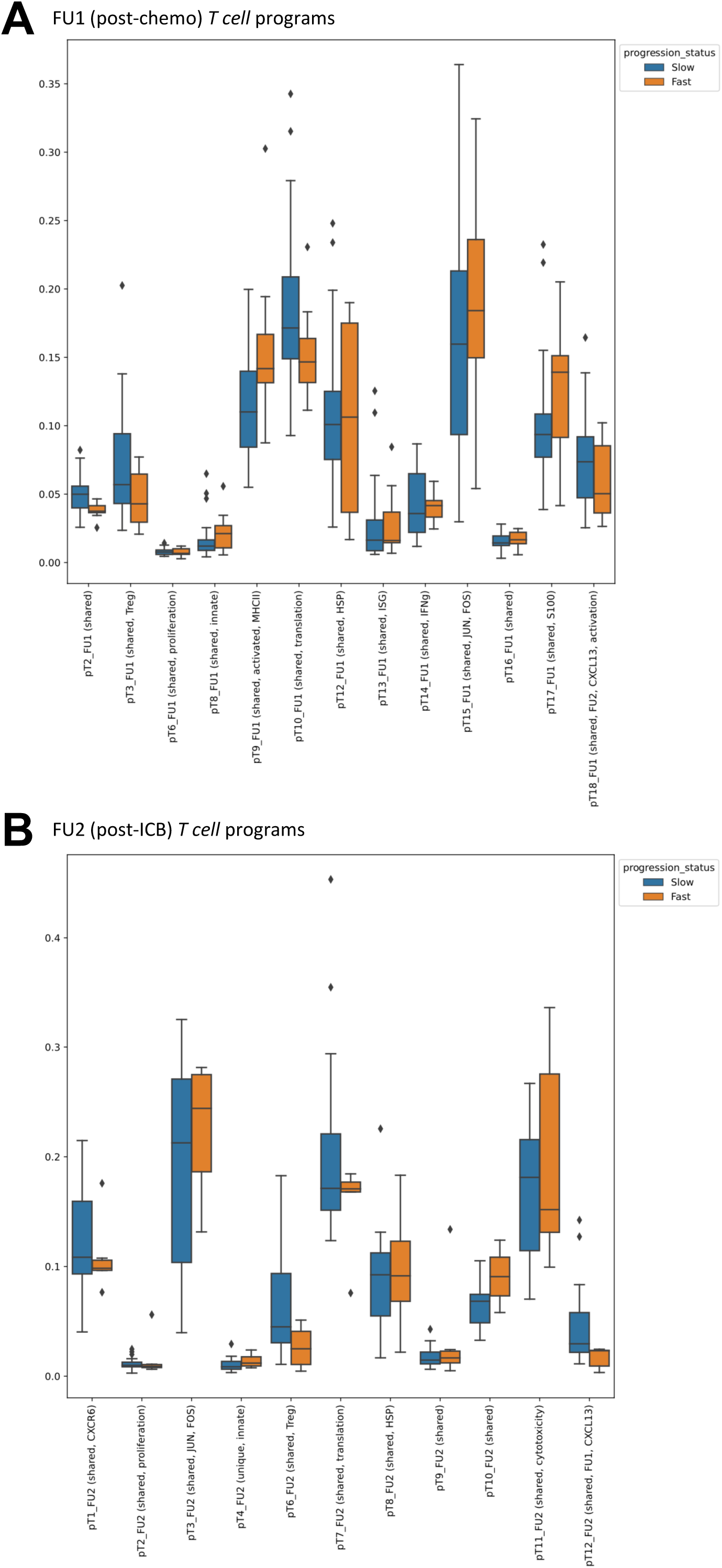
Boxplots showing usage of T cell cNMF programs in T cells of fast and slow progressors (A) post-chemotherapy (FU1) and (B) post-immunotherapy (FU2).

## Supplemental Table Legends

**Table S1.** Demographic data on all patients enrolled in the reported clinical trial.

**Table S2.** Annotated consensus non-negative matrix factorization (cNMF) programs obtained for each cell type from single-cell RNA-sequencing (scRNAseq) data.

**Table S3.** Significantly co-varying cNMF programs across samples by timepoint.

**Table S4.** Significant ligand-receptor interactions between cell types inferred using CellPhoneDB on scRNAseq data.

**Table S5.** Unique ligand-receptor interactions between cell types for each timepoint.

**Table S6.** Significant plasma proteins for time and progression effect obtained using linear mixed effect models (LMMs).

**Table S7.** Correlations between plasma protein abundance and tumor microenvironment subtypes across samples.

## References

1. Yoon, H. H. et al. Association of PD-L1 Expression and Other Variables With Benefit From Immune Checkpoint Inhibition in Advanced Gastroesophageal Cancer: Systematic Review and Meta-analysis of 17 Phase 3 Randomized Clinical Trials. JAMA Oncol 8, 1456–1465 (2022).

2. Shitara, K. et al. Nivolumab plus chemotherapy or ipilimumab in gastro-oesophageal cancer. Nature 603, 942–948 (2022).

3. Kim, R. et al. Early Tumor-Immune Microenvironmental Remodeling and Response to First-Line Fluoropyrimidine and Platinum Chemotherapy in Advanced Gastric Cancer. Cancer Discov. 12, 984–1001 (2022).

4. Tang, X. et al. Neoadjuvant PD-1 blockade plus chemotherapy induces a high pathological complete response rate and anti-tumor immune subsets in clinical stage III gastric cancer. Oncoimmunology 11, 2135819 (2022).

5. Xing, X. et al. Effect of neoadjuvant chemotherapy on the immune microenvironment in gastric cancer as determined by multiplex immunofluorescence and T cell receptor repertoire analysis. J Immunother Cancer 10, (2022).

6. Elhanani, O., Ben-Uri, R. & Keren, L. Spatial profiling technologies illuminate the tumor microenvironment. Cancer Cell (2023) doi:10.1016/j.ccell.2023.01.010.

7. Zaitsev, A. et al. Precise reconstruction of the TME using bulk RNA-seq and a machine learning algorithm trained on artificial transcriptomes. Cancer Cell 40, 879–894.e16 (2022).

8. Schürch, C. M. et al. Coordinated Cellular Neighborhoods Orchestrate Antitumoral Immunity at the Colorectal Cancer Invasive Front. Cell 183, 838 (2020).

9. Pelka, K. et al. Spatially organized multicellular immune hubs in human colorectal cancer. Cell 184, 4734–4752.e20 (2021).

10. Hwang, W. L. et al. Single-nucleus and spatial transcriptome profiling of pancreatic cancer identifies multicellular dynamics associated with neoadjuvant treatment. Nat. Genet. 54, 1178–1191 (2022).

11. Nirmal, A. J. et al. The Spatial Landscape of Progression and Immunoediting in Primary Melanoma at Single-Cell Resolution. Cancer Discov. 12, 1518–1541 (2022).

12. Kotliar, D. et al. Identifying gene expression programs of cell-type identity and cellular activity with single-cell RNA-Seq. Elife 8, (2019).

13. Kang, Y.-K. et al. Nivolumab plus chemotherapy versus placebo plus chemotherapy in patients with HER2-negative, untreated, unresectable advanced or recurrent gastric or gastro-oesophageal junction cancer (ATTRACTION-4): a randomised, multicentre, double-blind, placebo-controlled, phase 3 trial. Lancet Oncol. 23, 234–247 (2022).

14. Cancer Genome Atlas Research Network. .Comprehensive molecular characterization of gastric adenocarcinoma. Nature 513, 202–209 (2014).

15. Cristescu, R. et al. Molecular analysis of gastric cancer identifies subtypes associated with distinct clinical outcomes. Nat. Med. 21, 449–456 (2015).

16. Kim, S. T. et al. Comprehensive molecular characterization of clinical responses to PD-1 inhibition in metastatic gastric cancer. Nat. Med. 24, 1449–1458 (2018).

17. Kwon, M. et al. Determinants of Response and Intrinsic Resistance to PD-1 Blockade in Microsatellite Instability-High Gastric Cancer. Cancer Discov. 11, 2168–2185 (2021).

18. Luo, J. et al. Deciphering radiological stable disease to immune checkpoint inhibitors. Ann. Oncol. 33, 824–835 (2022).

19. Janjigian, Y. Y. et al. First-line nivolumab plus chemotherapy versus chemotherapy alone for advanced gastric, gastro-oesophageal junction, and oesophageal adenocarcinoma (CheckMate 649): a randomised, open-label, phase 3 trial. Lancet 398, 27–40 (2021).

20. Sun, J.-M. et al. Pembrolizumab plus chemotherapy versus chemotherapy alone for first-line treatment of advanced oesophageal cancer (KEYNOTE-590): a randomised, placebo-controlled, phase 3 study. Lancet 398, 759–771 (2021).

21. Janjigian, Y. Y. et al. Genetic Predictors of Response to Systemic Therapy in Esophagogastric Cancer. Cancer Discov. 8, 49–58 (2018).

22. Sun, K. et al. scRNA-seq of gastric tumor shows complex intercellular interaction with an alternative T cell exhaustion trajectory. Nat. Commun. 13, 4943 (2022).

23. Kumar, V. et al. Single-Cell Atlas of Lineage States, Tumor Microenvironment, and Subtype-Specific Expression Programs in Gastric Cancer. Cancer Discov. 12, 670–691 (2022).

24. Zhang, M. et al. Dissecting transcriptional heterogeneity in primary gastric adenocarcinoma by single cell RNA sequencing. Gut 70, 464–475 (2021).

25. Patel, A. P. et al. Single-cell RNA-seq highlights intratumoral heterogeneity in primary glioblastoma. Science 344, 1396–1401 (2014).

26. Gao, T. et al. Haplotype-aware analysis of somatic copy number variations from single-cell transcriptomes. Nat. Biotechnol. (2022) doi:10.1038/s41587-022-01468-y.

27. Bagaev, A. et al. Conserved pan-cancer microenvironment subtypes predict response to immunotherapy. Cancer Cell 39, 845–865.e7 (2021).

28. Helmink, B. A. et al. B cells and tertiary lymphoid structures promote immunotherapy response. Nature 577, 549–555 (2020).

29. Barkley, D. et al. Cancer cell states recur across tumor types and form specific interactions with the tumor microenvironment. Nat. Genet. 54, 1192–1201 (2022).

30. Neftel, C. et al. An Integrative Model of Cellular States, Plasticity, and Genetics for Glioblastoma. Cell 178, 835–849.e21 (2019).

31. Leung, W. K. et al. Expression of trefoil peptides (TFF1, TFF2, and TFF3) in gastric carcinomas, intestinal metaplasia, and non-neoplastic gastric tissues. J. Pathol. 197, 582–588 (2002).

32. Song, J.-Y. et al. Expression of MUC5AC and Trefoil Peptide 1 (TFF1) in the Subtypes of Intestinal Metaplasia. Clin. Endosc. 45, 151–154 (2012).

33. Lenos, K. J. et al. Molecular characterization of colorectal cancer related peritoneal metastatic disease. Nat. Commun. 13, 4443 (2022).

34. Hsu, H.-J. et al. Eradicating mesothelin-positive human gastric and pancreatic tumors in xenograft models with optimized anti-mesothelin antibody–drug conjugates from synthetic antibody libraries. Sci. Rep. 11, 15430 (2021).

35. Einama, T. et al. Luminal membrane expression of mesothelin is a prominent poor prognostic factor for gastric cancer. Br. J. Cancer 107, 137–142 (2012).

36. Hanada, K.-I. et al. A phenotypic signature that identifies neoantigen-reactive T cells in fresh human lung cancers. Cancer Cell 40, 479–493.e6 (2022).

37. Liu, B., Zhang, Y., Wang, D., Hu, X. & Zhang, Z. Single-cell meta-analyses reveal responses of tumor-reactive CXCL13 T cells to immune-checkpoint blockade. Nat Cancer 3, 1123–1136 (2022).

38. Kersten, K. et al. Spatiotemporal co-dependency between macrophages and exhausted CD8+ T cells in cancer. Cancer Cell 40, 624–638.e9 (2022).

39. Mantovani, A., Allavena, P., Marchesi, F. & Garlanda, C. Macrophages as tools and targets in cancer therapy. Nat. Rev. Drug Discov. 21, 799–820 (2022).

40. Nixon, B. G. et al. Tumor-associated macrophages expressing the transcription factor IRF8 promote T cell exhaustion in cancer. Immunity 55, 2044–2058.e5 (2022).

41. Mantovani, A., Marchesi, F., Malesci, A., Laghi, L. & Allavena, P. Tumour-associated macrophages as treatment targets in oncology. Nat. Rev. Clin. Oncol. 14, 399–416 (2017).

42. Cheng, S. et al. A pan-cancer single-cell transcriptional atlas of tumor infiltrating myeloid cells. Cell 184, 792–809.e23 (2021).

43. Zhang, Y. et al. Single-cell analyses reveal key immune cell subsets associated with response to PD-L1 blockade in triple-negative breast cancer. Cancer Cell 39, 1578–1593.e8 (2021).

44. Efremova, M., Vento-Tormo, M., Teichmann, S. A. & Vento-Tormo, R. CellPhoneDB v2.0: Inferring cell-cell communication from combined expression of multi-subunit receptor-ligand complexes. Preprint at https://doi.org/10.1101/680926.

45. Yang, R. et al. Galectin-9 interacts with PD-1 and TIM-3 to regulate T cell death and is a target for cancer immunotherapy. Nature (2021).

46. Dubrot, J. et al. In vivo CRISPR screens reveal the landscape of immune evasion pathways across cancer. Nat. Immunol. 23, 1495–1506 (2022).

47. Xie, X.-W., Jiang, S.-S. & Li, X. CLEC3B as a Potential Prognostic Biomarker in Hepatocellular Carcinoma. Front Mol Biosci 7, 614034 (2020).

48. Sun, J. et al. CLEC3B as a potential diagnostic and prognostic biomarker in lung cancer and association with the immune microenvironment. Cancer Cell Int. 20, 106 (2020).

49. Ayers, M. et al. IFN-γ-related mRNA profile predicts clinical response to PD-1 blockade. J. Clin. Invest. 127, 2930–2940 (2017).

50. Ott, P. A., et al. T-Cell-Inflamed Gene-Expression Profile, Programmed Death Ligand 1 Expression, and Tumor Mutational Burden Predict Efficacy in Patients Treated With Pembrolizumab Across 20 Cancers: KEYNOTE-028. J. Clin. Oncol. 37, 318–327 (2019).

51. Cristescu, R. et al. Transcriptomic Determinants of Response to Pembrolizumab Monotherapy across Solid Tumor Types. Clin. Cancer Res. 28, 1680–1689 (2022).

52. Galluzzi, L., Buqué, A., Kepp, O., Zitvogel, L. & Kroemer, G. Immunogenic cell death in cancer and infectious disease. Nat. Rev. Immunol. 17, 97–111 (2017).

53. Maier, B. et al. A conserved dendritic-cell regulatory program limits antitumour immunity. Nature 580, 257–262 (2020).

54. Huseni, M. A. et al. CD8+ T cell-intrinsic IL-6 signaling promotes resistance to anti-PD-L1 immunotherapy. Cell Rep Med 100878 (2022).

55. Mehta, A. et al. Plasma proteomic biomarkers identify non-responders and reveal biological insights about the tumor microenvironment in melanoma patients after PD1 blockade. Preprint at https://doi.org/10.1101/2022.02.02.478819.

56. Fisher, D. T., Appenheimer, M. M. & Evans, S. S. The two faces of IL-6 in the tumor microenvironment. Semin. Immunol. 26, 38–47 (2014).

57. Revel, M., Sautès-Fridman, C., Fridman, W.-H. & Roumenina, L. T. C1q+ macrophages: passengers or drivers of cancer progression. Trends Cancer Res. 8, 517–526 (2022).

58. Xiao, G., Deng, A., Liu, H., Ge, G. & Liu, X. Activator protein 1 suppresses antitumor T-cell function via the induction of programmed death 1. Proc. Natl. Acad. Sci. U. S. A. 109, 15419–15424 (2012).

59. Mullins, R. D. Z., Pal, A., Barrett, T. F., Heft Neal, M. E. & Puram, S. V. Epithelial-Mesenchymal Plasticity in Tumor Immune Evasion. Cancer Res. 82, 2329–2343 (2022).

60. Wang, M. et al. Acquired semi-squamatization during chemotherapy suggests differentiation as a therapeutic strategy for bladder cancer. Cancer Cell 40, 1044–1059.e8 (2022).

61. Wolf, D. M. et al. Redefining breast cancer subtypes to guide treatment prioritization and maximize response: Predictive biomarkers across 10 cancer therapies. Cancer Cell 40, 609–623.e6 (2022).

62. Li, K. et al. Multi-omic analyses of changes in the tumor microenvironment of pancreatic adenocarcinoma following neoadjuvant treatment with anti-PD-1 therapy. Cancer Cell 40, 1374– 1391.e7 (2022).

63. Chen, B. et al. Differential pre-malignant programs and microenvironment chart distinct paths to malignancy in human colorectal polyps. Cell 184, 6262–6280.e26 (2021).

64. Hingorani, S. R. Epithelial and stromal co-evolution and complicity in pancreatic cancer. Nat. Rev. Cancer 23, 57–77 (2023).

65. Labrie, M., Brugge, J. S., Mills, G. B. & Zervantonakis, I. K. Therapy resistance: opportunities created by adaptive responses to targeted therapies in cancer. Nat. Rev. Cancer 22, 323–339 (2022).

66. Kim, T. K., Vandsemb, E. N., Herbst, R. S. & Chen, L. Adaptive immune resistance at the tumour site: mechanisms and therapeutic opportunities. Nat. Rev. Drug Discov. 21, 529–540 (2022).

67. Shi, T. et al. DKK1 Promotes Tumor Immune Evasion and Impedes Anti-PD-1 Treatment by Inducing Immunosuppressive Macrophages in Gastric Cancer. Cancer Immunol Res 10, 1506–1524 (2022).

68. Hong, C. et al. cGAS–STING drives the IL-6-dependent survival of chromosomally instable cancers. Nature 607, 366–373 (2022).

69. Wang, S. et al. Intratumoral injection of a CpG oligonucleotide reverts resistance to PD-1 blockade by expanding multifunctional CD8+ T cells. Proc. Natl. Acad. Sci. U. S. A. 113, E7240–E7249 (2016).

70. Sato-Kaneko, F., et al. Combination immunotherapy with TLR agonists and checkpoint inhibitors suppresses head and neck cancer. JCI Insight 2, (2017).

71. Veneziani, I. et al. Toll-like receptor 8 agonists improve NK-cell function primarily targeting CD56brightCD16-subset. J Immunother Cancer 10, (2022).

72. Dias Costa, A., et al. Neoadjuvant Chemotherapy Is Associated with Altered Immune Cell Infiltration and an Anti-Tumorigenic Microenvironment in Resected Pancreatic Cancer. Clin. Cancer Res. 28, 5167–5179 (2022).

73. Grout, J. A. et al. Spatial Positioning and Matrix Programs of Cancer-Associated Fibroblasts Promote T-cell Exclusion in Human Lung Tumors. Cancer Discov. 12, 2606–2625 (2022).

74. Bortolomeazzi, M. et al. Immunogenomics of Colorectal Cancer Response to Checkpoint Blockade: Analysis of the KEYNOTE 177 Trial and Validation Cohorts. Gastroenterology vol. 161 1179–1193 Preprint at https://doi.org/10.1053/j.gastro.2021.06.064 (2021).

75. Biswas, S. K. & Mantovani, A. Macrophage plasticity and interaction with lymphocyte subsets: cancer as a paradigm. Nat. Immunol. 11, 889–896 (2010).

76. Ruffell, B. et al. Macrophage IL-10 blocks CD8+ T cell-dependent responses to chemotherapy by suppressing IL-12 expression in intratumoral dendritic cells. Cancer Cell 26, 623–637 (2014).

77. Peranzoni, E. et al. Macrophages impede CD8 T cells from reaching tumor cells and limit the efficacy of anti-PD-1 treatment. Proc. Natl. Acad. Sci. U. S. A. 115, E4041–E4050 (2018).

78. Fan, Y. et al. Epithelial SOX9 drives progression and metastases of gastric adenocarcinoma by promoting immunosuppressive tumour microenvironment. Gut (2022) doi:10.1136/gutjnl-2021-326581.

79. Zhang, L. et al. Single-Cell Analyses Inform Mechanisms of Myeloid-Targeted Therapies in Colon Cancer. Cell 181, 442–459.e29 (2020).

80. Liu, Y. et al. Immune phenotypic linkage between colorectal cancer and liver metastasis. Cancer Cell 40, 424–437.e5 (2022).

81. Yaddanapudi, K. et al. Control of tumor-associated macrophage alternative activation by macrophage migration inhibitory factor. J. Immunol. 190, 2984–2993 (2013).

82. Sivakumar, S. et al. Tissue and liquid biopsy profiling reveal convergent tumor evolution and therapy evasion in breast cancer. Nat. Commun. 13, 7495 (2022).

83. Kansara, M. et al. Early circulating tumor DNA dynamics as a pan-tumor biomarker for long-term clinical outcome in patients treated with durvalumab and tremelimumab. Mol. Oncol. (2022) doi:10.1002/1878-0261.13349.

84. Reichert, Z. R. et al. Prognostic value of plasma circulating tumor DNA fraction across four common cancer types: a real-world outcomes study. Ann. Oncol. 34, 111–120 (2023).

85. Maron, S. B. et al. Circulating Tumor DNA Sequencing Analysis of Gastroesophageal Adenocarcinoma. Clin. Cancer Res. 25, 7098–7112 (2019).

86. Jee, J. et al. Overall survival with circulating tumor DNA-guided therapy in advanced non-small-cell lung cancer. Nat. Med. 28, 2353–2363 (2022).

87. Sartore-Bianchi, A. et al. Circulating tumor DNA to guide rechallenge with panitumumab in metastatic colorectal cancer: the phase 2 CHRONOS trial. Nat. Med. 28, 1612–1618 (2022).

88. Santhanam, B., Oikonomou, P. & Tavazoie, S. Systematic assessment of prognostic molecular features across cancers. Cell Genomics (2023).

89. Ho, W. W., Pittet, M. J., Fukumura, D. & Jain, R. K. The local microenvironment matters in preclinical basic and translational studies of cancer immunology and immunotherapy. Cancer Cell 40, 701–702 (2022).

90. Reardon, D. A. et al. Glioblastoma Eradication Following Immune Checkpoint Blockade in an Orthotopic, Immunocompetent Model. Cancer Immunol Res 4, 124–135 (2016).

91. Horton, B. L., et al. Lack of CD8+ T cell effector differentiation during priming mediates checkpoint blockade resistance in non-small cell lung cancer. Sci Immunol 6, eabi8800 (2021).

92. Leibold, J. et al. Somatic mouse models of gastric cancer reveal genotype-specific features of metastatic disease. bioRxiv 2022.06.15.494941 (2022) doi:10.1101/2022.06.15.494941.

93. Li, H. & Durbin, R. Fast and accurate short read alignment with Burrows-Wheeler transform. Bioinformatics 25, 1754–1760 (2009).

94. McKenna, A. et al. The Genome Analysis Toolkit: a MapReduce framework for analyzing next-generation DNA sequencing data. Genome Res. 20, 1297–1303 (2010).

95. Cibulskis, K. et al. Sensitive detection of somatic point mutations in impure and heterogeneous cancer samples. Nat. Biotechnol. 31, 213–219 (2013).

96. Sherry, S. T. et al. dbSNP: the NCBI database of genetic variation. Nucleic Acids Res. 29, 308–311 (2001).

97. 1000 Genomes Project Consortium et al. A global reference for human genetic variation. Nature 526, 68–74 (2015).

98. International HapMap 3 Consortium, et al. Integrating common and rare genetic variation in diverse human populations. Nature 467, 52–58 (2010).

99. McLaren, W. et al. The Ensembl Variant Effect Predictor. Genome Biol. 17, 122 (2016).

100. Dobin, A. et al. STAR: ultrafast universal RNA-seq aligner. Bioinformatics 29, 15–21 (2013).

101. Li, B. & Dewey, C. N. RSEM: accurate transcript quantification from RNA-Seq data with or without a reference genome. BMC Bioinformatics 12, 323 (2011).

102. Hänzelmann, S., Castelo, R. & Guinney, J. GSVA: gene set variation analysis for microarray and RNA-seq data. BMC Bioinformatics 14, 7 (2013).

103. Stuart, T. et al. Comprehensive Integration of Single-Cell Data. Cell 177, 1888–1902.e21 (2019).

104. Wolock, S. L., Lopez, R. & Klein, A. M. Scrublet: Computational Identification of Cell Doublets in Single-Cell Transcriptomic Data. Cell Syst 8, 281–291.e9 (2019).

105. Korsunsky, I. et al. Fast, sensitive and accurate integration of single-cell data with Harmony. Nat. Methods 16, 1289–1296 (2019).

